# Myocarditis and Pericarditis following COVID-19 Vaccination: Evidence Syntheses on Incidence, Risk Factors, Natural History, and Hypothesized Mechanisms

**DOI:** 10.1101/2022.02.28.22271643

**Authors:** Jennifer Pillay, Lindsay Gaudet, Aireen Wingert, Liza Bialy, Andrew S. Mackie, D. Ian Paterson, Lisa Hartling

**Author notes:** **Declaration of competing interests:** The authors have no competing interests to declare.

## Abstract

**Objectives:** Myocarditis and pericarditis are adverse events of special interest after vaccination for COVID-19. Evidence syntheses were conducted on incidence rates, risk factors for myocarditis and pericarditis after COVID-19 mRNA vaccination, clinical presentation and short- and longer-term outcomes of cases, and proposed mechanisms and their supporting evidence.

**Design:** Systematic reviews and evidence reviews.

**Data sources:** Medline, Embase and the Cochrane Library were searched from October 2020 to January 10, 2022; reference lists and grey literature (to January 13, 2021).

**Review methods:** Large (>10,000) or population-based/multisite observational studies and surveillance data (incidence and risk factors) reporting on confirmed myocarditis or pericarditis after COVID-19 vaccination; case series (n≥5, presentation, short-term clinical course and longer-term outcomes); opinions/letters/reviews/primary studies focused on describing or supporting hypothesized mechanisms. A single reviewer completed screening and another verified 50% of exclusions, using a machine-learning program to prioritize records. A second reviewer verified all exclusions at full text, extracted data, and (for incidence and risk factors) risk of bias assessments using modified Joanna Briggs Institute tools. Team consensus determined certainty of evidence ratings for incidence and risk factors using GRADE.

**Results:** 46 studies were included (14 on incidence, 7 on risk factors, 11 on characteristics and short-term course, 3 on longer term outcomes, and 21 on mechanisms). Incidence of myocarditis after mRNA vaccines is highest in male adolescents and young adults (12-17y: range 50-139 cases per million [low certainty] and 18-29y: range 28-147 per million [moderate certainty]). For 5-11 year-old males and females and females 18-29 years of age, incidence of myocarditis after vaccination with Pfizer may be fewer than 20 cases per million (low certainty). There was very low certainty evidence for incidence after a third dose of an mRNA vaccine. For 18-29 year-old males and females, incidence of myocarditis is probably higher after vaccination with Moderna compared to Pfizer (moderate certainty). Among 12-17, 18-29 and 18-39 year-olds, incidence of myocarditis/pericarditis after dose 2 of an mRNA vaccine may be lower when administered ≥31 days compared to ≤30 days after dose 1 (low certainty). Data specific to males aged 18-29 indicated that the dosing interval may need to increase to ≥56 days to substantially drop incidence. For clinical course and short-term outcomes only one small series (n=8) was found for 5-11 year olds. In cases of adolescents and adults, the majority (>90%) of myocarditis cases involved 20-30 year-old males with symptom onset 2 to 4 days after second dose (71-100%). Most cases were hospitalized (≥84%) for a short duration (2-4 d). For pericarditis, data is limited but more variation has been reported in patient age, sex, onset timing and rate of hospitalization. Case series with longer-term (3 mo; n=38) follow-up suggest persistent ECG abnormalities, as well as ongoing symptoms and/or a need for medications or restriction from activities in >50% of patients. 16 hypothesized mechanisms are described, with little direct supporting or refuting evidence.

**Conclusions:** Adolescent and young adult males are at the highest risk of myocarditis after mRNA vaccination. Pfizer over Moderna and waiting more than 30 days between doses may be preferred for this population. Incidence of myocarditis in children aged 5-11 may be very rare but certainty was low. Data on clinical risk factors was very limited. Clinical course of mRNA related myocarditis appears to be benign although longer term follow-up data is limited. Prospective studies with appropriate testing (e.g., biopsy, tissue morphology) will enhance understanding of mechanism(s).

**Funding and Registration no:** This project was funded in part by the Canadian Institutes of Health Research (CIHR) through the COVID-19 Evidence Network to support Decision-making (COVID-END) at McMaster University. Not registered.

**Summary box:** **What is already known about this topic?**

Case reports and surveillance signals of myocarditis (inflammation of the heart muscle) and pericarditis (inflammation of the two-layered sac surrounding the heart) after COVID-19 vaccination appeared as early as April 2021.

These have prompted ongoing surveillance and research of these complications to investigate their incidence, possible attribution to the vaccines, and clinical course.

**What this study adds**

This review critically appraises and synthesizes the available evidence to-date on the incidence of and risk factors for myocarditis and pericarditis after COVID-19 vaccination in multiple countries. It summarizes the presentation and clinical course of over 8000 reported cases and describes some initial reports of longer term outcomes. Further, many possible mechanisms are outlined and discussed.

Though low, the incidence of myocarditis is probably the highest in young males aged 12-29 years and is probably higher with Moderna than Pfizer mRNA vaccines. Longer dosing intervals may be beneficial. Most cases are mild and self-limiting, though data in 5-11 year-olds is very limited. Continued active surveillance with longer term follow-up is warranted.

## Introduction

Case reports and surveillance signals^1–4^ of myocarditis (including myopericarditis) and pericarditis after COVID-19 vaccination appeared as early as April 2021, leading to the surveillance of adverse events of special interest following vaccination with messenger RNA (mRNA) vaccines manufactured by Pfizer and Moderna. Estimated rates of myocarditis in the United Kingdom (UK)^5^ and the United States (US)^6 7^ are 11 and 1-2 cases per 100,000 person-years, respectively, regardless of age. Estimated background/expected rates of myocarditis and pericarditis following COVID-19 vaccination in the US, that is, adjusted for a 7-day risk period where most cases appear, are 0.2 and 1.4 per 1 million people, respectively.^6^ Historically, myocarditis has been more prevalent in males than females, from childhood through young adulthood,^8^ and this tendency was reflected in early post-COVID-19 vaccination case series.^9 10^

Symptoms indicative of myocarditis or pericarditis typically include new onset and persisting chest pain, shortness of breath, and/or palpitations.^11^ Diagnosis of a probable case of myocarditis usually requires elevated troponin levels and/or findings from imaging (e.g., echocardiography, magnetic resonance imaging) or other testing (e.g., ECG); histopathology for a definitive diagnosis is not usually performed.^12^ Differential diagnoses, including COVID-19 infection and community-acquired myocarditis, should be considered and ruled out.^12 13^ Most people experiencing these conditions will fully recover; however, in rare cases, myocarditis can lead to heart failure or asymptomatic left ventricular dysfunction. Long-term consequences associated with pericarditis include one or more recurrences and, rarely, thickening of the pericardium and constrictive heart filling.^14^

This evidence synthesis included systematic reviews of incidence rates (0 to 39 years) and risk factors for myocarditis and pericarditis after COVID-19 mRNA vaccination, evidence reviews of initial presentation and clinical course and of longer-term outcomes of cases, and a description of proposed mechanisms and their supporting evidence. Our original reviews (literature search October 6, 2021) on incidence rates, risk factors, and case presentation and short-term outcomes of myo- and/or pericarditis after COVID-19 vaccination across all vaccines, ages and sexes indicated that incident rates were little-to-none after non-mRNA vaccines and in adults 40 years and older, that rates after mRNA vaccination varied by sex, age, and dose, and that the case characteristics and clinical course of myocarditis and pericarditis differ.^15^ There were no data on children aged <12 years or for people after receiving a third dose. For our review update reported herein, we refined the questions to focus on gaps and stakeholder/decision-maker priorities, specifically, mRNA vaccines, priority age and risk groups, and cases after a third vaccine dose. Where possible, we limited cases to those confirmed by medical record review and of myocarditis/myopericarditis or pericarditis rather than in combination (when data were available separately). We also expanded the scope to include evidence on longer-term outcomes. Finally, we added descriptions and studies of hypothesized mechanisms. This report focuses on the refined questions but includes findings on characteristics and short-term clinical course of myocarditis or pericarditis after any dose of the vaccine across all ages from the original review.

### Review Questions

1. What is the incidence of myocarditis and pericarditis following mRNA COVID-19 vaccination, by age and sex, in i) people 0-4 years, 5-11 years, 12-17 years, 18-29 years after their second dose, ii) recipients of any age after a third dose, and iii) individuals with prior history of myocarditis following mRNA COVID-19 vaccination?
2. Among individuals of a similar age and sex, are there risk or protective factors (e.g., pre-existing conditions [e.g. cardiac diseases, immunocompromise], previous SARS-CoV-2 infection [symptomatic or asymptomatic] or other viral infections, pharmacotherapies [e.g., hormones], type of vaccine product, length of vaccine dosing interval, vaccine combination for first vs second vs booster doses) for myocarditis and pericarditis following mRNA COVID-19 vaccination?
3. a. What are the characteristics and short-term clinical course of myocarditis or pericarditis after COVID-19 vaccination, in i) children <12 yrs, ii) recipients of any age after a third dose, and iii) individuals with prior history of myocarditis following mRNA COVID-19 vaccination?
b. (From original review) What are the characteristics and short-term clinical course of myocarditis or pericarditis after COVID-19 vaccination (i.e., across all ages, after a second vaccine dose)
4. Among individuals of a similar age and sex who experienced myocarditis or pericarditis after mRNA COVID-19 vaccination, what is the longer term (≥4 weeks) prognosis, and does this vary by patient or vaccine characteristics?
5. What are the hypothesized mechanisms involved in myocarditis and pericarditis following vaccination with mRNA COVID-19 vaccines, and do they vary by group?

## Methods

We conducted these evidence syntheses following a protocol developed before screening began. Knowledge users from the Public Health Agency of Canada (PHAC) contributed to scoping the reviews but did not participate in their conduct. We followed guidance for systematic reviews when conducting^16^ and reporting^17^ questions 1 and 2.

### Literature Search

We worked with an experienced medical information specialist to develop the search strategy, which was peer-reviewed (see Acknowledgements).^18^ Searches combined concepts for COVID-19, vaccines, and myocarditis/pericarditis/cardiovascular manifestations/adverse events/surveillance; each concept included various key word and Medical Subject Heading (MeSH) terms. The search was originally run on October 6, 2021 and an update was run January 10, 2022; the update search (**Appendix 1**) was slightly modified from the original search (e.g., adding Omicron, removing non-mRNA vaccine terms). We did not add limits for language, country, or study design, but had limits for case reports and news/newspaper articles. Using the multifile option as well as the deduping tool in Ovid, we searched Ovid MEDLINE(R) and Epub Ahead of Print, In-Process, In-Data-Review & Other Non-Indexed Citations and Daily <1946 to January 10, 2022> and Embase <1974 to 2022 January 10>. We searched grey literature by scanning 20 key national websites (e.g., Public Health Agency of Canada, UK’s Medicines & Healthcare Products Regulatory Agency, Centers for Disease Control and Prevention) to identify unpublished data. In January, we searched L-OVE, CT.gov, Cochrane COVID Reg, WHO Covid reg, and Google Scholar for additional grey literature. We scanned reference lists of included studies and systematic reviews, and consulted our clinician authors (AM and IP) and an expert consultant (Dr. Bruce McManus; see Acknowledgements) to identify potentially eligible studies on mechanisms. All studies/reports included in the original review were screened for eligibility for the revised questions in this update.

### Eligibility Criteria

Our inclusion criteria are outlined in **Table 1**.

**Table 1.** Eligibility Criteria for Each Question

|  |  |
| --- | --- |
| **Population** | People of any age; data for questions 1 and 2 must be reported using age categories (e.g., 0-4, 5-11, 12-17, 18-29, 30-39, ≥40 years) and by dose. If incidence rates were very low and for question 2 on risk factors, we included data across sexes if no other study reported by sex.<br>Incident rates after a second dose of vaccine in Q1 were intended to be limited to <30 year-olds; since several studies reported on an 18-39 year-old category we included this data, as well as that for 30-39 years olds, in a post hoc manner. |
| **Intervention/Exposure** | Q1: mRNA vaccines approved in Canada: BNT162b2 mRNA/PfizerBioNTech/Comirnaty, mRNA-1273/Maderna Spikevax (alternative manufacturers of same vaccine are eligible), by type of vaccine and dose.<br>Q2: Same as Q1, plus potential risk/protective factors: pre-existing conditions [e.g. cardiac diseases, immunocompromise], previous SARS-CoV-2 infection (symptomatic or asymptomatic) or other viral infections, length of vaccine dosing interval, type of vaccine, vaccine combination for first vs second vs booster doses.<br>Q3: Confirmed myocarditis or pericarditis after mRNA COVID-19 vaccination. For 3b case did not need to be confirmed.<br>Q4: Confirmed myocarditis or pericarditis after mRNA COVID-19 vaccination.<br>Q5: Confirmed myocarditis or pericarditis after mRNA COVID-19 vaccination.<br>At least one dose of the vaccine needed to be with an mRNA vaccine; one or more other doses may have been with a non-mRNA vaccine.<br>For Qs 3-5 we included exposure to myocarditis and/or pericarditis (combined) if there was no data on these separately. |
| **Control/Comparator** | Q1: People previously vaccinated with mRNA COVID-19 vaccine but no longer at risk for outcome, previously vaccinated with other vaccines (i.e., controlling for confounders associated with vaccine uptake), or unvaccinated people; or no comparator.<br>Q2: People vaccinated with mRNA COVID-19 vaccine but without the risk/protective factor. |
|  | Q3: No comparator.<br>Q4: No comparator, but will include data on any comparisons with people vaccinated and not experiencing myocarditis or pericarditis.<br>Q5: People previously vaccinated with mRNA COVID-19 vaccine who did not experience myocarditis or pericarditis; or no comparator. |
| **Outcome** | Q1: Incidence rate/cumulative risk of confirmed myocarditis (including myopericarditis) or pericarditis by dose. Effect measures: incidence rate/cumulative risk (may be risk difference if accounting for background rate in control group). Will include rates of myocarditis or pericarditis (reported collectively) if there are no data for these separately.<br>Q2: Ratio measures of incidence/reported events by risk/protective factor (e.g., RR or odds ratios), adjusted for key confounders (e.g., previous COVID-19 illness and severity) when reported. Will include rates of myocarditis or pericarditis (reported collectively) if there are no data for these separately.<br>Q3: Characteristics of the patients (e.g., age, sex, pre-existing conditions [e.g., cardiac diseases] and infections [e.g., recent/past SARS-CoV-2 infection], race/ethnicity) and case presentation (e.g., timing/dose/type of vaccine, diagnostics, illness severity, treatments provided, short-term outcomes).<br>Q4: As in Q3, plus any outcomes measured $\geq 4$ weeks after onset of myocarditis or pericarditis (e.g., re-hospitalization, functional capacity, chest pain).<br>Q5: Authors' summaries of any hypotheses or findings after investigating potential mechanisms (e.g., histology, experiments with viral spike glycoprotein of SARS-CoV-2 [encoded by mRNA vaccine]), gene panels, serology for innate and acquired immune system components, autoimmune antibodies). |
| **Setting** | Any setting and country.* |
| **Study design** | Q1: Large ($>10,000$ vaccinated people) sample or multisite/health system-based observational studies; reports or databases of confirmed case using surveillance data.<br>Q2: Observational studies (including case control studies) with $n \geq 10$ with the risk/protective factor; data for subset of people with myocarditis or pericarditis may come from passive reporting systems.<br>Q3: Case series $n \geq 5$; data may come from medical record review of cases reported to active or passive surveillance systems (if reporting more than age, sex, and dose and type of vaccine); for Q3b we relied first on case series reported in systematic reviews and added data on cases from more recent case series or studies included for incident rates if they reported sufficient data on clinical course.<br>Q4: Case series $n \geq 5$; data may come from medical record review of cases reported to active or passive surveillance systems.<br>Q5: Any primary study, systematic review, or expert opinion article/letter on the topic.<br>Letters and commentaries will be included if they provide sufficient data. |
| **Publication Language** | English full texts.<br>We will cite those excluded based on language. |
| **Publication Year & Status** | Oct 2020-onwards (vaccines were authorized mid-Sept 2020).<br>Pre-prints will be included. |

\* For questions 1 and 2, when reports collected data from similar/overlapping populations (e.g., US national surveillance data), we first included the study with the most recent data and then included additional studies if they differed in their methods (e.g., differing risk intervals [period after the vaccine where cases are counted]), differing age and/or sex stratification, use of unvaccinated or similar controls).

### Study Selection

All reviewers undertaking screening conducted a pilot round in Excel using 300 records. We then conducted screening in DistillerSR (Evidence Partners) using structured forms. For title and abstract review, we applied the machine learning program Daisy AI which continually reprioritizes records during screening.^19^ A single reviewer screened all titles/abstracts and another reviewer verified exclusions for the first 50% of records. For full text selection, a single reviewer reviewed all records, and a second reviewer verified all exclusions. Studies were further verified for inclusion during data extraction.

For question 3b, we mapped the case series by country and data source to identify the most recent and comprehensive data source for each geographic region (i.e., US, Canada, UK, Europe, and Israel).

For regions with multiple reports, we prioritized data that was most recent, most comprehensive (largest numbers), and that confirmed cases. We aimed to avoid overlap in data, i.e., the same cases reported in different sources. However, we did include sources that reported on specific sub-groups (e.g., 12 to 17 year-olds) to present more details on these groups of interest. We have noted in the evidence tables where there may be overlap in cases between reports and we avoided aggregating events/counts across studies.

### Data Extraction

We extracted all data into structured Word forms and conducted a pilot exercise with two studies for each question. Thereafter, one reviewer extracted data and another verified study eligibility and all data. Discrepancies were resolved by discussion or by a review lead (JP or LH).

For question 1, extracted data related to all elements of the eligibility criteria (Table 1) and data used for risk of bias assessment, focusing on methods for identifying cases (i.e., passive surveillance versus active surveillance/registry data), outcome ascertainment and confirmation/adjudication (including criteria for case definitions and classification), as well as the dosing interval and risk interval for which the events were captured. We preferred estimates of incidence compared with an unexposed group (i.e., excess incidence/risk differences) over those without a control. For question 2, we extracted data as for question 1, along with associations or subgroup analyses based on pre-existing conditions, different vaccine types/products, dosing interval, and combinations of vaccines. We extracted rates in each group and (if reported) the relative effects between groups (e.g., incidence rate ratio (IRR), odds ratios), adjusted for key confounders (i.e., infection status, cardiac and immunodeficiency/autoimmune conditions) when reported.

For questions 3 and 4, we based data extraction on the evidence table presented in an existing review^9^ as it covered most of the items specified under our outcomes in Table 1. We added the following items: criteria for confirmation of cases, breakdown of cases by diagnosis (myocarditis, pericarditis, myopericarditis), case source, age included, percent with pre-existing conditions, percent admitted to an intensive care unit (ICU), percent hospitalized, number of fatalities. For question 4, we also added longer-term investigations and outcomes.

For question 5, we extracted verbatim authors’ summaries of any hypotheses and, where available, findings by the authors or cited works investigating potential mechanisms (e.g., histology, gene panels, serology for innate and acquired immune system components, autoimmune antibodies, tissue biopsies, autopsy findings, etc.). We checked references used to support statements made by authors in proposing or explaining hypotheses to identify whether they provided direct empirical evidence (i.e., specific to COVID-19 mRNA vaccination). We involved three content experts who reviewed proposed mechanisms for comprehensiveness and interpretation.

### Risk of Bias Assessment

For questions 1 and 2, all reviewers involved in the risk of bias assessments piloted the risk of bias tool with two papers. We used the JBI (formerly Joanna Briggs Institute) checklist for cohort studies,^20^ with modifications mainly to ascertain valid risk factor measurement (**Appendix 2**). We focused on valid and reliable case finding and confirmation, and, for question 2, accounting for key confounders. Assessments were completed by one reviewer and verified by another. Discrepancies were resolved by discussion or by a review lead.

For questions 3 and 4, we did not assess the risk of bias of the included case series. The intent of these questions was to characterize these patients and their clinical course without providing quantitative estimates, e.g., of incidence. The main differentiating feature between existing case series is whether they only report on cases confirmed/verified by clinicians; we either limited inclusion to confirmed cases (questions 3a and 4) or extracted this information and considered this factor when summarizing the results (question 3b).

### Data Synthesis & Certainty Assessments

For question 1, we did not pool results due to heterogeneity in case finding (passive vs. active surveillance), dosing and risk intervals, age groupings, and, in some cases, some degree of overlap in cases expected between studies. We tabulated all results and compared and contrasted findings between studies based on the major differentiating population, vaccine and methodologic variables. We report the range in incidence rates across studies, or, when all rates are low, concluded that the incidence is <20 cases per million which, based on our clinical investigator input, we considered very rare. Based on input from clinicians and our knowledge users from PHAC we developed primary age categories (0-4, 5-11, 12-17, 18-29) to rely on when possible. If a study contributed more than one result within these (e.g., 20-24 and 25-29, results for each mRNA vaccine) we calculated a weighted average (e.g., by proportion of years in the age range) of the incident rates. When a study reported an incidence rate (or data to calculate this) and an IRR compared with a control/background rate, but not the difference in incidence (excess incidence over background rate), we calculated the excess incidence (i.e., crude incidence – [crude incidence/IRR]).

We assessed the certainty for each of our conclusion statements using Grading of Recommendations, Assessment, Development and Evaluation (GRADE) guidance.^21–23^ We did not apply a threshold for an important incidence rate, but in general considered fewer than 20 cases per million doses to be very rare. For question 2, associations ≥1.5 (odds ratio/relative risk) were considered clinically relevant (i.e., OR <1.5 shows “little-to-no association”). Observational studies for incident rates (question 1) started at low certainty (i.e., our confidence in the incident rate is limited; the true rate may be substantially different from the estimate) and studies on risk factors (question 2) started at high certainty^21^ (i.e., we are very confident that the true difference in rate between risk groups lies close to the estimate). We rated down for serious concerns about risk of bias, inconsistency/lack of consistency (single study), indirectness, imprecision, and/or reporting biases. We rated down for risk of bias when only studies having high risk for case ascertainment contributed to an outcome (e.g., passive surveillance where we assume there is under-ascertainment), and for indirectness for comparisons across both sexes or if the age group reported did not match one of our groups of interest (e.g., 13-39 year olds) and the incident rates may vary substantially among ages. We rated down for imprecision in groups with fewer people than would be expected for 1 event to occur based on estimated baseline incidence rates (e.g., a subgroup with 8000 people but reporting zero events). We considered uprating due to large incidence rates (e.g., twice or more than our 20 per million threshold) when no other major limitations were evident and we had moderate confidence in our conclusion, often reported as a range of incidences.^24^ In our conclusions, we have used “probably”, “may” and “uncertain” to reflect, respectively, moderate, low, or very low certainty evidence based on GRADE. When we have very low certainty we make no conclusions about specific incident rates or other outcomes.

For questions 3 and 4, we present the details for each case series in an evidence table and provide a descriptive summary. For question 5, we present a summary of the mechanisms and a more descriptive table.

### Patient and Public Involvement

Two patient/public partners joined the research team (see Acknowledgements). They were not involved in decisions about the questions or outcomes, which were determined by the funders. The research leads met with these partners to present and discuss the findings and their implications. The partners co-developed with the leads key messages from their perspective.

## Results

### Study Selection

After deduplication, our database searches on October 6 2021 and January 1 2022 retrieved 3,439 and 2,191 citations and other sources identified 18 and 805 citations. After title/abstract screening, we retrieved and screened full texts of 159 and 258 citations during each search. Forty-six studies were included in this update (**Figure 1**). Findings from 12 studies from the original review^15^ were carried forward (Q1=5^25–29^, Q2=2^29 30^, Q3b.^25–28 31–36^). From the search update, we identified 34 new reports across all questions (Q1=9^37–45^, Q2=5^37 41 45–47^, Q3=1^48^, Q4=3^39 49 50^, Q5=21^9 51–70^); we excluded five studies that only reported on combined myo- and/or pericarditis (as we located other studies with these reported separately),^71–75^ eight studies that did not report on confirmed cases,^76–83^ and three studies^84–86^ that did not contribute additional data to other included studies on the same cases. Tables of study characteristics, with all data on results, and risk of bias ratings for questions 1 and 2 are in **Appendix 2. Appendix 3** contains descriptions of the various national surveillance systems from which reports were collated or data collected.

**Figure 1.**
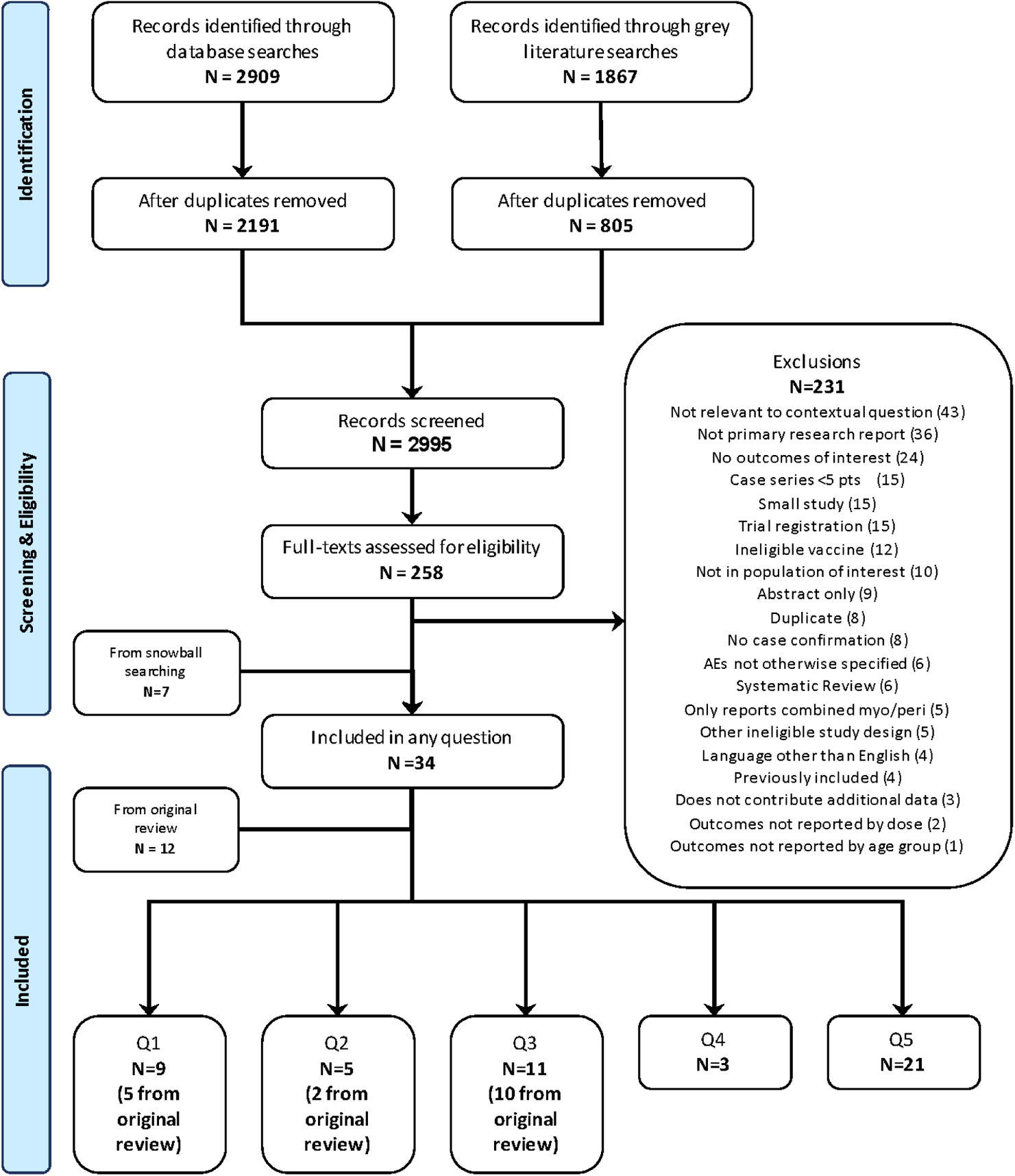
Literature Flow from Update Search

### Findings

#### Question 1: Incidence of Myocarditis and Pericarditis Following COVID-19 Vaccination

We included 14 studies (reported in publications, presentations, online reports, accessible data; all are considered here as “studies” for simplicity), with 8 using active surveillance data sources (e.g., medical records/ registries) and 6 using passive reporting systems. Data came from the US, Canada, the UK, Israel, Denmark, Singapore, and internationally (Moderna Global Safety Database). Four studies used the US VAERS data source for cases, and there was overlapping populations in analyses for 5-11^43 44^ and 12-17^25 29^ year olds where two reports were included for each because they applied different risk intervals (7-days and any duration). Four studies included a comparator group or background rate to estimate excess incidence rates. All studies on myocarditis reported on incident rates by age and sex; the only study reporting on pericarditis by age (5-11 year-olds) reported data across both sexes.^39^ Risk of bias was rated as some concerns (n=2) and high (n=6, mostly from lack of adjustment for variables other than age and sex) for studies using data from active surveillance systems and high for all six studies using passive surveillance data.

**Table 2** includes the results by study, the conclusions for each age and sex category, and the GRADE certainty assessments. We refer to studies by their source of adverse event data and the date of data collection; references for each study using this format are in **Appendix 3**.

**Table 2:**
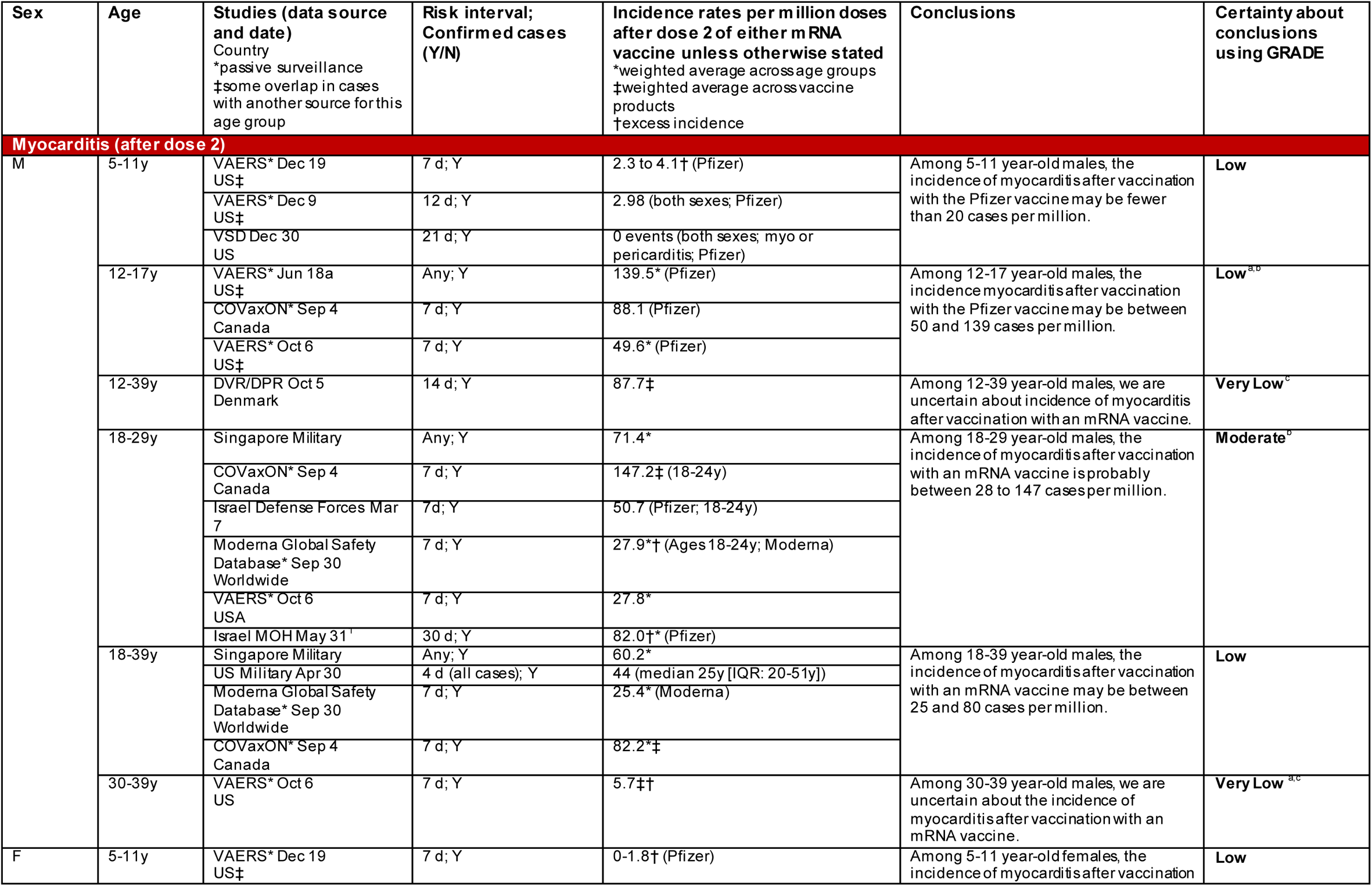
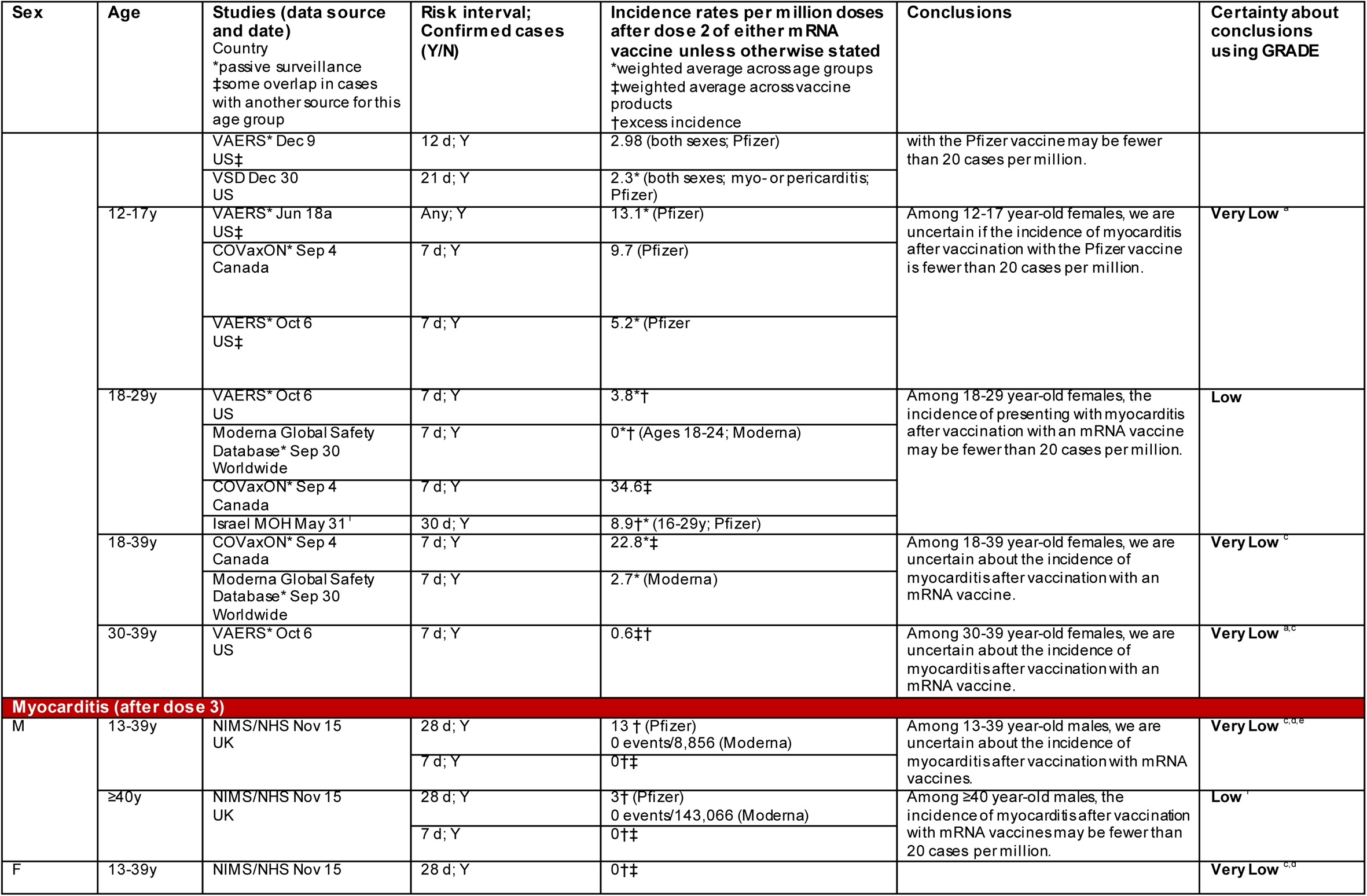
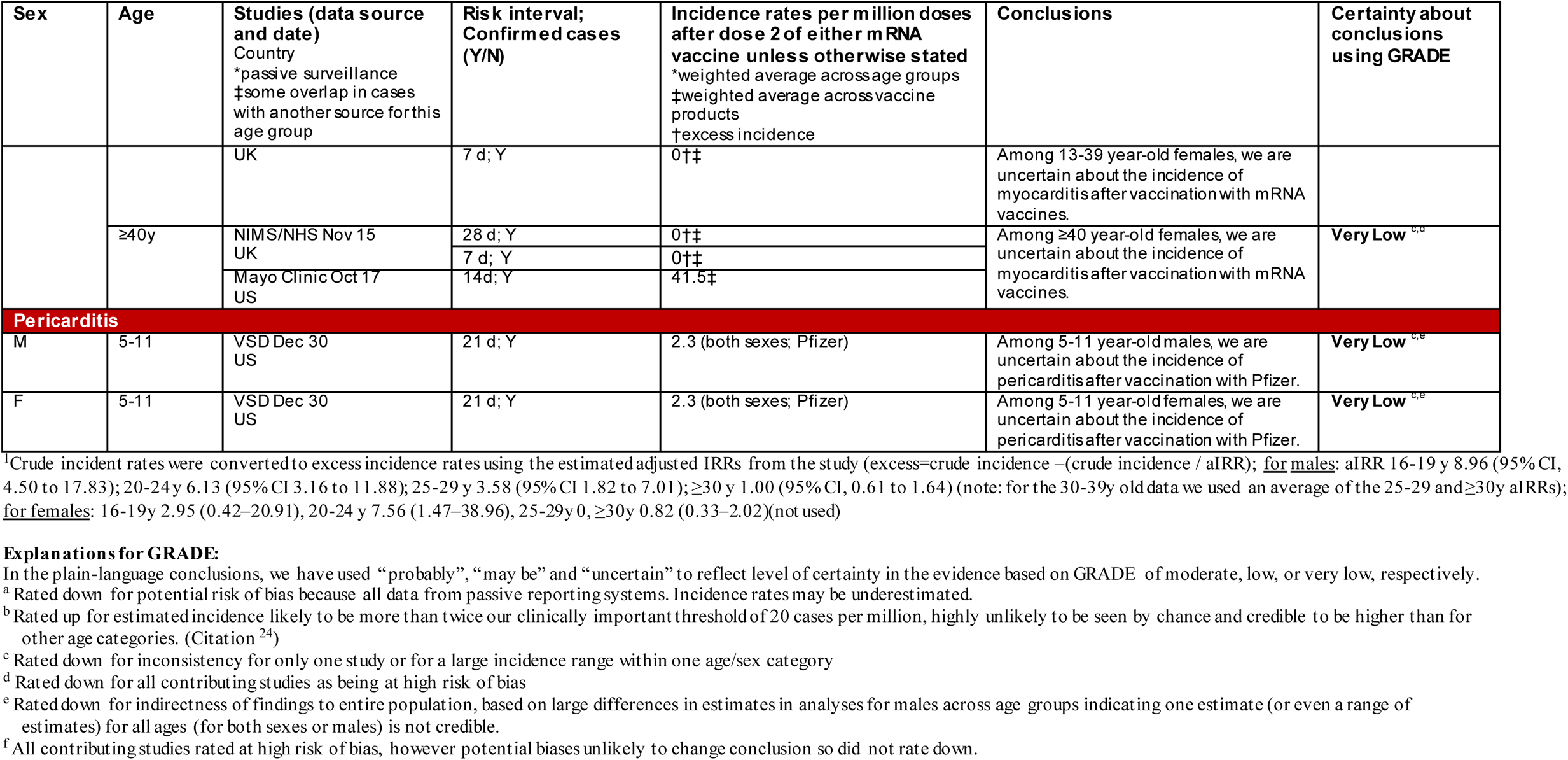
Summary of Findings for Incident Rates after Receipt of Either mRNA Vaccine (Question 1)

##### Myoc arditis after dose 2

For 5-11 year-old males and females, the incidence of myocarditis after vaccination with Pfizer may be fewer than 20 cases per million in both groups (low certainty). The incidence of myocarditis after mRNA vaccines is highest in male adolescents and young adults (12-17y: range 50-139 cases per million [low certainty] and 18-29y: range 28-147 per million [moderate certainty]). For 18-39 year-old males, the incidence of myocarditis may be between 25 and 82 cases per million (low certainty). Among females 18-29 years of age, the incidence of myocarditis after vaccination may be less than 20 cases per million (low certainty). We are very uncertain about the incidence of myocarditis after vaccination with mRNA vaccines in 12-17 year-old females, 30-39 year-old males and females, and in 18-39 year old females (very low certainty from risk of bias and inconsistency).

##### Myocarditis after dose 3

Among ≥40 year old males, the incidence of myocarditis after a third dose of an mRNA vaccines may be fewer than 20 cases per million (low certainty). For 13-39 year-old males or females and ≥40 year-old females, we are uncertain about the incidence of myocarditis after a third dose of an mRNA vaccine due to concerns about imprecision and inconsistency from multiple studies (very low certainty).

##### Peric ardit is

Based on a single study only reporting across both sexes, we are uncertain about the incidence of pericarditis after Pfizer vaccination in 5-11 year old males and females (very low certainty).

#### Question 2. Risk Factors

In this question, we assessed the relative differences in outcomes across subgroups. It is important to note, however, that these relative results must be taken in context with question 1 findings reporting on incidence; the relative differences in subgroups in females and older age groups should be given less weight in policy decision-making, based on the very low-to-no incidence of myocarditis after mRNA vaccination in these groups. **Table 3** includes the results by study (n=7), the conclusions for risk factor by age and (when available) sex, and the GRADE certainty assessments. We refer to studies by their source of adverse event data and the date of data collection; references for each study using this format are in **Appendix 3**.

**Table 3.**
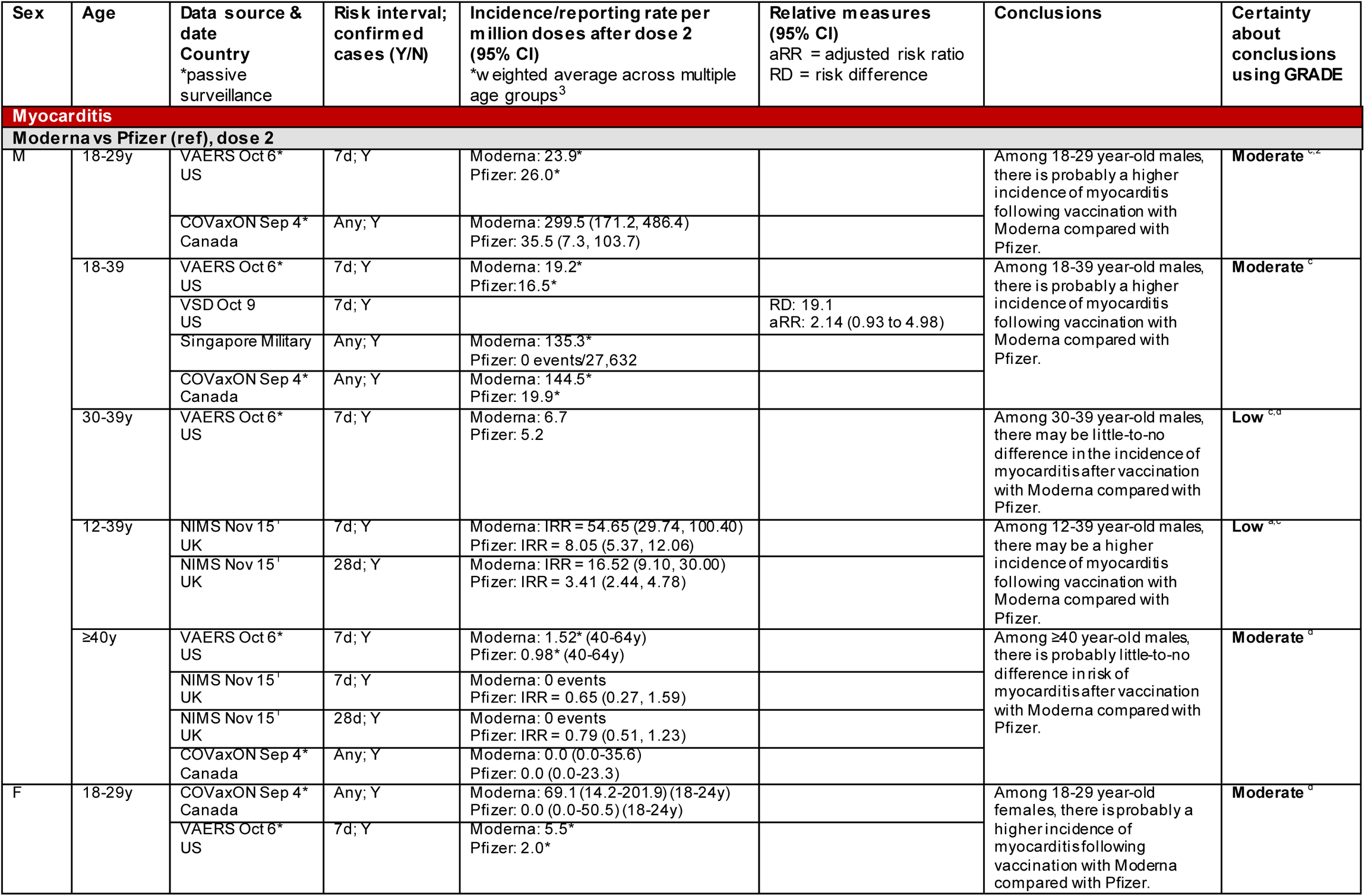
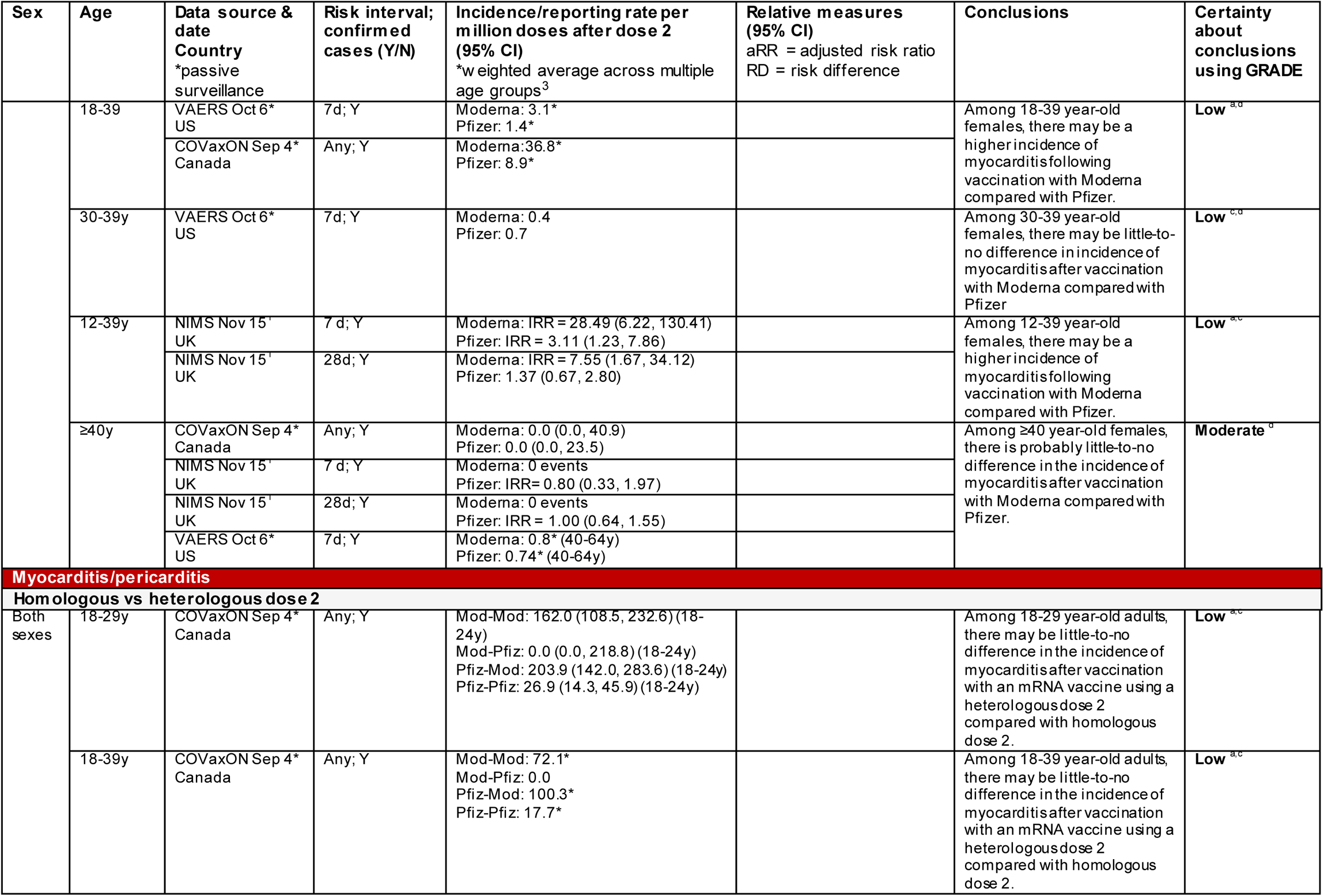
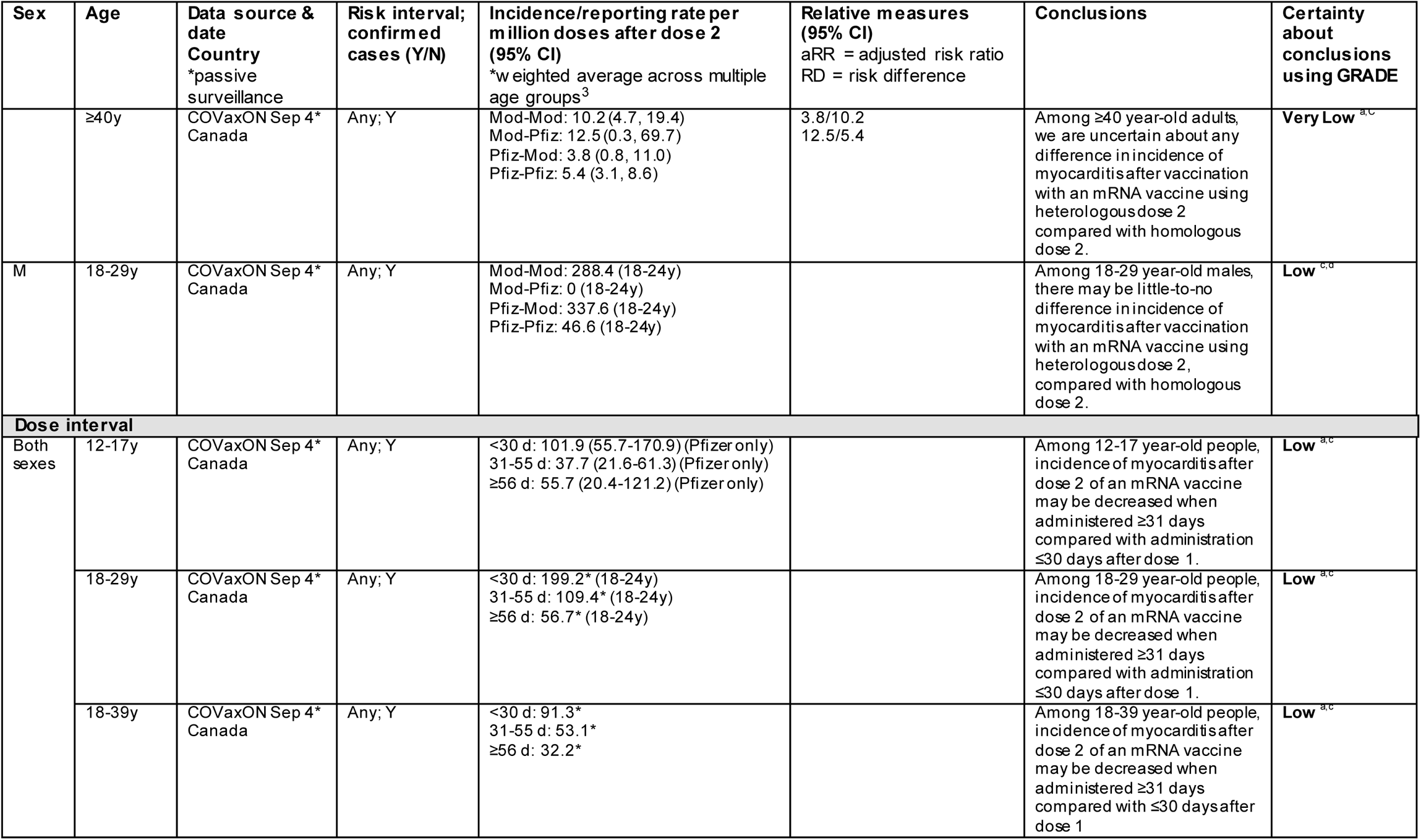
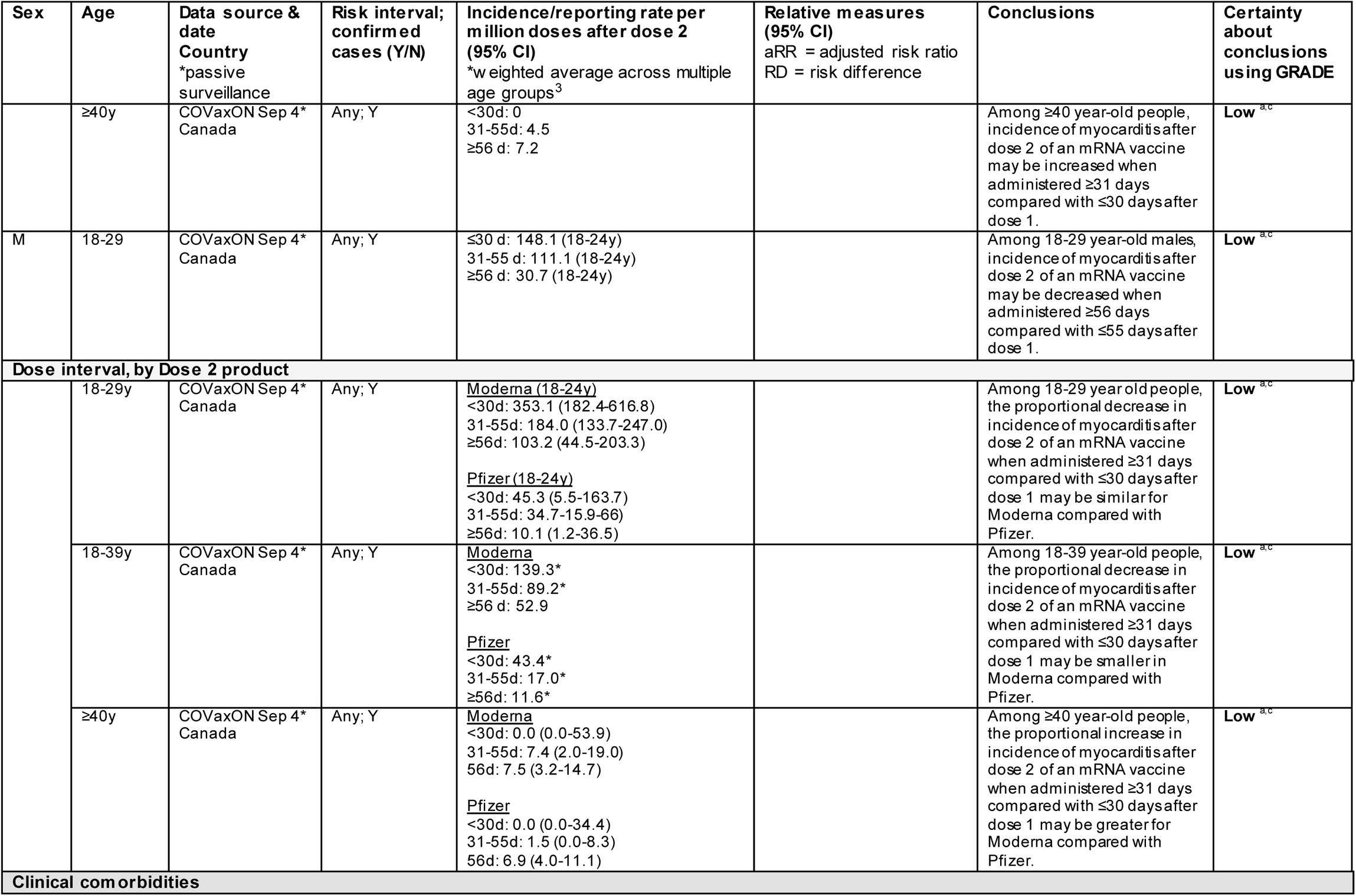
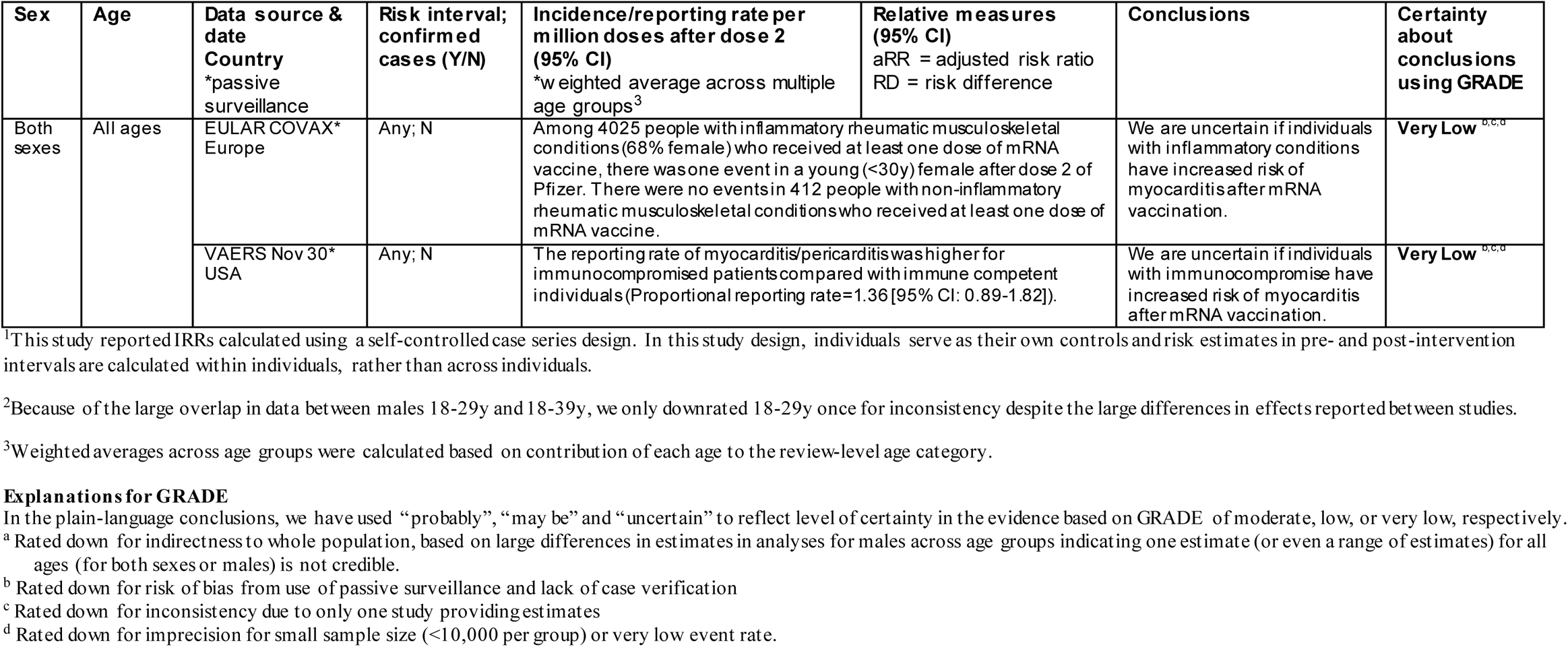
Summary of Findings for Possible Risk Factors for Myocarditis after mRNA Vaccination (Question 2)

#### Myoc arditis

##### Moderna versus Pfizer, after dose 2

For 18-29 year-old males and females, and 18-39 year-old males, the incidence of myocarditis is probably higher after vaccination with Moderna compared to Pfizer (moderate certainty). For 18-39 year-old females, the incidence of myocarditis may be higher after vaccination with Moderna compared to Pfizer (low certainty). For 30-39 year-old males and females, there may be little-to-no difference in the incidence of myocarditis after vaccination with Moderna compared to Pfizer (low certainty). Among ≥40 year-old males and females, there is probably little-to-no difference in risk of myocarditis after vaccination with Moderna compared to Pfizer (moderate certainty).

##### Myoc arditis and/or peric arditis

###### Homologous vs heterologous vaccine for dose 2

Among 18-29 and 18-39 year-old adults, and 18-29 year-old males, there may be little-to-no difference in the incidence of myocarditis/pericarditis after mRNA vaccination with a heterologous dose 2 (mixed Moderna and Pfizer with either for dose 2) compared to homologous dose 2 (low certainty). For adults ≥40 years old, we are uncertain about any difference in the incidence of myocarditis/pericarditis after mRNA vaccination with a heterologous dose 2 compared to homologous dose 2 (very low certainty).

###### Dose interval

Among 12-17, 18-29 and 18-39 year-old individuals, the incidence of myocarditis/pericarditis after dose 2 of an mRNA vaccine may be lower when administered ≥31 days compared to ≤30 days after dose 1 (low certainty). Data specific to males aged 18-29 indicated that the dosing interval may need to increase to ≥56 days to substantially drop incidence. For 18-29 year-olds, the proportional decrease in incidence of myocarditis/pericarditis after dose 2 of an mRNA vaccine when administered ≥31 days compared to ≤30 days after dose 1 may be similar for Moderna and Pfizer. This proportional decrease may be smaller with Moderna compared to Pfizer for 18-39 year olds (low certainty). Among ≥40 year-old people, incidence of myocarditis/pericarditis after dose 2 of an mRNA vaccine may be higher when administered ≥31 days compared to ≤30 days after dose 1 (low certainty). In this group, the proportional increase in incidence of myocarditis/pericarditis after dose 2 when administered ≥31 days compared to ≤30 days after dose 1 may be greater for Moderna compared to Pfizer.

###### Clinical comorbidities

We are uncertain if people with immunocompromise or inflammatory conditions have a different risk of myocarditis after mRNA vaccination (very low certainty from single studies using passive reporting systems without case confirmation and having inadequate sample sizes).

#### Question 3a. Case Characteristics and Short-term Clinical Course in <12 year-olds, After a Third Dose, or With Prior History of Myocarditis Following mRNA COVID-19 Vaccination

For this update question, we included one study (also included for question 1) reporting data on 5-11 year-olds with confirmed myocarditis.^43^ No study was found that reported on case presentation after a third dose or for those with a previous experience of myocarditis after a COVID-19 vaccine. In a series of 5-11 year olds receiving Pfizer (n=8), 50% were male and median age was 9 (range 6-11) years.

Race/ethnicity was not reported. Six children presented after their second dose and median time to present was 3 days. Seven presented with chest pain; of six tested three had abnormal ECG findings; and of five tested one had an abnormal echocardiogram. Of six with known outcomes, five had complete resolution of symptoms. Data is presented in the last column of **Table 4**.

**Table 4.**
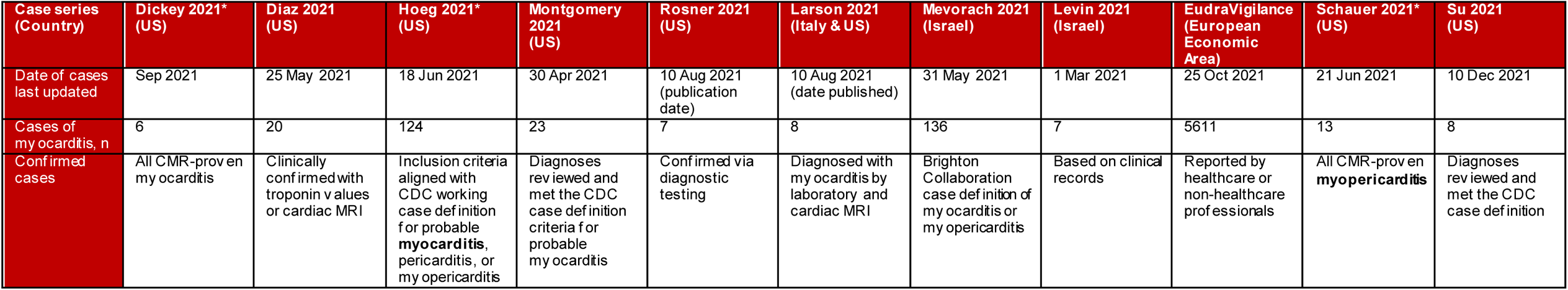
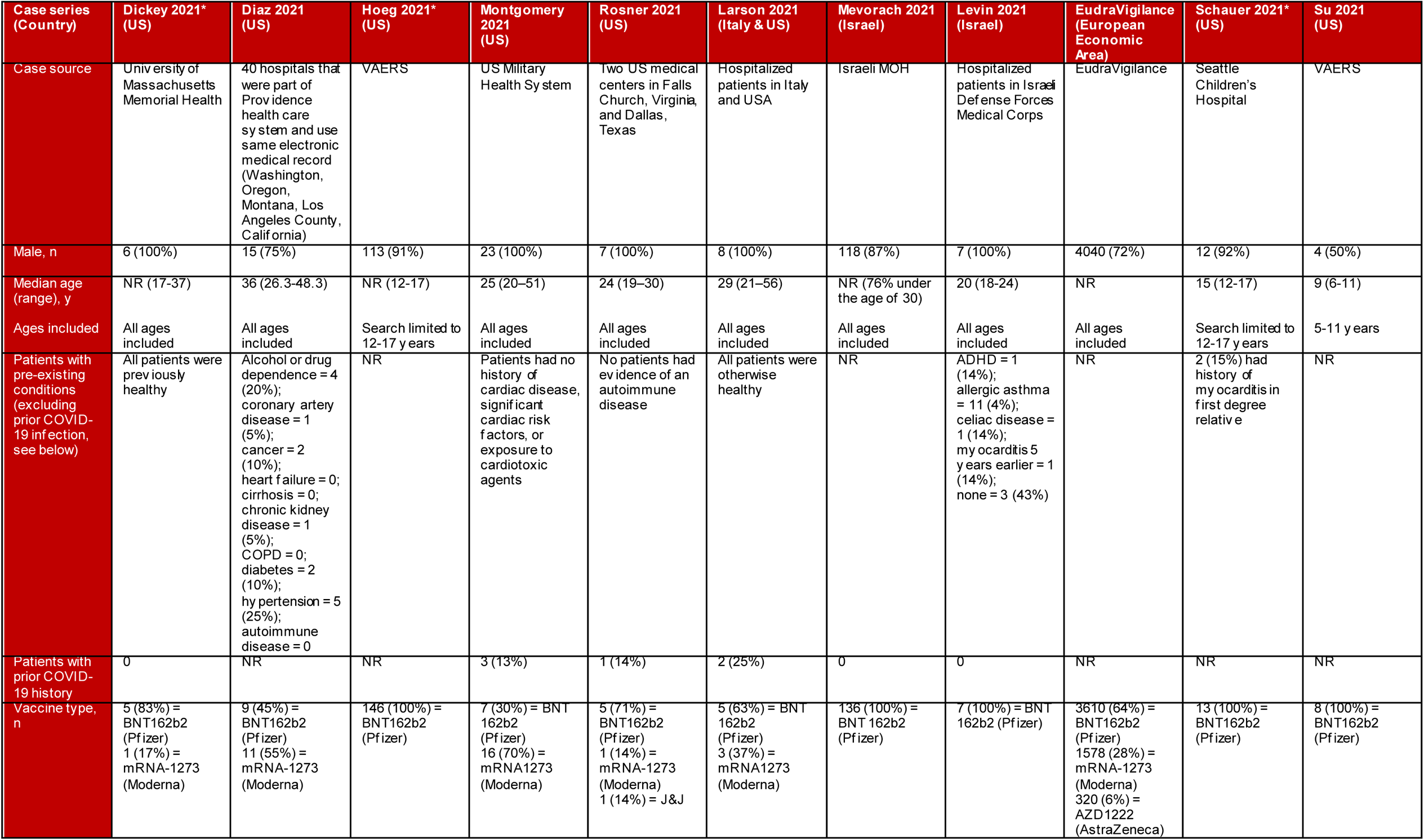
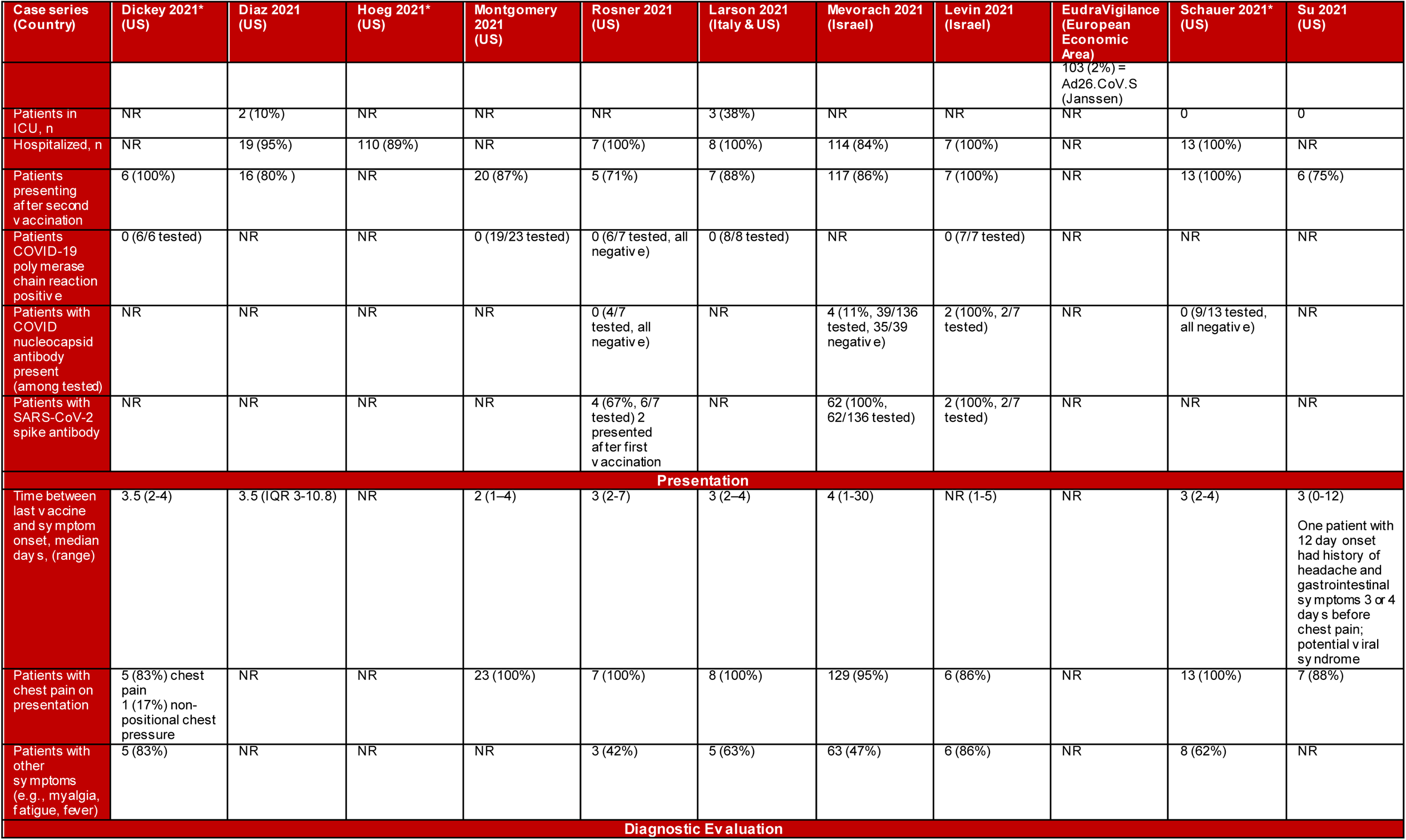
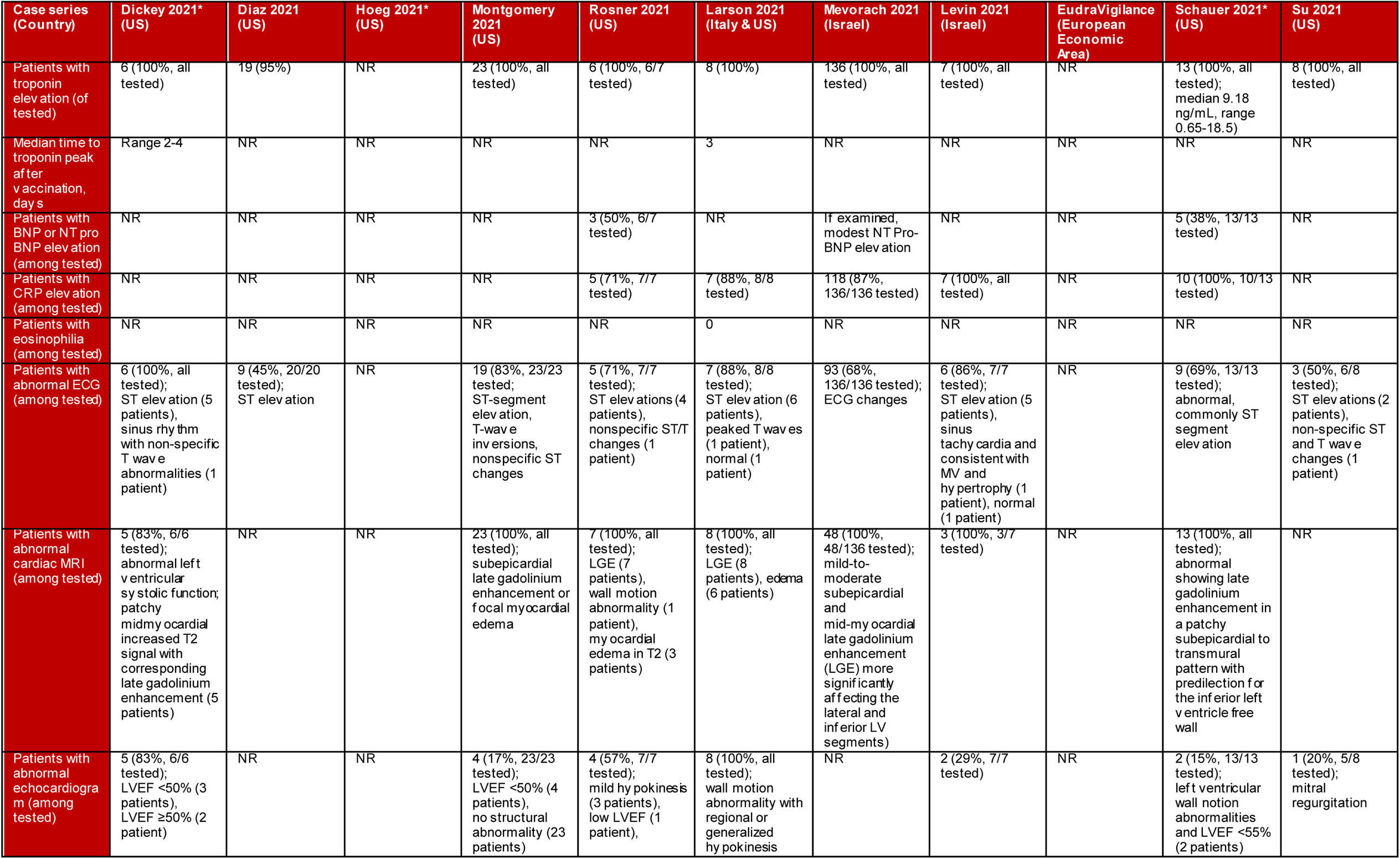
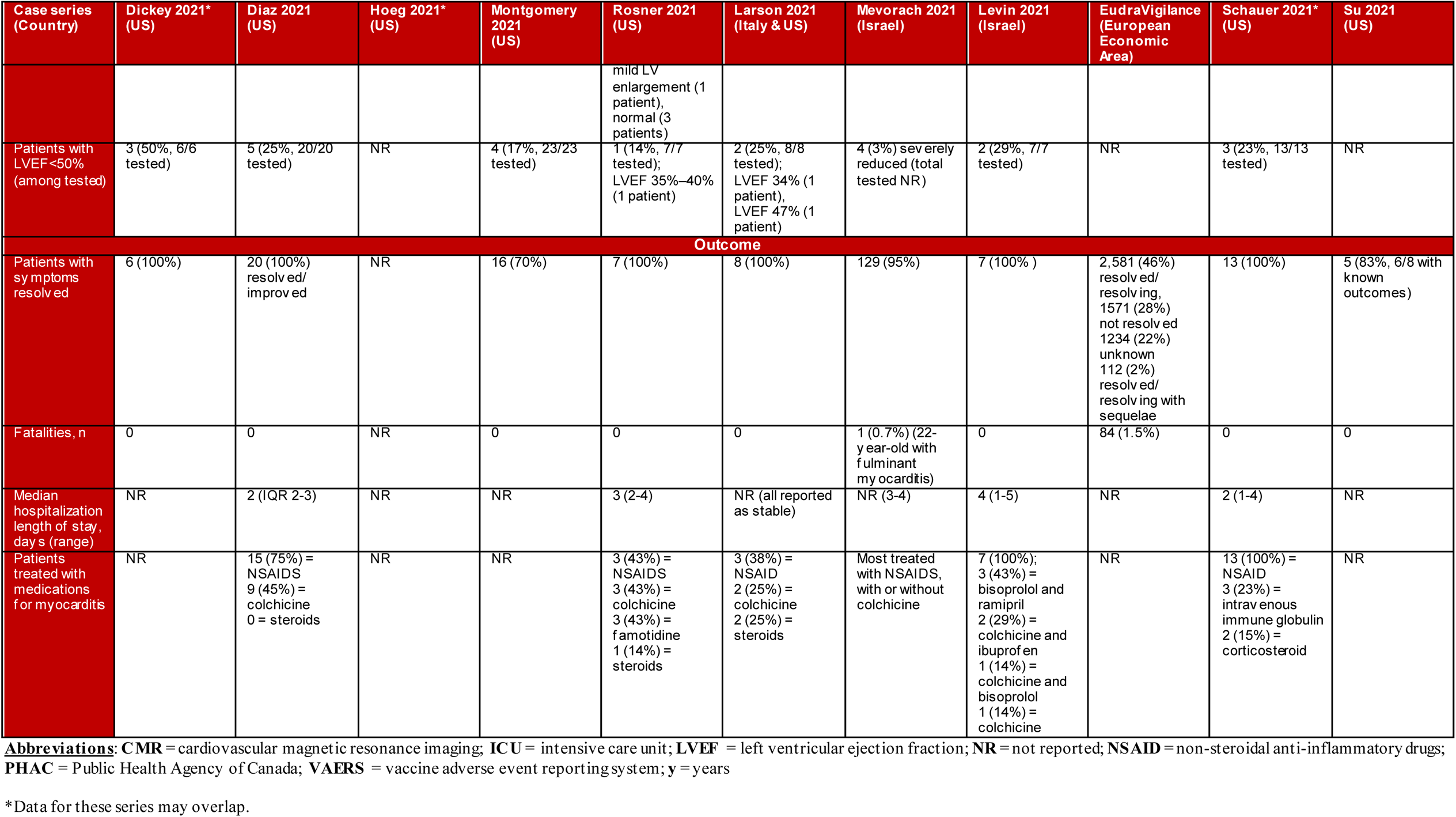
Results from Case Series with Short-term Follow-up of Myocarditis and Myopericarditis after COVID-19 Vaccination (Question 3a and b)

| Case series (Country) | Dickey 2021* (US) | Díaz 2021 (US) | Hoeg 2021* (US) | Montgomery 2021 (US) | Rosner 2021 (US) | Larson 2021 (Italy & US) | Mevorach 2021 (Israel) | Levin 2021 (Israel) | EudraVigilance (European Economic Area) | Schauer 2021* (US) | Su 2021 (US) |
| --- | --- | --- | --- | --- | --- | --- | --- | --- | --- | --- | --- |
| Date of cases last updated | Sep 2021 | 25 May 2021 | 18 Jun 2021 | 30 Apr 2021 | 10 Aug 2021 (publication date) | 10 Aug 2021 (date published) | 31 May 2021 | 1 Mar 2021 | 25 Oct 2021 | 21 Jun 2021 | 10 Dec 2021 |
| Cases of myocarditis, n | 6 | 20 | 124 | 23 | 7 | 8 | 136 | 7 | 5611 | 13 | 8 |
| Confirmed cases | All CMR-proven myocarditis | Clinically confirmed with troponin values or cardiac MRI | Inclusion criteria aligned with CDC working case definition for probable **myocarditis**, pericarditis, or myopericarditis | Diagnoses reviewed and met the CDC case definition criteria for probable myocarditis | Confirmed via diagnostic testing | Diagnosed with myocarditis by laboratory and cardiac MRI | Brighton Collaboration case definition of myocarditis or myopericarditis | Based on clinical records | Reported by healthcare or non-healthcare professionals | All CMR-proven **myopericarditis** | Diagnoses reviewed and met the CDC case definition |

| Case series (Country) | Dickey 2021* (US) | Diaz 2021 (US) | Hoeg 2021* (US) | Montgomery 2021 (US) | Rosner 2021 (US) | Larson 2021 (Italy & US) | Mevorach 2021 (Israel) | Levin 2021 (Israel) | EudraVigilance (European Economic Area) | Schauer 2021* (US) | Su 2021 (US) |
| --- | --- | --- | --- | --- | --- | --- | --- | --- | --- | --- | --- |
| Case source | University of Massachusetts Memorial Health | 40 hospitals that were part of Providence health care system and use same electronic medical record (Washington, Oregon, Montana, Los Angeles County, California) | VAERS | US Military Health System | Two US medical centers in Falls Church, Virginia, and Dallas, Texas | Hospitalized patients in Italy and USA | Israeli MOH | Hospitalized patients in Israeli Defense Forces Medical Corps | EudraVigilance | Seattle Children's Hospital | VAERS |
| Male, n | 6 (100%) | 15 (75%) | 113 (91%) | 23 (100%) | 7 (100%) | 8 (100%) | 118 (87%) | 7 (100%) | 4040 (72%) | 12 (92%) | 4 (50%) |
| Median age (range), y | NR (17-37) | 36 (26.3-48.3) | NR (12-17) | 25 (20-51) | 24 (19-30) | 29 (21-56) | NR (76% under the age of 30) | 20 (18-24) | NR | 15 (12-17) | 9 (6-11) |
| Ages included | All ages included | All ages included | Search limited to 12-17 years | All ages included | All ages included | All ages included | All ages included | All ages included | All ages included | Search limited to 12-17 years | 5-11 years |
| Patients with pre-existing conditions (excluding prior COVID-19 infection, see below) | All patients were previously healthy | Alcohol or drug dependence = 4 (20%); coronary artery disease = 1 (5%); cancer = 2 (10%); heart failure = 0; cirrhosis = 0; chronic kidney disease = 1 (5%); COPD = 0; diabetes = 2 (10%); hypertension = 5 (25%); autoimmune disease = 0 | NR | Patients had no history of cardiac disease, significant cardiac risk factors, or exposure to cardiotoxic agents | No patients had evidence of an autoimmune disease | All patients were otherwise healthy | NR | ADHD = 1 (14%); allergic asthma = 11 (4%); celiac disease = 1 (14%); myocarditis 5 years earlier = 1 (14%); none = 3 (43%) | NR | 2 (15%) had history of myocarditis in first degree relative | NR |
| Patients with prior COVID-19 history | 0 | NR | NR | 3 (13%) | 1 (14%) | 2 (25%) | 0 | 0 | NR | NR | NR |
| Vaccine type, n | 5 (83%) = BNT162b2 (Pfizer)<br>1 (17%) = mRNA-1273 (Moderna) | 9 (45%) = BNT162b2 (Pfizer)<br>11 (55%) = mRNA-1273 (Moderna) | 146 (100%) = BNT162b2 (Pfizer) | 7 (30%) = BNT162b2 (Pfizer)<br>16 (70%) = mRNA1273 (Moderna) | 5 (71%) = BNT162b2 (Pfizer)<br>1 (14%) = mRNA-1273 (Moderna)<br>1 (14%) = J&J | 5 (63%) = BNT162b2 (Pfizer)<br>3 (37%) = mRNA1273 (Moderna) | 136 (100%) = BNT162b2 (Pfizer) | 7 (100%) = BNT162b2 (Pfizer) | 3610 (64%) = BNT162b2 (Pfizer)<br>1578 (28%) = mRNA-1273 (Moderna)<br>320 (6%) = AZD1222 (AstraZeneca) | 13 (100%) = BNT162b2 (Pfizer) | 8 (100%) = BNT162b2 (Pfizer) |
|  |  |  |  |  |  |  |  |  | 103 (2%) = Ad26.CoV.S (Janssen) |  |  |
| Patients in ICU, n | NR | 2 (10%) | NR | NR | NR | 3 (38%) | NR | NR | NR | 0 | 0 |
| Hospitalized, n | NR | 19 (95%) | 110 (89%) | NR | 7 (100%) | 8 (100%) | 114 (84%) | 7 (100%) | NR | 13 (100%) | NR |
| Patients presenting after second v accination | 6 (100%) | 16 (80%) | NR | 20 (87%) | 5 (71%) | 7 (88%) | 117 (86%) | 7 (100%) | NR | 13 (100%) | 6 (75%) |
| Patients COVID-19 polymerase chain reaction positive | 0 (6/6 tested) | NR | NR | 0 (19/23 tested) | 0 (6/7 tested, all negative) | 0 (8/8 tested) | NR | 0 (7/7 tested) | NR | NR | NR |
| Patients with COVID nucleocapsid antibody present (among tested) | NR | NR | NR | NR | 0 (4/7 tested, all negative) | NR | 4 (11%, 39/136 tested, 35/39 negative) | 2 (100%, 2/7 tested) | NR | 0 (9/13 tested, all negative) | NR |
| Patients with SARS-CoV-2 spike antibody | NR | NR | NR | NR | 4 (67%, 6/7 tested) 2 presented after first v accination | NR | 62 (100%, 62/136 tested) | 2 (100%, 2/7 tested) | NR | NR | NR |
| Presentation |  |  |  |  |  |  |  |  |  |  |  |
| Time between last v accine and symptom onset, median days, (range) | 3.5 (2-4) | 3.5 (IQR 3-10.8) | NR | 2 (1-4) | 3 (2-7) | 3 (2-4) | 4 (1-30) | NR (1-5) | NR | 3 (2-4) | 3 (0-12)<br>One patient with 12 day onset had history of headache and gastrointestinal symptoms 3 or 4 days before chest pain; potential viral syndrome |
| Patients with chest pain on presentation | 5 (83%) chest pain<br>1 (17%) non-positional chest pressure | NR | NR | 23 (100%) | 7 (100%) | 8 (100%) | 129 (95%) | 6 (86%) | NR | 13 (100%) | 7 (88%) |
| Patients with other symptoms (e.g., myalgia, fatigue, fever) | 5 (83%) | NR | NR | NR | 3 (42%) | 5 (63%) | 63 (47%) | 6 (86%) | NR | 8 (62%) | NR |
| Diagnostic Evaluation |  |  |  |  |  |  |  |  |  |  |  |
| Patients with troponin elevation (of tested) | 6 (100%, all tested) | 19 (95%) | NR | 23 (100%, all tested) | 6 (100%, 6/7 tested) | 8 (100%) | 136 (100%, all tested) | 7 (100%, all tested) | NR | 13 (100%, all tested); median 9.18 ng/mL, range 0.65-18.5) | 8 (100%, all tested) |
| Median time to troponin peak after vaccination, days | Range 2-4 | NR | NR | NR | NR | 3 | NR | NR | NR | NR | NR |
| Patients with BNP or NT pro BNP elevation (among tested) | NR | NR | NR | NR | 3 (50%, 6/7 tested) | NR | If examined, modest NT Pro-BNP elevation | NR | NR | 5 (38%, 13/13 tested) | NR |
| Patients with CRP elevation (among tested) | NR | NR | NR | NR | 5 (71%, 7/7 tested) | 7 (88%, 8/8 tested) | 118 (87%, 136/136 tested) | 7 (100%, all tested) | NR | 10 (100%, 10/13 tested) | NR |
| Patients with eosinophilia (among tested) | NR | NR | NR | NR | NR | 0 | NR | NR | NR | NR | NR |
| Patients with abnormal ECG (among tested) | 6 (100%, all tested); ST elevation (5 patients), sinus rhythm with non-specific T wave abnormalities (1 patient) | 9 (45%, 20/20 tested); ST elevation | NR | 19 (83%, 23/23 tested); ST-segment elevation, T-wave inversions, nonspecific ST changes | 5 (71%, 7/7 tested); ST elevations (4 patients), nonspecific ST/T changes (1 patient) | 7 (88%, 8/8 tested); ST elevation (6 patients), peaked T waves (1 patient), normal (1 patient) | 93 (68%, 136/136 tested); ECG changes | 6 (86%, 7/7 tested); ST elevation (5 patients), sinus tachycardia and consistent with MV and hypertrophy (1 patient), normal (1 patient) | NR | 9 (69%, 13/13 tested); abnormal, commonly ST segment elevation | 3 (50%, 6/8 tested); ST elevations (2 patients), non-specific ST and T wave changes (1 patient) |
| Patients with abnormal cardiac MRI (among tested) | 5 (83%, 6/6 tested); abnormal left ventricular systolic function; patchy midmyocardial increased T2 signal with corresponding late gadolinium enhancement (5 patients) | NR | NR | 23 (100%, all tested); subepicardial late gadolinium enhancement or focal myocardial edema | 7 (100%, all tested); LGE (7 patients), wall motion abnormality (1 patient), myocardial edema in T2 (3 patients) | 8 (100%, all tested); LGE (8 patients), edema (6 patients) | 48 (100%, 48/136 tested); mild-to-moderate subepicardial and mid-myocardial late gadolinium enhancement (LGE) more significantly affecting the lateral and inferior LV segments) | 3 (100%, 3/7 tested) | NR | 13 (100%, all tested); abnormal showing late gadolinium enhancement in a patchy subepicardial to transmural pattern with predilection for the inferior left ventricle free wall | NR |
| Patients with abnormal echocardiogram (among tested) | 5 (83%, 6/6 tested); LVEF <50% (3 patients), LVEF ≥50% (2 patient) | NR | NR | 4 (17%, 23/23 tested); LVEF <50% (4 patients), no structural abnormality (23 patients) | 4 (57%, 7/7 tested); mild hypokinesis (3 patients), low LVEF (1 patient), mild LV enlargement (1 patient), normal (3 patients) | 8 (100%, all tested); wall motion abnormality with regional or generalized hypokinesis | NR | 2 (29%, 7/7 tested) | NR | 2 (15%, 13/13 tested); left ventricular wall motion abnormalities and LVEF <55% (2 patients) | 1 (20%, 5/8 tested); mitral regurgitation |
| Patients with LVEF <50% (among tested) | 3 (50%, 6/6 tested) | 5 (25%, 20/20 tested) | NR | 4 (17%, 23/23 tested) | 1 (14%, 7/7 tested); LVEF 35%–40% (1 patient) | 2 (25%, 8/8 tested); LVEF 34% (1 patient), LVEF 47% (1 patient) | 4 (3%) severely reduced (total tested NR) | 2 (29%, 7/7 tested) | NR | 3 (23%, 13/13 tested) | NR |
| **Outcome** |  |  |  |  |  |  |  |  |  |  |  |
| Patients with symptoms resolved | 6 (100%) | 20 (100%) resolved/improved | NR | 16 (70%) | 7 (100%) | 8 (100%) | 129 (95%) | 7 (100%) | 2,581 (46%) resolved/resolving, 1571 (28%) not resolved, 1234 (22%) unknown, 112 (2%) resolved/resolving with sequelae | 13 (100%) | 5 (83%, 6/8 with known outcomes) |
| Fatalities, n | 0 | 0 | NR | 0 | 0 | 0 | 1 (0.7%) (22-year-old with fulminant myocarditis) | 0 | 84 (1.5%) | 0 | 0 |
| Median hospitalization length of stay, days (range) | NR | 2 (IQR 2-3) | NR | NR | 3 (2-4) | NR (all reported as stable) | NR (3-4) | 4 (1-5) | NR | 2 (1-4) | NR |
| Patients treated with medications for myocarditis | NR | 15 (75%) = NSAIDs<br>9 (45%) = colchicine<br>0 = steroids | NR | NR | 3 (43%) = NSAIDs<br>3 (43%) = colchicine<br>3 (43%) = famotidine<br>1 (14%) = steroids | 3 (38%) = NSAID<br>2 (25%) = colchicine<br>2 (25%) = steroids | Most treated with NSAIDs, with or without colchicine | 7 (100%); 3 (43%) = bisoprolol and ramipril<br>2 (29%) = colchicine and ibuprofen<br>1 (14%) = colchicine and bisoprolol<br>1 (14%) = colchicine | NR | 13 (100%) = NSAID<br>3 (23%) = intravenous immune globulin<br>2 (15%) = corticosteroid | NR |

**Abbreviations:** CMR = cardiovascular magnetic resonance imaging; ICU = intensive care unit; LVEF = left ventricular ejection fraction; NR = not reported; NSAID = non-steroidal anti-inflammatory drugs; PHAC = Public Health Agency of Canada; VAERS = vaccine adverse event reporting system; y = years
\*Data for these series may overlap.

#### Question 3b. Case Characteristics and Short-term Clinical Course after the First or Second Dose in People of Any Age

From the original review, we report on 10 case series (6 published as case series^31–36^ and 4 included in Q1 but having relevant data on cases^25–28^) reporting on a total of 10,264 cases (median 22, range 6-5611) of myocarditis or pericarditis. All reports except the one using EudraVigilance data from the European Economic Area included clinician-confirmed cases. **Table 4** (myocarditis or myopericarditis) and **Table 5** (pericarditis) include data from the 10 case series. **Table 4** also contains data from the case series of 5-11 year-olds included for the update question 3a.

**Table 5.**
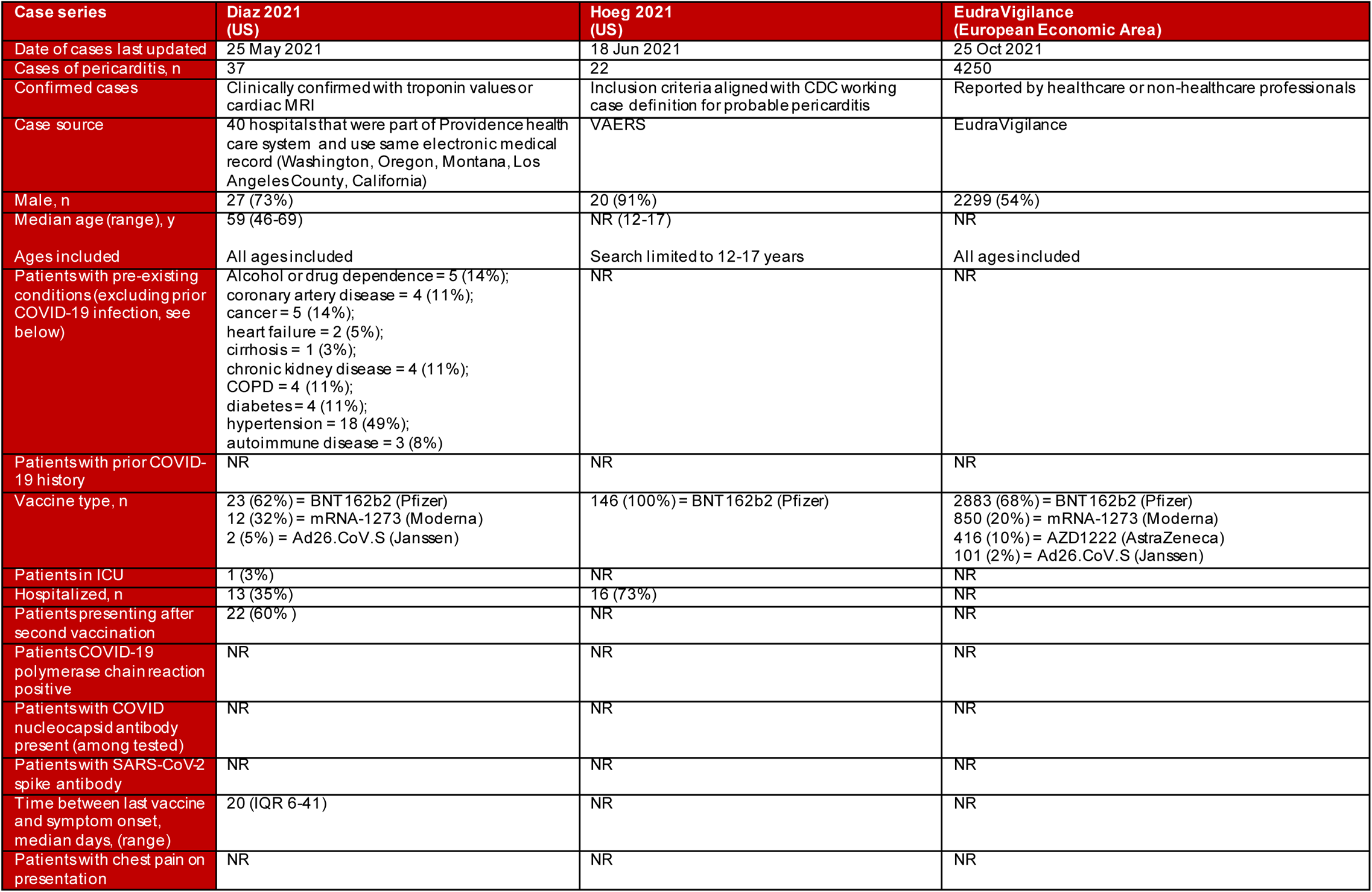
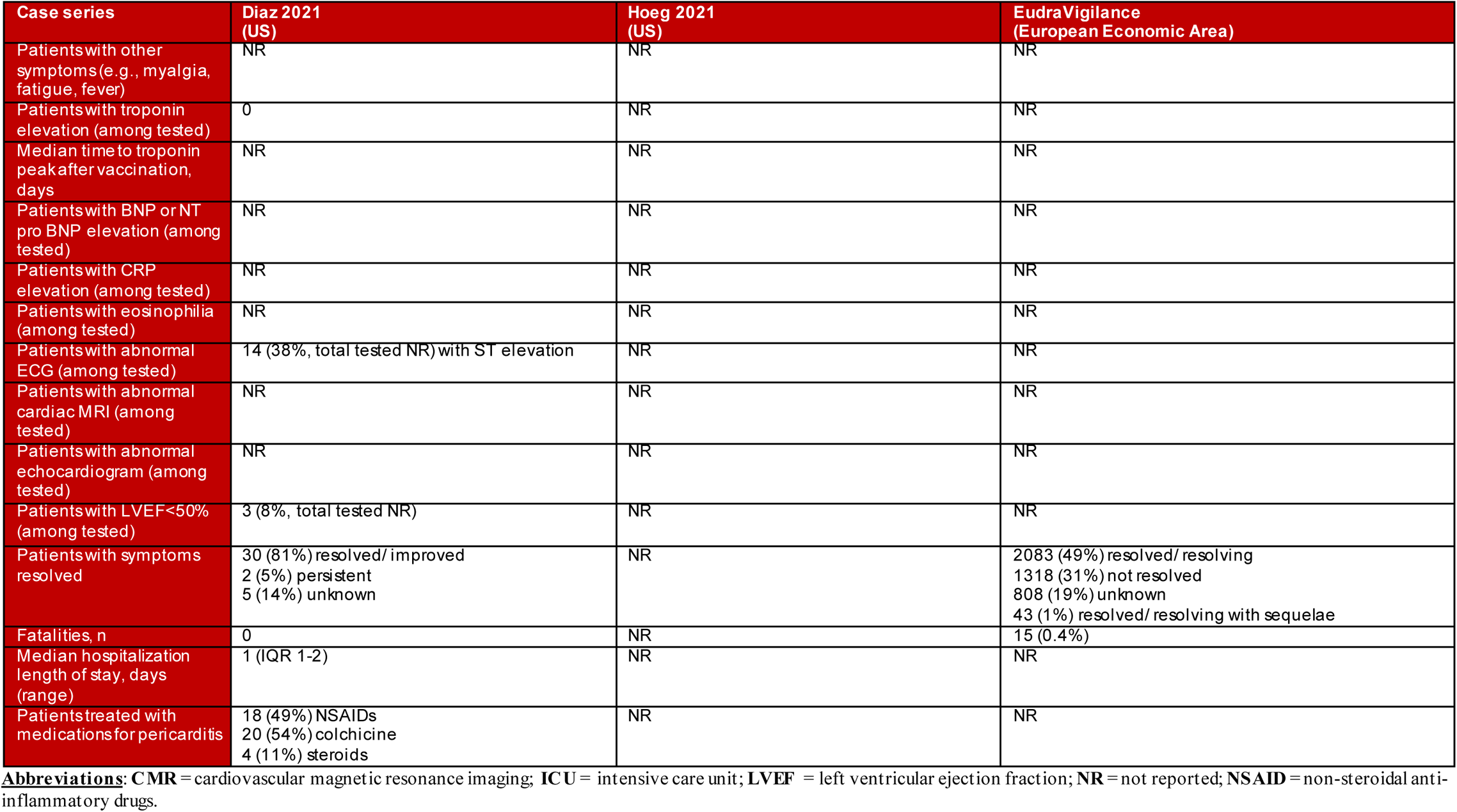
Results from Case Series with Short-term Follow-up of Pericarditis after COVID-19 Vaccination (Question 3b)

| Case series | Diaz 2021 (US) | Hoeg 2021 (US) | EudraVigilance (European Economic Area) |
| --- | --- | --- | --- |
| Date of cases last updated | 25 May 2021 | 18 Jun 2021 | 25 Oct 2021 |
| Cases of pericarditis, n | 37 | 22 | 4250 |
| Confirmed cases | Clinically confirmed with troponin values or cardiac MRI | Inclusion criteria aligned with CDC working case definition for probable pericarditis | Reported by healthcare or non-healthcare professionals |
| Case source | 40 hospitals that were part of Providence health care system and use same electronic medical record (Washington, Oregon, Montana, Los Angeles County, California) | VAERS | EudraVigilance |
| Male, n | 27 (73%) | 20 (91%) | 2299 (54%) |
| Median age (range), y | 59 (46-69) | NR (12-17) | NR |
| Ages included | All ages included | Search limited to 12-17 years | All ages included |
| Patients with pre-existing conditions (excluding prior COVID-19 infection, see below) | Alcohol or drug dependence = 5 (14%);<br>coronary artery disease = 4 (11%);<br>cancer = 5 (14%);<br>heart failure = 2 (5%);<br>cirrhosis = 1 (3%);<br>chronic kidney disease = 4 (11%);<br>COPD = 4 (11%);<br>diabetes = 4 (11%);<br>hypertension = 18 (49%);<br>autoimmune disease = 3 (8%) | NR | NR |
| Patients with prior COVID-19 history | NR | NR | NR |
| Vaccine type, n | 23 (62%) = BNT 162b2 (Pfizer)<br>12 (32%) = mRNA-1273 (Moderna)<br>2 (5%) = Ad26.CoV.S (Janssen) | 146 (100%) = BNT 162b2 (Pfizer) | 2883 (68%) = BNT 162b2 (Pfizer)<br>850 (20%) = mRNA-1273 (Moderna)<br>416 (10%) = AZD1222 (AstraZeneca)<br>101 (2%) = Ad26.CoV.S (Janssen) |
| Patients in ICU | 1 (3%) | NR | NR |
| Hospitalized, n | 13 (35%) | 16 (73%) | NR |
| Patients presenting after second vaccination | 22 (60%) | NR | NR |
| Patients COVID-19 polymerase chain reaction positive | NR | NR | NR |
| Patients with COVID nucleocapsid antibody present (among tested) | NR | NR | NR |
| Patients with SARS-CoV-2 spike antibody | NR | NR | NR |
| Time between last vaccine and symptom onset, median days, (range) | 20 (IQR 6-41) | NR | NR |
| Patients with chest pain on presentation | NR | NR | NR |
| Patients with other symptoms (e.g., myalgia, fatigue, fever) | NR | NR | NR |
| Patients with troponin elevation (among tested) | 0 | NR | NR |
| Median time to troponin peak after vaccination, days | NR | NR | NR |
| Patients with BNP or NT pro BNP elevation (among tested) | NR | NR | NR |
| Patients with CRP elevation (among tested) | NR | NR | NR |
| Patients with eosinophilia (among tested) | NR | NR | NR |
| Patients with abnormal ECG (among tested) | 14 (38%, total tested NR) with ST elevation | NR | NR |
| Patients with abnormal cardiac MRI (among tested) | NR | NR | NR |
| Patients with abnormal echocardiogram (among tested) | NR | NR | NR |
| Patients with LVEF < 50% (among tested) | 3 (8%, total tested NR) | NR | NR |
| Patients with symptoms resolved | 30 (81%) resolved/improved<br>2 (5%) persistent<br>5 (14%) unknown | NR | 2083 (49%) resolved/resolving<br>1318 (31%) not resolved<br>808 (19%) unknown<br>43 (1%) resolved/resolving with sequelae |
| Fatalities, n | 0 | NR | 15 (0.4%) |
| Median hospitalization length of stay, days (range) | 1 (IQR 1-2) | NR | NR |
| Patients treated with medications for pericarditis | 18 (49%) NSAIDs<br>20 (54%) colchicine<br>4 (11%) steroids | NR | NR |

**Abbreviations:** CMR = cardiovascular magnetic resonance imaging; ICU = intensive care unit; LVEF = left ventricular ejection fraction; NR = not reported; NSAID = non-steroidal anti-inflammatory drugs.

Most case series focused on myocarditis (10 reports, n=5,955). Across the case series, the majority of cases (often >90%) involved males with reported median age most often between 20 and 29 years; confirmed cases ranged from 12 to 56 years. Race/ethnicity was not reported in any case series. Time between last vaccine and symptom onset was on average 2 to 4 days, and the majority (71-100% across reports) presented after a second dose. Most cases presented with chest pain or pressure, and troponin elevation; when tested a minority (14-29% across studies) showed left ventricular dysfunction (i.e., LVEF<50%).

The majority of myocarditis cases were hospitalized (≥84%) with few admitted to ICU; average length of hospital stay was 2-4 days in four series (N=47) reporting on this. NSAIDs were most often used as treatment; other interventions included bisoprolol, ramipril, colchicine, famotidine, steroids (for myocarditis) and intravenous immune globulin (for myopericarditis). Among the series of confirmed cases of myocarditis that reported on fatalities (N=220), one fatality was reported in Israel in a 22-year-old with fulminant myocarditis. The unconfirmed series from the EudraVigilance data reported 84 fatalities among 5,611 cases (1.5%), though cause of death was not confirmed.

Three reports provided data for pericarditis (n=4,309; > 98% unconfirmed). The majority involved males; however, there was variation across reports (54 to 91%). Median age (59 years) was only reported in one small series (n=37). The same series reported a median interval of 20 days between last vaccine and symptom onset, with 60% presenting after the second vaccination. Hospitalization varied (35% and 73% reported in two series with a total of 59 cases); one series (n=37) reported 3% admitted to ICU and 1 day for median length of stay. One series (n=37) reported 0 fatalities, and a larger series of unconfirmed cases (n=4,250) reported 15 deaths (0.4%).

#### Question 4. Longer-term outcomes

Three reports^39 49 50^ reported on 38 cases with follow-up approximately 90 days after diagnosis of myocarditis following vaccination with an mRNA COVID-19 vaccine (**Table 6**). Among 14 patients hospitalized for myocarditis after vaccination in the two smaller case series (n=14), patients were males aged 13-19 years and followed up for ∼90-105 days after diagnosis. In the case series of five patients,^49^ repeat cardiac MRI was undertaken in two patients, with both showing persistent but decreased late gadolinium enhancement similar to the distribution of the initial MRI but no new abnormalities. Further, three patients had self-resolving mild intermittent chest pain after discharge, one had recurrent chest pain after discontinuing the NSAID prescribed at discharge, and three had recurrent symptoms that prompted emergency department visits post-discharge. In the series of nine patients,^50^ none were on heart failure medication at 90 day follow-up. In the larger case series (n=43),^39^ among cases of myocarditis and/or pericarditis (n=2) with long-term follow-up (n=24, mean follow-up time 89 days), the majority were males aged 12 to 17 years. Nine out of 18 patients receiving ECG had abnormal findings; 2 out of 17 with an echocardiogram showed abnormalities. Few (8%) patients were on medications such as NSAIDs and colchicine after discharge, and 46% had no symptoms, medications, or exercise restrictions at follow-up.

**Table 6.**
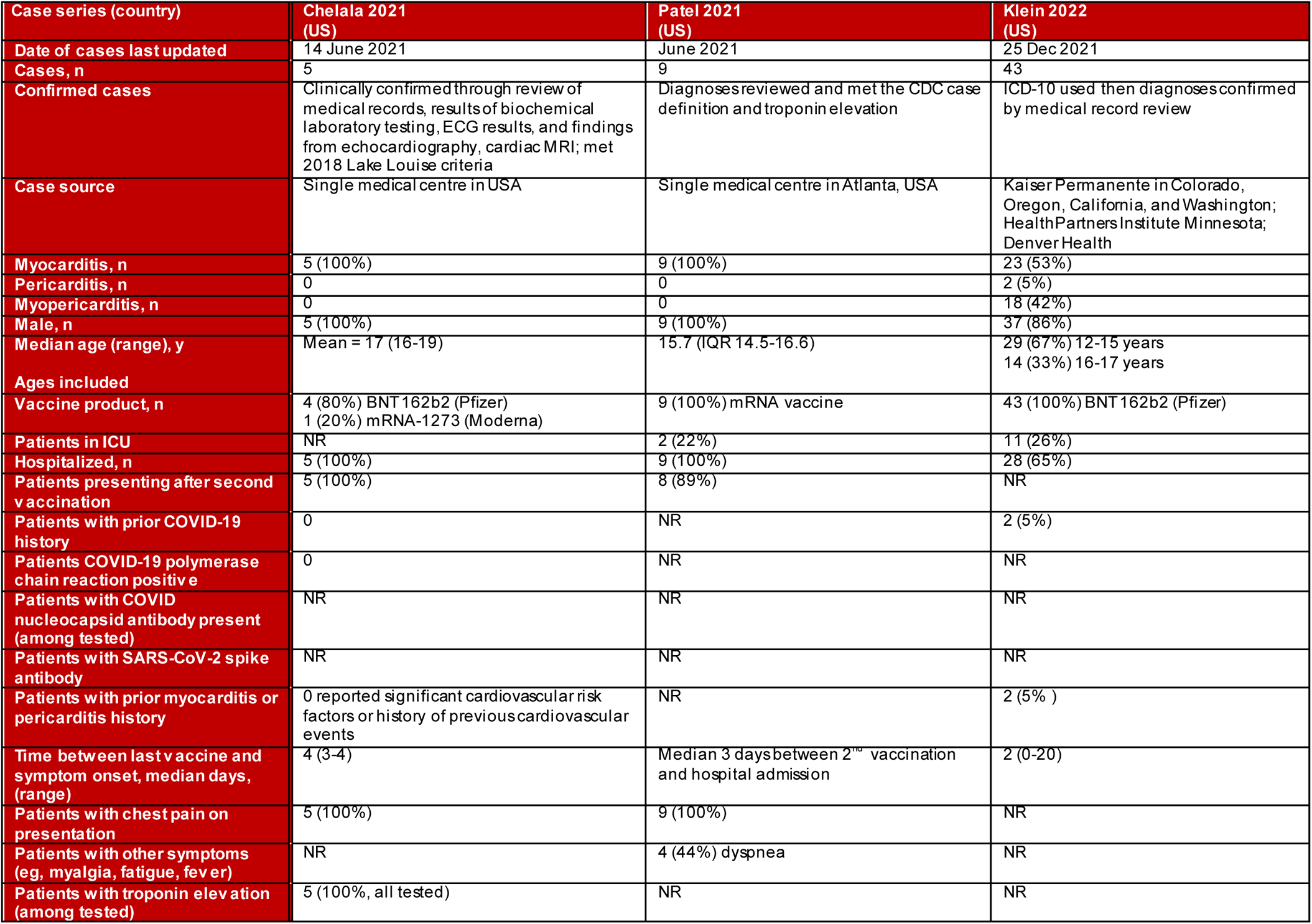
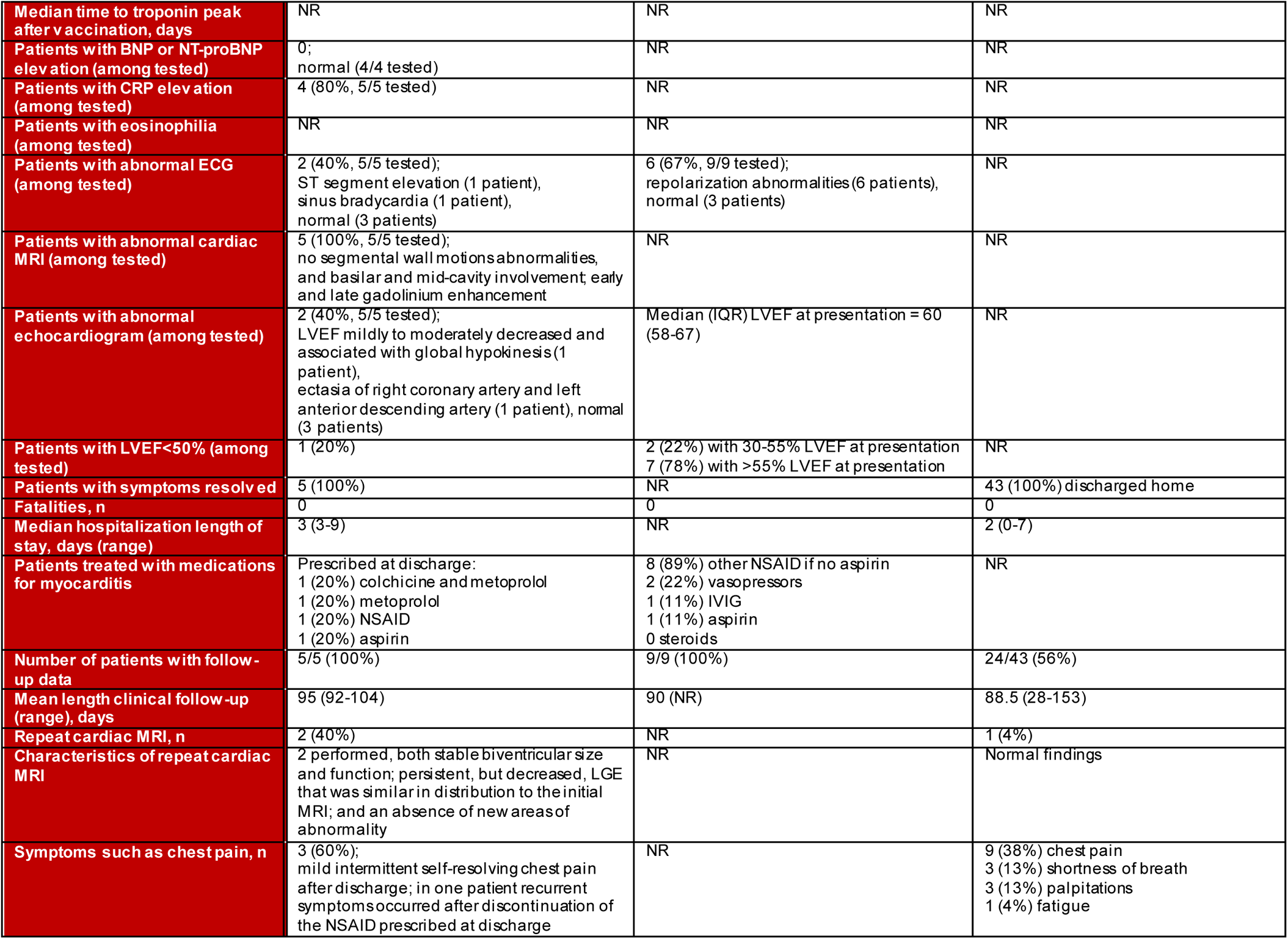
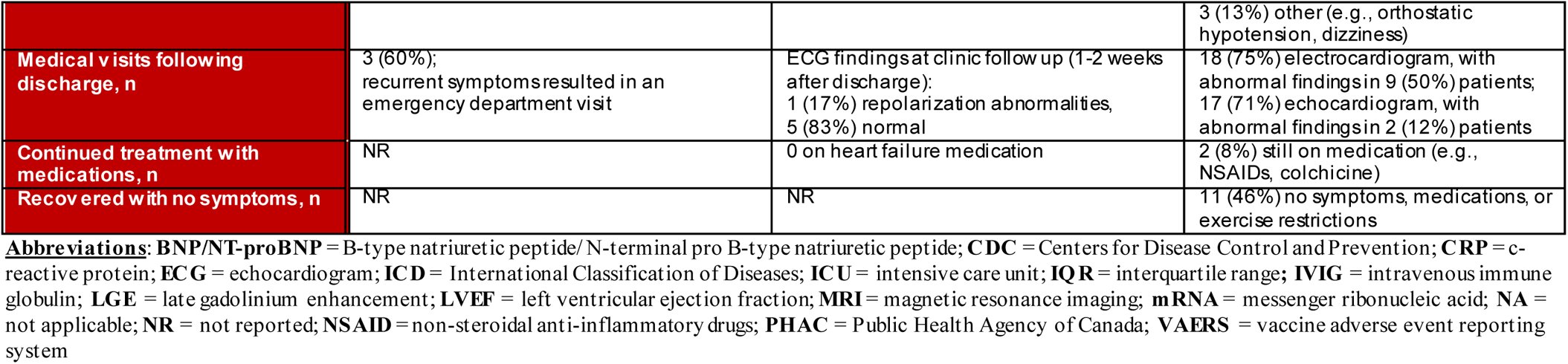
Case Series of Longer-term Follow-up of Myocarditis or Pericarditis after mRNA COVID-19 Vaccination (Question 4)

#### Question 5. Hypothesized Mechanisms

We included 21 papers,^9 51–70^ including narrative reviews, opinion pieces, letters to the editor, case reports, case series, a retrospective study, and a protocol for a prospective observational study. Across the included papers, we identified 16 hypotheses that are presented in **Table 7**. Additional details for each hypothesis are available in **Appendix 4**. All hypotheses related to myocarditis rather than pericarditis. The most commonly discussed hypotheses were: hyper immune/inflammatory response; autoimmunity triggered by molecular mimicry or other mechanism; delayed hypersensitivity (serum sickness); eosinophilic myocarditis; and hypersensitivity to vaccine vehicle components (e.g., polyethylene glycol [PEG] and tromethamine; lipid nanoparticle sheath). A number of novel hypotheses were put forward by single papers, such as low residual levels of double-strand RNA (dsRNA), hyperviscosity inducing cardiac problems, and strenuous exercise induced secretion of proinflammatory IL-6. A number of papers discussed observed differences in incidence by sex which could be attributed to sex steroid hormones or under-diagnosis in females.

**Table 7.**
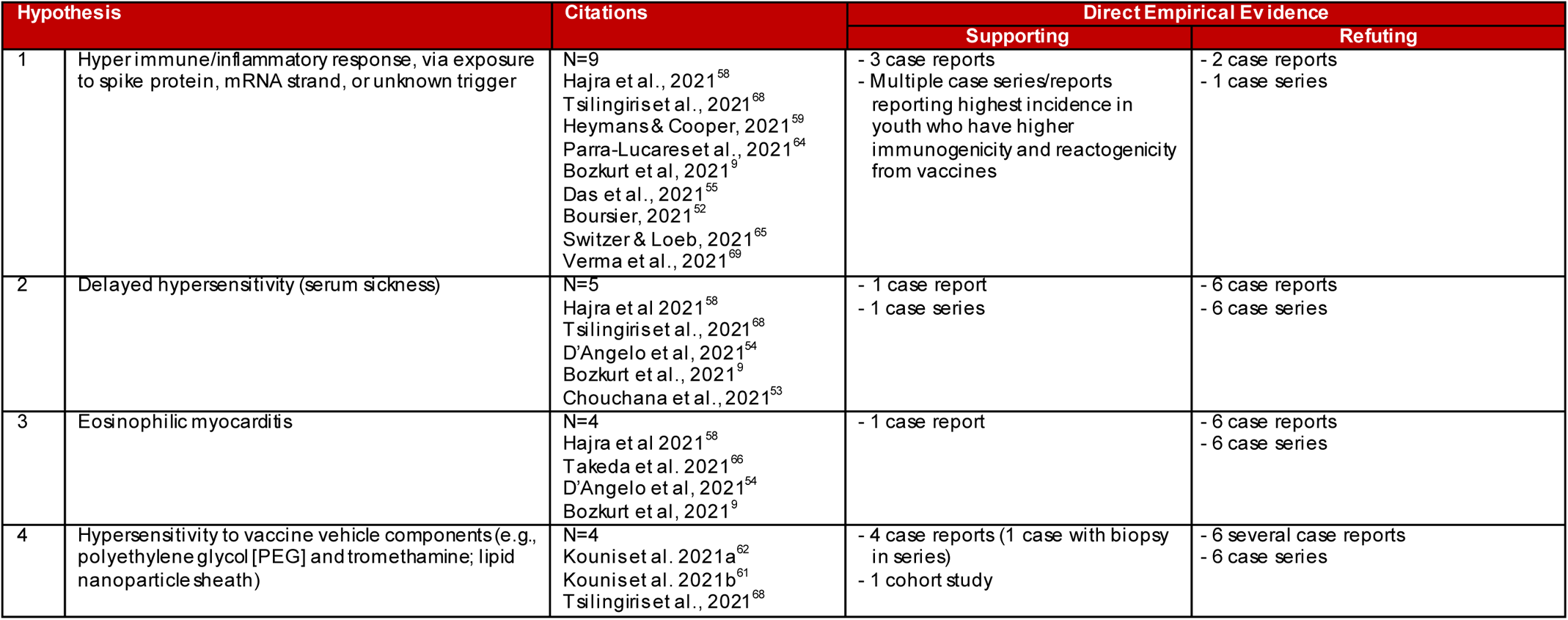
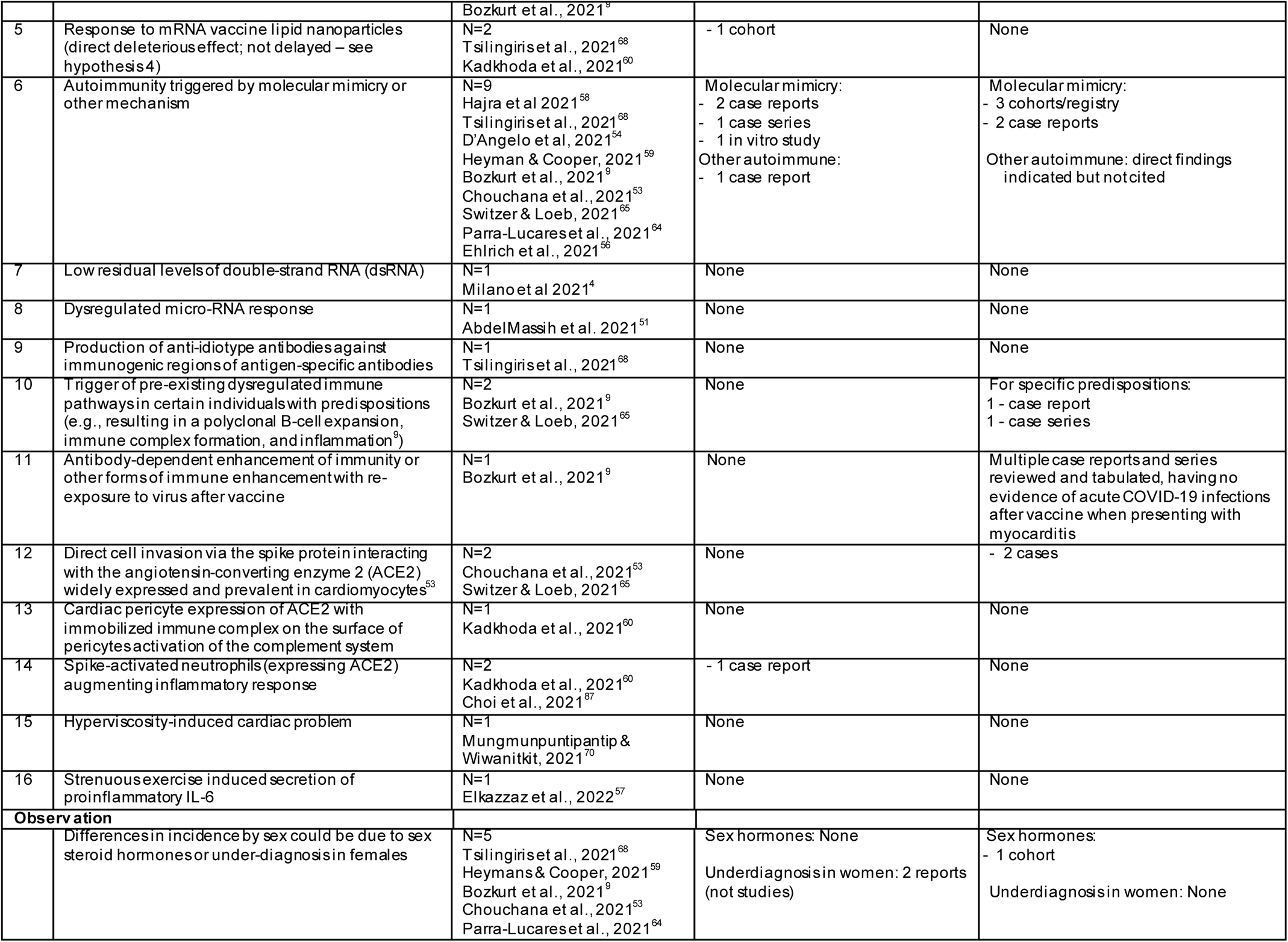
Hypothesized mechanisms for myocarditis following mRNA COVID-19 vaccination and direct (i.e., on myocarditis after COVID-19 vaccine) supporting/refuting empirical evidence (Question 5)

## Discussion

Adolescent and young adult males are at the highest risk of myocarditis after mRNA vaccination and the incidence may be as high as 140 cases per million (1.4 per 10,000). For 5-11 year-old males and females, the incidence of myocarditis after vaccination with Pfizer may be fewer than 20 cases per million although the evidence is of low certainty. Data on incidence rates after a third vaccine dose is limited. For 18-29 year-old males and females, incidence of myocarditis is probably higher after vaccination with Moderna compared to Pfizer (moderate certainty). Among 12-17, 18-29 and 18-39 year-olds, incidence of myocarditis/pericarditis after dose 2 of an mRNA vaccine may be lower when administered ≥31 days compared to ≤30 days after dose 1 (low certainty). Data specific to males aged 18-29 indicated that the dosing interval may have needed to increase to ≥56 days to substantially drop incidence. Data on other potential risk factors was very limited. The clinical course of myocarditis in children 5-11 years, for those after a third dose, and for those with previous myocarditis after mRNA vaccination is largely unknown. Though the short-term course has consistently demonstrated to be quite mild and self-limiting, more data on longer-term prognosis is needed.

Several mechanisms have been hypothesized to account for COVID-19 mRNA vaccine-associated myocarditis. The hyper immune/inflammatory response hypothesis raises the question of whether the response is a systemic process or local to the heart. Multi-organ injury is commonly seen in systemic inflammation, however, a cardiac insult is easier to detect due to chest pain symptoms and measurable changes in cardiac biomarkers and imaging. While autoimmunity triggered by molecular mimicry or other mechanism is among the more commonly discussed hypothesis, the observed response timing after the second vaccine dose (1-5 days) is considered early for this type of mechanism. If mRNA vaccine related myocarditis occurs from exposure to partial antigens (epitopes of SARs-CoV2 spike protein), then this mechanism should also account for myocarditis after COVID infection. Additionally, vaccines using adenoviral vector-based platforms produce the spike protein but have not been implicated in causing higher than background rates of myocarditis. The delayed hypersensitivity hypothesis is supported by earlier work of other viruses (e.g., coxsackieviruses, echoviruses). Eosinophilic myocarditis is unlikely a major cause of post-vaccination cardiac inflammation as the rate of myocarditis would be expected to approximate the rate of true allergic reactions to the vaccine. Hypersensitivity to vaccine vehicle components is a commonly discussed hypothesis; however, this is not likely to account for a major mechanism as allergic reactions have been very rare with the vaccines. The different incidence seen across sexes also suggests a non-allergic reaction predominating. The mechanism(s) may be very similar to that for myocarditis with COVID-19 infection, but at a lower incidence due to the smaller quantity of spike protein exposure. Therefore, exploring some of the mechanisms in the COVID-19 myocarditis literature may be valuable. One potential hypothesis that was not described in the examined articles relates to microvessel partial or complete thrombosis with multi-focal ischemic injury related to endothelial ACE2 expression and fibrin-platelet interactions in susceptible individuals.

Several limitations exist in the mechanistic literature we identified: i) little direct empiric evidence was available to support or refute the proposed hypotheses; where direct empiric evidence was available, it often derived from case reports or small series, ii) when assessing laboratory findings in case reports/series/retrospective studies, it is not clear whether any differences seen (e.g., increases in NK cells, autoantibodies) reflect a causal pathological immune response or reactive adaptive responses to the myocardial inflammation, iii) the emergence of new studies refuting some of the proposed mechanisms; for example, articles stating no reports of eosinophilia, are out-dated due to reports finding evidence of this, iv) there has been a lack of invasive investigations (e.g., biopsy, tissue morphology, special studies to detect injury, immune activity, virus, etc.) given the typically mild course of the clinical conditions observed, and v) it is difficult to confirm a causal link; for example, an important proportion of cases observed or reported may not be vaccine-related and this will contribute to the heterogeneity of presentations, clinical characteristics, and resulting hypotheses. Choi et al.^87^ described a fatal case of myocarditis after mRNA vaccination and compared the case to another fatality reported by Verma et al.^69^ both of which had comprehensive clinicopathological analysis. The two cases were remarkably different, suggesting “that myocarditis after COVID-19 mRNA vaccination is heterogeneous, both clinically and histologically.”^87^ Moreover, this data also supports the concept that post-COVID-19 vaccination related myocarditis can arise from different mechanisms.

Several key messages were co-developed with our patient partners to reflect their perspectives. First, the highest risk for myocarditis after COVID-19 vaccination exists for male adolescents and young adults (12-29y) although it is likely still small and occurrences appear to be quite mild with full recovery. There is probably some benefit for this population to receive Pfizer rather than Moderna and there may be value for dosing to be prolonged to some degree. Second, clear and effective communication of the risks (rates of myocarditis and likely clinical course) and benefits from vaccination, and the availability of good alternatives (e.g., non-mRNA vaccines) will be critical for young males and their parents. Finally, more research on children and what personal risk factors (e.g., pre-existing conditions) may put someone at higher risk is urgently needed.

### Strengths and limitations of the review

There are several strengths of this review. A comprehensive, peer-reviewed search strategy was used and inclusion of grey literature captured very recent data. A second reviewer screened the most relevant (based on machine learning) citations, and verified all data and risk of bias assessments. GRADE assessments were based on team consensus including clinical experts. Patient partners reviewed the evidence and developed interpretations from the patient perspective. The main limitation is that in the era of COVID-19 the literature base is evolving with incredible rapidity and new evidence will emerge; nevertheless, there was some moderate certainty evidence found in this review. We are unaware of any other comprehensive examination, and critique, of existing descriptions of the potential mechanisms that may be involved in COVID-19 vaccine-associated myocarditis. We may have missed some mechanistic studies because of the addition of this question for this update, after the original review was completed.

## Conclusions

Adolescent and young adult males are at the highest risk of myocarditis after an mRNA vaccination. Pfizer over Moderna and waiting more than 30 days between doses may be preferred for this population. As the incidence of myocarditis after mRNA vaccination remains a rare adverse event, the findings must be considered alongside the overall benefits of vaccination and with detailed risk-benefit analyses to support policy recommendations for optimal dosing intervals and vaccine products for different populations. As the COVID-19 pandemic enters its third year, continued surveillance of myocarditis after mRNA vaccines, especially in younger ages, after dose 3 (and subsequent doses) and in previous cases is needed to support continued decision making for COVID-19 boosters. Additional monitoring of populations with clinical comorbidities of interest (e.g., cardiac conditions, previous history of myocarditis, immunocompromised, etc.) is also needed in order to protect the already medically vulnerable. Long term follow-up of myocarditis cases is needed to better understand the natural history including disease recurrence. Finally, multi-center prospective studies with appropriate testing (e.g., biopsy, tissue morphology) will enhance understanding of the mechanism(s) of myocarditis post-vaccination which in turn will help identify and guide recommendations for those who may be at higher risk.

### Contributors

JP, LH, AM and IP designed the study. JP, LH, LB, LG, and AW screened citations for inclusion and were involved in data extraction and interpretation. JP, LB, LG and AW were involved is risk of bias assessments. All authors were involved in interpretation of the data. JP wrote the draft manuscript with input from all authors. All authors approved the final version of the manuscript. LH and JP are the guarantors of this manuscript and accept full responsibility for the work and the conduct of the study, had access to the data, and controlled the decision to publish. The corresponding author attests that all listed authors meet authorship criteria and that no others meeting the criteria have been omitted.

## Data Availability

All data produced in the present work are contained in the manuscript and appendices

## Acknowledgements

The authors thank Dr Bruce McManus for his insight on the proposed mechanisms and the patient/public partners Linda Wilhelm (New Brunswick) and Natasha Trehan (Toronto, Ontario) for contributing key message from their perspective; Susanna Ogunnaike-Cooke and Natalia Abraham (Knowledge Users; Public Health Agency of Canada) and Dr. Andrea Tricco (Unity Health Toronto, Ontario and SPOR Evidence Alliance) for reviewing the protocol and draft report from which this article stems; and Becky Skidmore, MLS and Kaitryn Campbell, MLS, MSc (St. Joseph’s Healthcare Hamilton/McMaster University) for drafting and running the searches, and for peer reviewing the search strategy, respectively.

## Funding Acknowledgement(s) & Disclaimer

This project was funded by the Canadian Institutes of Health Research (CIHR) through the COVID-19 Evidence Network to support Decision-making (COVID-END) at McMaster University and the SPOR Evidence Alliance. The funders provided input on the design of the study, but did not take part in the collection, analysis, or interpretation of data. They reviewed a report generated from the findings but did not make decisions about submission of the article for publication.

This review was developed through the analysis, interpretation and synthesis of scientific research and/or systematic reviews published in peer-reviewed journals, institutional websites and other distribution channels. This document may not fully reflect all the scientific evidence available at the time this report was prepared. Other relevant scientific findings may have been reported since completion of this synthesis report.

SPOR Evidence Alliance, COVID-END and the project team at ARCHE make no warranty, express or implied, nor assume any legal liability or responsibility for the accuracy, completeness, or usefulness of any information, data, product, or process disclosed in this report. Conclusions drawn from, or actions undertaken on the basis of, information included in this report are the sole responsibility of the user.

Dr. Hartling is supported by a Canada Research Chair in Knowledge Synthesis and Translation, and is a Distinguished Researcher with the Stollery Science Lab supported by the Stollery Children’s Hospital Foundation.

## Competing interests

All authors have completed the ICMJE uniform disclosure form at www.icmje.org/disclosure-of-interest/ and declare support from the funders for the submitted work; no financial relationships with any organisations that might have an interest in the submitted work in the previous three years; no other relationships or activities that could appear to have influenced the submitted work.

## Ethics approval

This research did not involve human research participants and thus did not require ethics approval.

## Transparency declaration

The lead authors of this work, JP and LH, affirm that the manuscript is an honest, accurate, and transparent account of the study being reported; that no important aspects of the study have been omitted; and that any discrepancies from the study as originally planned in the protocol have been explained.

## Data sharing

All of the data extracted for this review are included in the manuscript and associated supplementary files.

## Notes

**Funding statement:** This project was funded by the Canadian Institutes of Health Research (CIHR) through the COVID-19 Evidence Network to support Decision-making (COVID-END) at McMaster University and the SPOR Evidence Alliance. The funders provided input on the design of the study, but did not take part in the collection, analysis, or interpretation of data. They reviewed a report generated from the findings but did not make decisions about submission of the article for publication. Dr. Hartling is supported by a Canada Research Chair in Knowledge Synthesis and Translation, and is a Distinguished Researcher with the Stollery Science Lab supported by the Stollery Children’s Hospital Foundation.

### Competing Interest Statement

The authors have declared no competing interest.

