## Appendices for "Myocarditis and Pericarditis following COVID-19 Vaccination: Evidence Syntheses on Incidence, Risk Factors, Natural History, and Hypothesized Mechanisms"

### Appendix 1. Search Strategies

Database: Ovid MEDLINE(R) and Epub Ahead of Print, In-Process, In-Data-Review & Other Non-Indexed Citations and Daily <1946 to January 07, 2022>

Search Strategy:

--------------------------------------------------------------------------------

1 COVID-19 Vaccines/ [COVID VACCINES PT 1] (7756)

2 COVID-19/ (130502)

3 SARS-CoV-2/ (103370)

4 Coronavirus/ (4880)

5 Betacoronavirus/ (33227)

6 Coronavirus Infections/ (45252)

7 (COVID-19 or COVID19).tw,kf. (188432)

8 ((coronavirus* or corona virus*) and (hubei or wuhan or beijing or shanghai)).tw,kf. (5712)

9 (wuhan adj5 virus*).tw,kf. (278)

10 (2019-nCoV or 19nCoV or 2019nCoV).tw,kf. (1921)

11 (nCoV or n-CoV or "CoV 2" or CoV2).tw,kf. (72984)

12 (SARS-CoV-2 or SARS-CoV2 or SARSCoV-2 or SARSCoV2 or SARS2 or SARS-2 or severe acute respiratory syndrome coronavirus 2).tw,kf. (74134)

13 (2019-novel CoV or Sars-coronavirus2 or Sars-coronavirus-2 or SARS-like coronavirus* or ((novel or new or nouveau) adj2 (CoV or nCoV or covid or coronavirus* or corona virus or Pandemi*2)) or (coronavirus* and pneumonia)).tw,kf. (22369)

14 (novel coronavirus* or novel corona virus* or novel CoV).tw,kf. (11162)

15 ((coronavirus* or corona virus*) adj2 "2019").tw,kf. (41834)

16 ((coronavirus* or corona virus*) adj2 "19").tw,kf. (6655)

17 ("coronavirus 2" or "corona virus 2").tw,kf. (22546)

18 (OC43 or NL63 or 229E or HKU1 or HCoV* or Sars-coronavirus*).tw,kf. (4050)

19 COVID-19.rx,px,ox. or severe acute respiratory syndrome coronavirus 2.os. (6179)

20 (coronavirus* or corona virus*).ti. (25521)

21 COVID.ti. (146315)

22 ("B.1.1.7" or "B.1.351" or "B.1.617" or "B.1.427" or "B.1.429" or **"B.1.617.2"**).tw,kf,rx,px,ox. (1260)

23 ("P.1" and (Brazil* or variant?)).tw,kf,rx,px,ox. (1715)

24 (((alpha or beta or delta or eta or gamma or iota or kappa or lambda or **omicron**) adj3 variant?) and (coronavirus* or corona virus* or covid*)).tw,kf. (835)

25 or/2-24 [COVID-19] (228139)

26 exp Vaccination/ (95585)

27 ((COVID or COVID-19 or COVID19) adj5 (immunis* or immuniz* or inoculat* or vaccin*)).tw,kf. (11305)

28 ((coronavirus* or corona virus*) adj5 (immunis* or immuniz* or inoculat* or vaccin*)).tw,kf. (2060)

29 ((2019-nCoV or nCoV or n-CoV or SARS-CoV-2 or SARS-CoV2 or SARSCoV-2 or SARSCoV2 or SARS2 or SARS-2 or OC43 or NL63 or 229E or HKU1 or HCoV*) adj5 (immunis* or immuniz* or inoculat* or vaccin*)).tw,kf. (5651)

30 (BNT162 or BNT162-01 or BNT162a1 or BNT162b1 or BNT162b2 or BNT162c2 or N38TVC63NU).tw,kf. (1352)

31 (mRNA-1273 or EPK39PL4R4).tw,kf. (451)

32 (mRNA adj5 (immunis* or immuniz* or inoculat* or vaccin*)).tw,kf. (3357)

33 (messenger RNA adj5 (immunis* or immuniz* or inoculat* or vaccin*)).tw,kf. (338)

34 CX-024414.tw,kf. (3)

35 (Moderna adj5 (immunis* or immuniz* or inoculat* or vaccin*)).tw,kf. (384)

36 Spikevax$2.tw,kf. (15)

37 ((Pfizer or Pfizer-BioNTech) adj5 (immunis* or immuniz* or inoculat* or vaccin*)).tw,kf. (957)

38 (Comirnaty$2 or Tozinameran$2).tw,kf. (149)

39 or/26-38 [**VACCINES - GENERAL, MRNA VACCINES**] (109244)

40 25 and 39 [COVID-VACCINES PT 2] (16740)

41 1 or 40 [COVID-VACCINES PTS 1-2] (17964)

42 COVID-19 Vaccines/ae [adverse events] (922)

43 Viral Vaccines/ae [adverse events] (1942)

44 Adverse Drug Reaction Reporting Systems/ (8285)

45 ((immunis* or immuniz* or inoculat* or vaccin*) **adj5** (adverse* or ADE or ADEs or ADR or ADRs or complication* or harm* or safe or safety or side effect? or undesirable effect? or undesirable consequence? or undesirable outcome? or unintended effect? or unintended consequence? or unintended outcome?)).tw,kf. (26149)

46 ((post-vaccin* or post-immuni#ation* or post-innoculat* or after vaccin* or after immuni#ation* or after innoculat* or (following adj3 vaccin*) or (following adj3 immuni#ation*) or (following adj3 innoculat*)) **adj5** (adverse* or ADE or ADEs or ADR or ADRs or complication* or harm* or safe or safety or side effect? or undesirable effect? or undesirable consequence? or undesirable outcome? or unintended effect? or unintended consequence? or unintended outcome?)).tw,kf. (2369)

47 ((post-vaccin* or post-immuni#ation* or post-innoculat* or after vaccin* or after immuni#ation* or after innoculat* or (following adj3 vaccin*) or (following adj3 immuni#ation*) or (following adj3 innoculat*)) **adj5** (complication? or consequenc* or effect? or event? or harm* or outcome?)).tw,kf. (2683)

48 (vaccine-associated or vaccine-induced or vaccine-related).tw,kf. (9832)

49 (immuni#ation-associated or immuni#ation-induced or immuni#ation-related).tw,kf. (880)

50 ((data or vaccin*) adj3 (monitor* or surveillance*)).tw,kf. (31797)

51 Pharmacovigilance/ (2711)

52 (pharmacovigilan* or pharmaco-vigilan*).tw,kf. (6265)

53 (Canada Vigilance or Eudravigilance* or FAERS or VAERS).tw,kf. (1100)

54 Product Surveillance, Postmarketing/ (7482)

55 ((postmarket* or post-market*) adj3 surveillance*).tw,kf. (3754)

56 "Clinical Trial, Phase IV".pt. (2241)

57 ((Phase 4 or Phase IV) adj3 (evaluation? or study or studies or trial?)).tw,kf. (1715)

58 Cardiomyopathies/ (30527)

59 (cardiomyopath* or cardio-myopath* or myocardiopath* or myo-cardiopath*).tw,kf. (81484)

60 Myocarditis/ (15714)

61 myocarditis.tw,kf. (18216)

62 ((myocard* or myo-card) adj3 inflam*).tw,kf. (4049)

63 carditis.tw,kf. (2006)

64 exp Pericarditis/ (12119)

65 pericarditis.tw,kf. (13080)

66 ((pericard* or peri-card*) adj3 inflam*).tw,kf. (427)

67 epicarditis.tw,kf. (115)

68 ((epicard* or epi-card*) adj3 inflam*).tw,kf. (105)

69 (myopericarditis or myo-pericarditis).tw,kf. (751)

70 ((myopericard* or myo-pericard*) adj inflam*).tw,kf. (4)

71 (pleuropericarditis or pleuro-pericarditis).tw,kf. (185)

72 ((pleuropericard* or pleuropericard*) adj3 inflam*).tw,kf. (3)

73 or/42-72 [AEs, MYOCARDITIS, PERICARDITIS] (222568)

74 41 and 73 [COVID-VACCINES - AEs, MYOCARDITIS, PERICARDITIS] (4119)

75 (news or newspaper article).pt. (227385)

76 74 not 75 [NEWS ITEMS REMOVED] (4071)

77 limit 76 to yr="2020-current" (3989)

78 (202010* or 202011* or 202012* or 2021* or 2022*).dt. (1979506)

79 77 and 78 [**RECORDS SINCE 1 OCT 2020**] (3814)

80 ((Adaptive Clinical Trial or "Clinical Trial, Phase I" or "Clinical Trial, Phase II") not ("Clinical Trial, Phase III" or "Controlled Clinical Trial" or "Randomized Controlled Trial" or "Equivalence Trial" or "Pragmatic Clinical Trial" or "Clinical Trial, Phase IV")).pt. (35736)

81 79 not 80 [**EARLY PHASE CLINICAL TRIALS ONLY REMOVED**] (3812)

82 case reports.pt. (2237656)

83 81 not 82 [**CASE REPORTS REMOVED**] (3342)

84 (202110* or 202111* or 202112* or 2022*).dt. (399469)

85 83 and 84 [**UPDATE PERIOD – OCT 2021-CURRENT**] (1083)

86 (comment or editorial or letter).pt. (2027605)

87 83 and 86 [**OPINION PIECES - ORIGINAL AND UPDATE PERIOD**] (317)

88 85 or 87 [**UPDATE RECORDS, PLUS RECOVERY OF OPINION PIECES ORIGINAL PERIOD**] (1324)

### Appendix 2. Study Characteristics and Risk of Bias

#### Study characteristics: active surveillance/registry studies (Question 1)

| Dataset<br>Dates<br>Country | Vaccines Studied | Sample Size;<br>Demographics;<br>Previous Covid-19 diagnoses | Study Group(s) | Outcome(s); Risk Interval;<br>Case Ascertainment | Analysis | Results |
|---|---|---|---|---|---|---|
| **Dose 3** | | | | | | |
| NIMS/NHS<br><br>Dec 1 2020 to Nov 15 2021<br><br>England<br><br>Patone 2021 (7268) | Pfizer-BioNTech<br><br>Moderna<br><br>Dose 1, 2 or 3<br><br>Interval between doses NR | 21,554,158 with at least one dose, aged ≥13 y<br><br>Previous COVID in 54.7% of total sample<br><br>People with history of myocarditis in previous 2 years excluded | Pfizer<br><br>Dose 3: n= 10,599,183<br><br>Moderna<br><br>Dose 3: n= 343,716 | Hospitalization due to myocarditis<br><br>Risk interval: 28 d after any dose<br><br>Cases identified by ICD-10 codes: I40, I400, I401, I408, I409, I41, I410-412, I418, I514 | Incidence rate ratio using self-controlled case series (SCCS) method, stratified by sex and age | **Excess events per 1 mil persons receiving dose 3 (95% CI)**<br>1-28d — Dose 1 — Dose 2 — Dose 3<br>*Pfizer*<br><u><40y</u><br>Female — NR — NR — NR<br>Male — 3 (1, 5) — 12 (10, 13) — 13 (7, 15)<br><u>≥40y</u><br>Female — NR — NR — NR<br>Male — NR — NR — 3 (2, 4)<br><br>*Moderna*<br><u><40 y</u><br>Female — NR — 8 (4, 9) — NR<br>Male — 12 (1, 17) — 101 (95, 104) —<br><u>≥40y</u><br>Female — NR — NR — NR<br>Male — NR — NR — NR |
| Mayo Clinic Enterprise<br><br>Dec 1 2020 to Oct 17 2021<br><br>United States<br><br>Niesen, 2021 (4913) | Pfizer-BioNTech (78%)<br>Dose 1 & 2 18-28 d apart, Dose 3 ≥28 d after 2nd<br><br>Moderna<br>Dose 1 & 2: 25-35 d apart; dose 3 ≥28 d after dose 2<br><br>Dose 3 | 47,999 receiving exactly 3 doses (78% Pfizer)<br><br>Female 56.1%<br>Mean age: Pfizer 64 y (SD 17); Moderna 65 y (SD 13)<br>Hispanic or Latino 2%; Not Hispanic or Latino 95%; Unknown 3%<br><br>Covid-19 diagnoses NR | Received 3 homologous doses<br><br>Mean time dose 1 to 2: 28.6 d<br><br>Mean time dose 2 to 3: 173.0 d | Myocarditis<br><br>Risk interval: 0-14 d after each dose<br><br>Cases identified via electronic health records using a BERT-based classification model; identified cases were manually reviewed and confirmed by two investigators | Cumulative incidence | Events: 1 in female >40 years old (Moderna; 1 d after dose 3)<br><br>Cumulative incidence: 0.00% (95% CI 0% to 0.01%)<br><br>5,047 recipients of three doses of BNT162b2 and 558 recipients of three doses of mRNA-1273 were under 40 years of age.<br><br>33,662 recipients of three doses of BNT162b2 (57% female) and 9,582 recipients of three doses of mRNA-1273 (51% female) were 40 years of age or older. |
| **Age 5+** | | | | | | |

| Dataset<br>Dates<br>Country | Vaccines Studied | Sample Size; Demographics; Previous Covid-19 diagnoses | Study Group(s) | Outcome(s); Risk Interval; Case Ascertainment | Analysis | Results |
|---|---|---|---|---|---|---|
| VSD Dec 30<br><br>Thru Dec 30 2021<br><br>United States<br><br>Klein 2022 | Pfizer<br>Dose 1: 587,786<br>Dose 2: 556035 | Total doses: 1143821<br>5-11y: 431,485<br>12-15y: 750,772<br>16-17y: 393,049 | 1. Participants aged 5-11 y receiving at least 1 dose of Pfizer<br><br>2. Participants aged 12-17 y receiving at least 1 dose of Pfizer<br><br>2. Similar vaccinee in comparison interval (days 22-42) after COVID-19 vaccination. | Myocarditis, pericarditis, or myopericarditis<br><br>Risk interval: 21 d<br><br>Initial chart review followed with adjudication by an infectious disease clinician and/or a cardiologist to confirm cases meet CDC case definition | Excess cases based on comparison interval, adjusted for age group, sex, race/ethnicity, VSD site, and calendar date. | **5-11**<br>0 verified cases of myocarditis or myopericarditis<br>1 verified case of acute pericarditis in an 11 year-old.<br><br>**12-17**<br>12-15 years: 29 cases<br>16-17 years: 14 cases<br><br>43 validated cases among 12–17-year-olds, 0-21 days after vaccination<br>39 validated cases among 12–17-year-olds, 0-7 days after vaccination<br><br>Interval / Excess Cases per 1 million doses / 2-sided p-value<br>0-21 d<br>Dose 1 / 0.7 / 0.873<br>Dose 2 / 70.8 / <0.001<br><br>0-7 d<br>Dose 1 / 0.3 / 0.836<br>Dose 2 / 70.2 / <0.001 |
| **Age 12+** | | | | | | |
| DVR/DPR<br><br>Oct 1 2020 to Oct 5 2021<br><br>Denmark<br><br>Husby 2021 (4309) | Pfizer<br>n=3,482,295 (98% with 2 doses)<br>Median (IQR) 35 (24-36) days between dose 1 & dose 2<br><br>Moderna<br>n=498,814 (96.9% with 2 doses)<br>Median (IQR) 31 (28-35) days between dose 1 & dose 2 | 4,931,775 individuals contributing 4,717,464 person years<br><br>Pfizer<br>n=3,482,295<br>(98% with 2 doses)<br><br>Moderna<br>n=498,814<br>(96.9% with 2 doses)<br><br>Excluded individuals with a positive COVID-19 test result | Danish residents 12y or older and either<br>1) receiving Pfizer<br>2) receiving Moderna<br>3) Unvaccinated (14 d pre-vaccination period) | Myocarditis or myopericarditis<br><br>Risk interval: 28 d after any dose<br><br>Defined myocarditis or myopericarditis as a hospital diagnosis code of myocarditis or pericarditis (ICD-10 codes listed in table S1) and co-occurrence of elevated troponin levels, with a hospital stay >24 hours | 14 d pre-risk period before vaccination for each dose | **Absolute rate per 100,000 vaccinated persons (95% CI)**<br>Moderna / Pfizer<br>12-39y<br>Males, dose 2 / 9.80 (4.20, 22.84) / 1.54 (0.62, 3.81)<br><br>Other age/sex categories NR |
| **Age 16+** | | | | | | |

| Dataset<br>Dates<br>Country | Vaccines Studied | Sample Size;<br>Demographics;<br>Previous Covid-19 diagnoses | Study Group(s) | Outcome(s); Risk Interval; Case Ascertainment | Analysis | Results |
|---|---|---|---|---|---|---|
| Singapore Military<br><br>Jan 14 to Aug 3 2021<br><br>Singapore<br><br>Tan 2021 (4421) | Pfizer (37,367 individuals with 1+ dose)<br><br>Moderna (27,294 individuals with 1+ dose)<br><br>Homologous dose 2 administered between 21 and 56 days after dose 1 | 127,081 doses administered to 64,661 people (96.5% with 2 doses)<br><br>92.1% male<br><br>Previous or concurrent COVID-19 diagnosis NR | Singapore military personnel receiving at least 1 dose of an mRNA COVID-19 vaccine | Myocarditis<br><br>Risk interval: NR<br><br>Case ascertainment via military doctor or hospital diagnosis | Incidence rates and rate ratios after dose 2 versus dose 1 for both mRNA vaccines together and separately, with 95% confidence intervals | 3 events; all male, 18-21y, all after Moderna, none with cardiac history.<br><br>Reporting rate per 100,000 doses administered (95% CI)<br>*Any product* Dose 1 Dose 2<br><u>18-19 y</u><br>Female 0/955 0/903<br>Male 0/11,120 2/10,521<br><u>20-29 y</u><br>Female 0/2,819 0/2,717<br>Male 0/32,850 1/31,656<br><u>30-39y</u><br>Female 0/671 0/656<br>Male 0/7,807 0/7,625<br><br>Note: Only male data included in report; too few females for valid estimates |
| US Military Apr 30<br><br>Jan 1 to Apr 30 2021<br><br>United States<br><br>Montgomery 2021 | Pfizer-BioNTech or Moderna | 2,810,00 doses (38% dose 2)<br><br>Males 100%<br>Median age 25 (20-51)<br><br>Tested cases for Covid-19 n=0 but all cases after dose 2 (n=3) had previous Covid-19 | 1. Vaccinated<br>Expected numbers within 30 d after vaccination | Myocarditis<br><br>Cases identified via referrals to Defense Health Agency clinical specialists and through review<br>of VAERS reports; each cases adjudicated using CDC definition for probable<br><br>Risk interval: all presented within 4 d | Incidence in vaccinated<br><br>Observed vs expected cases: expected number based on an expected annual incidence ranging from 1-10 per 100 000 person-years (US) to 22 per 100 000 person-years (internationally); presenting within a 30-day period after vaccination. | Events: 23 (20 after dose 2)<br><br>**Observed vs expected:**<br>Total doses: 23 v vs 2 to 52<br>Dose 2: 20 vs 1 to 20<br>Dose 2 to military members: 19 vs 0 to 10<br>Dose 2 to male military members: 19 vs 0 to 8<br><br>**Incidence:**<br>Total doses: 0.8 per 100,000 doses<br>Dose 2: 1.9 per 100,000 doses<br>Dose 2 to military members: 3.5 per 100,000 doses<br>Dose 2 to male military members: 4.4 per 100,000 doses |

| Dataset Dates Country | Vaccines Studied | Sample Size; Demographics; Previous Covid-19 diagnoses | Study Group(s) | Outcome(s); Risk Interval; Case Ascertainment | Analysis | Results |
|---|---|---|---|---|---|---|
| Israel MOH May 31<br><br>Dec 20 2020 to May 31 2021<br><br>Israel<br><br>Mevorach 2021 | Pfizer-BioNTech<br><br>2 doses to 5.12 million people | Surveillance population 9,289,765 Israeli residents ≥16 y<br><br>29 of 98 cases in unvaccinated population had confirmed Covid-19 (timing NR); NR for vaccinated<br><br>Age, sex, race/ethnicity NR<br><br>None of the post-vaccination cases had concurrent Covid-19 via symptoms or PCR; 35/39 had negative serology tests | 1. Received dose 1 (n=5,442,696)<br>2. Received dose 2 (n=5,125,635)<br>3. Concurrent unvaccinated persons matched by date<br>4. Historical controls from 2017-2019<br><br>Interval between doses 21 d | Myocarditis/myopericarditis<br><br>ICD-9 codes 422.0-9x and 429.0x; cases 12-29 yrs old confirmed by medical record review using Brighton Criteria via consensus<br><br>Assessors not blinded to vaccine status<br><br>Cases of pericarditis without myocarditis were excluded<br><br>Risk interval: 21 d for dose 1 and 30d for dose 2 | Incidence in groups 1 and 2 (cumulative risks)<br>Risk difference (incidence) between 1 vs 2<br>Standardized incidence ratio of observed-to-expected (historical controls)<br>Attributable risk to dose 2<br>IRR group 2 vs 3 (weighted by age and sex)<br><br>Subgroups: sex, 5-yr age categories <30; 10 yr after 30; shorter risk interval 0-7 d | Total events in groups 1 and 2: 142 (136 definitive or probable [117 after dose 2], used for analysis except for standardized IR); group 3: 101<br><br>**Dose 2 incidence per 100,000 persons (0-30d risk interval)**<br>Males / Rate / aIRR (95% CI)<br>16-19 y / 15.07 / 8.96 (95% CI, 4.50 to 17.83)<br>20-24 y / 10.86 / 6.13 (95% CI 3.16 to 11.88)<br>25-29 y / 6.99 / 3.58 (95% CI 1.82 to 7.01)<br><br>Females<br>16-19 y / 1.00 / NE<br>20-24 y / 2.16 / 7.56 (95 CI, 1.47 to 8.96)<br>25-29 y / 0 / NE<br><br>**7 d risk interval (after dose 2):**<br>Males 16-19 y:<br>Risk difference: 13.62 per 100,000 persons (95% CI, 8.31 to 19.03)<br>IRR: 31.90 (95% CI, 15.88 to 64.08) |
| Israeli Defense Forces May 7<br><br>Dec 28 2020 to Mar 7 2021<br><br>Israel<br><br>Levin 2021 | Pfizer-BioNTech<br><br>138,000 military personnel receiving 2 doses | 138,000<br><br>NR<br><br>NR | 1. Vaccinated with 2 doses (n=138,000)<br><br>5. Interval between doses NR | Myocarditis<br><br>Medical record review, requiring ECG, echocardiography, or MRI findings<br><br>Risk interval: 7 d after dose 2<br><br>Not blinded | Crude cumulative incidence | Events: 7 confirmed in risk interval (100% male; Age 18-24)<br><br>Incidence: 5.07 per 100,000 people |

**Abbreviations**: DVR/DPR = Danish Vaccination Register & Danish Patient Register; NHS = National Health Service, which is the single-payer national health system in the UK; NIMS = NHS Immunisation Management Service database; VSD = Vaccine Safety Datalink.

### Study characteristics: passive surveillance/registry studies (Question 1)

| Dataset<br>Dates of data<br>Country of Data | Vaccines Studied | Outcome(s); Case Ascertainment & Risk Interval | Analysis | Results |
|---|---|---|---|---|
| **5y+** | | | | |

| Dataset<br>Dates of data<br>Country of Data | Vaccines Studied | Outcome(s); Case Ascertainment & Risk Interv al | Analysis | Results |
|---|---|---|---|---|
| VAERS Dec 19<br><br>Thru Dec 19 2021<br><br>USA<br><br>Su 2022 | Pfizer-BioNTech<br><br>Dose 1 or 2 (5-17 y)<br>5-11 y: n=8,674,37<br>12-17 y: n=18,707,169<br><br><br>Dose 3 (16-24 y)<br>16-17y: n= 47,040<br>18-24 y: n = 929,842 | Myocarditis<br><br>Risk interval: 7 d<br><br>Cases reported to VAERS confirmed using CDC working case definition | Reporting rate per million doses, compared to estimated background rate of 0.2 to 1.9 per 1 million person 7-day riskperiod | **Reporting rate of myocarditis per 1 million doses administered**<br>Dose 1 / Dose 2<br>5-11 y<br>Male 0.00 4.3*<br>Female NE 2.0<br>12-15 y<br>Male 4.8* 45.7*<br>Female 1.0 3.8*<br>16-17 y<br>Male 6.1* 70.2*<br>Female 0.00 7.6*<br><br>*Exceeds background incidence<br><br>**Reporting rate per 1 million Dose 3 in adolescents and young adults**<br>Events: 4 Verified cases; 2 among 16-17y, 2 among 18-24 y<br>16-17y: 2/47,040 = 42.5 per million doses<br>18-24y: 2/929,842 = 2.2 per million doses |
| VAERS Dec 9<br><br>Nov 2 to Dec 10 2021<br><br>USA<br><br>Su 2021a | Pfizer-BioNTech<br>7,141,428 doses<br>Dose 1: 5,126,642 (72%)<br>Dose 2: 2,014,786 (28%)<br><br>Dose interval NR | Myocarditis in 5-11yo<br><br>Risk interval: 0-12 d after any dose (VAERS)<br><br>Cases reported to VAERS confirmed using CDC working case definition | Reporting rate per million doses (estimated) | Events:<br>VAERS: 8 (50% female); 2 after dose 1, 6 after dose 2<br><br>**Crude reporting rate per 1 million doses administered**<br>Either dose: 8/7,141,428 = 1.12<br>Dose 1: 2/5,126,642 = 0.39<br>Dose 2: 6/2,014,786 = 2.98 |
| **12y+** | | | | |

<table>
<tr><th>Dataset<br>Dates of data<br>Country of Data</th><th>Vaccines Studied</th><th>Outcome(s); Case Ascertainment & Risk Interval</th><th>Analysis</th><th>Results</th></tr>
<tr><td>VAERS Oct 6<br><br>Up to Oct 6, 2021<br><br>USA<br><br>Su 2021b</td><td>Pfizer-BioNTech, Moderna and Janssen</td><td>Myopericarditis (myocarditis +/- pericardtis); pericarditis<br><br>Risk interval: 7 d<br><br>Screening via 30 MedDRA terms and ICD-10; all analyzed reports verified to meet CDC case definition by provider interview or medical record review</td><td>Crude reporting rates of confirmed cases of myopericarditis after each dose</td><td>For myopericarditis: 67% Pfizer; 29% Moderna; 76% after dose 2 (50 preliminary reports after Janssen not in analysis)<br><br>**Events: 935 verified cases; 797 in males; 138 in females.**<br><br>**Reporting rate of myopericarditis per 1 million doses administered**
<table>
<tr><th></th><th>Pfizer</th><th></th><th>Moderna</th><th></th></tr>
<tr><td>Males</td><td>Dose 1</td><td>Dose 2</td><td>Dose 1</td><td>Dose 2</td></tr>
<tr><td>12-15y</td><td>**4.2**</td><td>**39.9**</td><td>0.0</td><td>not calculated</td></tr>
<tr><td>16-17y</td><td>**5.7**</td><td>**69.1**</td><td>0.0</td><td>not calculated</td></tr>
<tr><td>18-24y</td><td>**2.3**</td><td>**36.8**</td><td>**6.1**</td><td>**38.5**</td></tr>
<tr><td>25-29y</td><td>1.3</td><td>**10.8**</td><td>**3.4**</td><td>**17.2**</td></tr>
<tr><td>30-39y</td><td>0.5</td><td>**5.2**</td><td>**2.3**</td><td>**6.7**</td></tr>
<tr><td>Females</td><td></td><td></td><td></td><td></td></tr>
<tr><td>12-15y</td><td>0.4</td><td>**3.9**</td><td>0.0</td><td>0.0</td></tr>
<tr><td>16-17y</td><td>0.0</td><td>**7.9**</td><td>0.0</td><td>0.0</td></tr>
<tr><td>18-24y</td><td>0.2</td><td>**2.5**</td><td>0.6</td><td>**5.3**</td></tr>
<tr><td>25-29y</td><td>0.2</td><td>1.2</td><td>0.4</td><td>**5.7**</td></tr>
<tr><td>30-39y</td><td>0.6</td><td>0.7</td><td>0.5</td><td>0.4</td></tr>
</table>
*Reporting rates exceed background incidence (bolded data above))<br><br>An estimated 1–10 cases of myocarditis per 100,000 person years occurs among people in the United States, regardless of vaccination status; adjusted for the 7-day risk period, this estimated background is 0.2 to 1.9 per 1 million person 7-day risk period</td></tr>
<tr><td>VAERS Jun 18a<br><br>Jan 1 to Jun 18 2021<br><br>USA<br><br>Høeg 2021</td><td>Pfizer-BioNTech<br><br>Moderna (only 1 of 257 cases; not approved for <18y)<br><br>Dose schedule NR</td><td>Myocarditis<br><br>“Myocarditis,” “pericarditis,” “myopericarditis” or “chest pain” in the symptom notes; “troponin” required element in the laboratory data; cases meeting CDC working case definition of probable myocarditis.<br><br>Risk interval: Any timing</td><td>Crude rates per million vaccinees<br><br>Cases with an unknown dose number were assigned to dose 1 or dose 2 in the same proportion as the known doses: 15% occurred following dose 1 and 85% occurred following dose 2</td><td>Events: 257 (92% within 5 d; 90% males)<br><br>**Crude reporting rate of myocarditis cases per million vaccinees**<br>Dose 2<br>Males 12-15 y: 162.2<br>Males 16-17 y: 94.0<br>Females 12-15 y: 13.0<br>Females 16-17 y: 13.4</td></tr>
</table>

| Dataset<br>Dates of data<br>Country of Data | Vaccines Studied | Outcome(s); Case Ascertainment & Risk Interval | Analysis | Results |
|---|---|---|---|---|
| Moderna Global Safety Database Sep 30<br><br>Dec 18 2020 to Sep 30 2021<br><br>Global<br><br>Strauss 2021 (4889) | Moderna (275,252,007 doses) | Myocarditis and/or myopericarditis<br><br>Risk interval: 7 d after any dose<br><br>Brighton Collaboration case definition and CDC working case definitions for acute myocarditis | Cumulative incidence of myocarditis/myopericarditis was assessed by calculating the reported rate after any known dose of mRNA-1273 according to age and sex, compared to population-based incidence (US Military) | **Reported Rate per 100,000 doses Within 7 Days**<br>Dose 1 / Dose 2 / Expected rate / RD<br><u><18 years</u><br>Females 0 / 0.15 / 1.74 / -1.59<br>Males 0.56 / 1.02 / 2.12 / -1.1<br><u>18-24y</u><br>Females 0.20 / 0.44 / 1.23 / -0.79<br>Males 1.15 / 4.91 / 2.12 / 2.79<br><u>25-39y</u><br>Females 0.09 / 0.19 / 1.23<br>Males 0.48 / 1.44 / 2.12 |
| COVaxON and Public Health Case and Contact Management Solution*<br><br>Jun 1 2020 to Sep 4 2021<br><br>Canada<br><br>Buchan 2021 (7142) | Moderna<br><br>Pfizer-BioNTech<br><br>Dose 1 or dose 2 (19,740,741 doses total) | Myocarditis<br><br>7-day risk interval<br><br>Group of specialized nurses and physicians classified cases according to Brighton Collaboration definition for myocarditis (level 1-2) | Crude rate per million doses, by dose | **Rate per million doses (95% CI), BC level 1-2 cases on or after Jun 1 2021**<br>*Pfizer* Dose 1 / Dose 2<br><u>12-17 y</u><br>Female 8.1 (1.0-29.1) / 9.7 (1.2-35.1)<br>Male 34.2 (15.6-64.9) / 88.1 (53.0-137.5)<br><u>18-24 y</u><br>Female 7.9 (0.2-44.1) / 0.0 (0.0-50.5)<br>Male 13.1 (1.6-47.3) / 35.5 (7.3-103.7)<br><u>25-39 y</u><br>Female 0.0 (0.0-14.3) / 13.1 (1.6-47.5)<br>Male 17.9 (5.8-41.8) / 12.6 (1.5-45.4)<br><br>*Moderna* Dose 1 / Dose 2<br><u>18-24 y</u><br>Female 0.0 (0.0-95.1) / 69.1 (14.2-201.9)<br>Male 0.0 (0.0-68.7) / 299.5 (171.2-486.4)<br><u>25-39 y</u><br>Female 0.0 (0.0 - 45.4) / 21.5 (2.6 - 77.7)<br>Male 28.8 (5.9-84.3) / 72.1 (31.1-142.0)<br><br>Note: Moderna not authorized for use in 12-17y in Canada |

COVaxON – Covid-19 Vaccinations Ontario is a central data repository for COVID-19 vaccine data and reporting in Ontario, administered by the Ontario Ministry of Health.

VAERS – Vaccine Adverse Events Reporting System. Passive surveillance system for the United States, to which healthcare providers and patients can report adverse events from medical products, including vaccines. Providers are required to report to VAERS adverse events (including administration errors, serious adverse events, cases of multisystem inflammatory syndrome, and cases of COVID-19 that result in hospitalization or death) that occur after receipt of any COVID-19 vaccine. Limitations include possible bias in reporting, inconsistent data quality, and incomplete information; in addition, VAERS has no direct comparison group. The VAERS system was not designed to assess causality; therefore, VAERS data generally cannot be used to determine whether a causal association between an adverse event and a vaccine exists

### Study characteristics of studies/reporting systems on risk factors (Question 2)

| Dataset<br>Dates of data *(mmm dd yyyy)*<br>Country of Data<br>Author year *(RefID)* | Vaccines Studied<br>Manufacturer<br>Dose # | Sample Size;<br>Demographics;<br>Previous Covid-19 diagnoses | Outcome(s)<br>Myo-, peri- and/or myopericarditis;<br>Case Ascertainment & Risk Interval;<br>Risk/protective factors considered | Outcome measures<br>**Analysis** *(e.g., adjustment for confounders)* | Results<br>Stratified by age and sex<br>If required, zero cell correction proportional to the reciprocal of the size of the contrasting study arm (i.e., # events = 1/n of the other arm) |
|---|---|---|---|---|---|
| **Risk factors related to vaccines** | | | | | |
| NIMS Nov 15<br>Dec 1 2020 to Nov 15 2021<br>England<br>Patone 2021 (7268) | Pfizer or Moderna<br>Pfizer<br>Dose 1 n=20,391,600;<br>Dose 2: n=17,294,004;<br>Dose 3: n= 10,599,183<br>Moderna<br>Dose 1 n=1,162,558;<br>Dose 2: n=1,039,919;<br>Dose 3: n= 343,716<br>Dosing scheduled NR | 21,554,158 with at least one dose, aged ≥13 y<br>Previous COVID in 54.7% of total sample.<br>People with history of myocarditis in previous 2 years excluded | Hospitalization due to myocarditis<br>28d risk interval<br>Cases identified by ICD-10 codes: I40, I400, I401, I408, I409, I41, I410-412, I418, I514<br>Pfizer vs. Moderna, by dose | Events per million doses<br>IRR calculated through self-control case series method<br>estimated crude ratio measures comparing Pfizer to Moderna by age group, by dose | **IRR (95% CI) - 0-28d**<br><u>≥40y</u> Dose 1 / Dose 2 / Dose 3<br>*Females*<br>Moderna 0 events / 0 events / 0 events<br>Pfizer 1.42 (0.96, 2.09) / 1.00 (0.64, 1.55) / 1.64 (0.91, 2.96)<br>*Males*<br>Moderna 0 events / 0 events / 0 events<br>Pfizer 0.97 (0.65, 1.47) / 0.79 (0.51, 1.23) / 2.48 (1.46, 4.19)<br>**IRR (95% CI) – 1-7d**<br><u>≥40y</u> Dose 1 / Dose 2 / Dose 3<br>*Females*<br>Moderna 0 events / 0 events / 0 events<br>Pfizer 1.40 (0.72, 2.74) / 0.80 (0.33, 1.97) / 2.32 (1.09, 4.94)<br>*Males*<br>Moderna 7.97 (3.17, 20.05) / 54.65 (29.74, 100.40) / NR<br>Pfizer 2.98 (1.75, 5.07) / 8.05 (5.37, 12.06) / NR |

| **Dataset**<br>**Dates of data** *(mmm dd yyyy)*<br>**Country of Data**<br>**Author year** *(RefID)* | **Vaccines Studied**<br>Manufacturer<br>Dose # | **Sample Size;**<br>**Demographics;**<br>**Previous Covid-19 diagnoses** | **Outcome(s)**<br>Myo-, peri- and/or myopericarditis;<br>Case Ascertainment & Risk Interval;<br>Risk/protective factors considered | **Outcome measures**<br>**Analysis** *(e.g., adjustment for confounders)* | **Results**<br>Stratified by age and sex<br>If required, zero cell correction proportional to the reciprocal of the size of the contrasting study arm (i.e., # events = 1/n of the other arm) |
|---|---|---|---|---|---|
| VSD Oct 9<br>Dec 14 2020 to Oct 9<br>USA<br>Klein, 2021 | Pfizer-BioNTech (60% of dose 2) or Moderna | Total doses 14,214,955 (71.5% eligible people fully vaccinated) to 7.5 million people<br>NR<br>Excluded from analysis if COVID-19 diagnosis ≤30 d before vaccination | Myocarditis/pericarditis/ myopericarditis (separate for head-to-head comparison)<br>Risk interval: 0-21 d & 0-7 d<br>Cases identified by ICD-10 codes; cases among 12-39 y confirmed by medical record review | Weekly, rapid cycle analysis<br>Adjusted incident rate ratio (aIRR) estimated by Poisson regression adjusted for age, sex, race and ethnicity, health plan, and calendar day; 1-sided p<0.0048 for statistical signal<br><u>Exploratory analyses</u> (no pre-specified level of significance): by dose and vaccine type; for 12-39 y by dose, vaccine type and with shorter risk intervals; head-to-head Moderna vs Pfizer in 18-39 y (also excluded pericarditis) | **Head-to-head Moderna vs Pfizer in 18-39 y (excluding pericarditis cases)**<br><u>0-7 d risk interval</u><br>aIRR / Excess cases per 1 million doses<br>Dose 2 (males): 2.14 (0.93 to 4.98) / 19.1 |

| **Dataset**<br>**Dates of data** *(mmm dd yyyy)*<br>**Country of Data**<br>**Author year** *(RefID)* | **Vaccines Studied**<br>Manufacturer<br>Dose # | **Sample Size;**<br>**Demographics;**<br>**Previous Covid-19 diagnoses** | **Outcome(s)**<br>Myo-, peri- and/or myopericarditis;<br>Case Ascertainment & Risk Interval;<br>Risk/protective factors considered | **Outcome measures**<br>**Analysis** *(e.g., adjustment for confounders)* | **Results**<br>Stratified by age and sex<br>If required, zero cell correction proportional to the reciprocal of the size of the contrasting study arm (i.e., # events = 1/n of the other arm) |
|---|---|---|---|---|---|
| VAERS* Oct 6<br>To Oct 6 2021<br>US<br>Su 2021 (7936) | Pfizer or Moderna<br>Dose 1 or Dose 2<br>Dosing interval NR | 366,062,239 doses of mRNA vaccine (either dose 1 or dose 2)<br>Doses NR by age/sex categories<br>Previous COVID-19 infection NR | Myocarditis<br>7 day risk period<br>Reports verified to meet case definition by provider interview or medical record review<br>Pfizer vs. Moderna | Reporting rate of myocarditis per 1 mil doses administered<br>Compared to background risk of 0.2 to 1.9 per 1 million person 7 day risk period<br>estimated crude Rate Ratios (for 18+ only; Moderna not authorized in <18y) | **Moderna vs. Pfizer** |

| | | Events per 1 mil doses | | crude Risk Ratio | |
|---|---|---|---|---|---|
| | | Dose 1 | Dose 2 | Dose 1 | Dose 2 |
| <u>18-24 y</u> | | | | | |
| Female | Moderna | 0.6 | 5.3* | 3.0 | 2.12 |
| | Pfizer | 0.2 | 2.5* | | |
| Male | Moderna | 6.1* | 38.5* | 2.65 | 1.05 |
| | Pfizer | 2.3* | 36.8* | | |
| <u>25-29 y</u> | | | | | |
| Female | Moderna | 0.4 | 5.7* | 2 | 4.75 |
| | Pfizer | 0.2 | 1.2 | | |
| Male | Moderna | 3.4* | 17.2* | 2.62 | 1.59 |
| | Pfizer | 1.3 | 10.8 | | |
| <u>30-39 y</u> | | | | | |
| Female | Moderna | 0.5 | 0.4 | 0.83 | 0.57 |
| | Pfizer | 0.6 | 0.7 | | |
| Male | Moderna | 2.3 | 6.7 | 4.6 | 1.29 |
| | Pfizer | 0.5 | 5.2 | | |
| <u>40-49 y</u> | | | | | |
| Female | Moderna | 0.2 | 1.4 | 2 | 1.27 |
| | Pfizer | 0.1 | 1.1 | | |
| Male | Moderna | 0.2 | 2.9 | 0.67 | 1.45 |
| | Pfizer | 0.3 | 2.0 | | |
| <u>50-64 y</u> | | | | | |
| Female | Moderna | 0.5 | 0.4 | 1.67 | 0.8 |
| | Pfizer | 0.3 | 0.5 | | |
| Male | Moderna | 0.5 | 0.6 | 2.5 | 2 |
| | Pfizer | 0.2 | 0.3 | | |
| <u>65y+</u> | | | | | |
| Female | Moderna | 0.0 | 0.3 | NE | 1.0 |
| | Pfizer | 0.1 | 0.3 | | |
| Male | Moderna | 0.1 | 0.3 | 0.5 | 3 |
| | Pfizer | 0.2 | 0.1 | | |

| **Dataset**<br>**Dates of data** *(mmm dd yyyy)*<br>**Country of Data**<br>**Author year** *(RefID)* | **Vaccines Studied**<br>Manufacturer<br>Dose # | **Sample Size;**<br>**Demographics;**<br>**Previous Covid-19 diagnoses** | **Outcome(s)**<br>Myo-, peri- and/or myopericarditis;<br>Case Ascertainment & Risk Interval;<br>Risk/protective factors considered | **Outcome measures**<br>**Analysis** (*e.g., adjustment for confounders)* | **Results**<br>Stratified by age and sex<br>If required, zero cell correction proportional to the reciprocal of the size of the contrasting study arm (i.e., # events = 1/n of the other arm) |
|---|---|---|---|---|---|
| Singapore Military Aug 3<br><br>Jan 14 to Aug 3 2021<br><br>Singapore<br><br>Tan 2021 (4421) | Pfizer<br>(37,367 individuals with 1+ dose)<br><br>Moderna<br>(27,294 individuals with 1+ dose)<br><br>Homologous dose 2 administered between 21 and 56 days after dose 1 | 127,081 doses administered to 64,661 military members (96.5% with 2 doses)<br><br>92.1% male<br><br>Previous or concurrent COVID-19 diagnosis NR | Myocarditis<br><br>Risk interval NR<br><br>Case ascertainment via military doctor or hospital diagnosis<br><br>Pfizer vs Moderna | Descriptive report only; crude numbers estimated by ARCHE | 3 events; all male, 18-21y, all after dose 2 of Moderna; 0 cases with history of cardiac conditions.<br><br>Overall rate: 2.4 per 100,000 doses<br><br>Dose 1 / Dose 2<br><u>18-20 y</u><br>**Pfizer**<br>Male 0/3,789 0/3,762<br>Female 0/326 0/323<br>**Moderna**<br>Male 0/7,331 2/6,759<br>Female 0/629 0/580<br><br><u>20-29 y</u><br>**Pfizer**<br>Male 0/18,278 0/18,203<br>Female 0/1,568 0/1,562<br>**Moderna**<br>Male 0/14,572 1/13,453<br>Female 0/1,251 0/1,155<br><br><u>30-39 y</u><br>**Pfizer**<br>Male 0/5,713 0/5,667<br>Female 0/491 0/487<br>**Moderna**<br>Male 0/2,094 0/1,958<br>Female 0/180 0/169 |

| Dataset<br>Dates of data *(mmm dd yyyy)*<br>Country of Data<br>Author year *(RefID)* | Vaccines Studied<br>Manufacturer<br>Dose # | Sample Size;<br>Demographics;<br>Previous Covid-19 diagnoses | Outcome(s)<br>Myo-, peri- and/or myopericarditis;<br>Case Ascertainment & Risk Interval;<br>Risk/protective factors considered | Outcome measures<br>**Analysis** *(e.g., adjustment for confounders)* | Results<br>Stratified by age and sex<br>If required, zero cell correction proportional to the reciprocal of the size of the contrasting study arm (i.e., # events = 1/n of the other arm) |
|---|---|---|---|---|---|
| COVaxON and Public Health Case and Contact Management Solution*<br><br>Dec 14 2020 to Sep 4 2021<br><br>Canada<br><br>Buchan 2021 (7142)<br><br>Buchan 2021 (7142) cont | Pfizer, Moderna<br><br>One or two doses<br><br>Dose interval NR | 19,740,741 doses<br><br>Demographics NR<br><br>History of COVID-19 NR | Myocarditis (product type)<br>Myocarditis/pericarditis<br><br>Group of specialized nurses and physicians classified cases according to Brighton Collaboration definition (level 1-3; myocarditis meeting level 1-2);<br><br>Risk interval: any time after vaccination (97.1% onset within 30 days).<br><br>Inter-dose interval ≤30 vs. ≥56 d;<br>Moderna dose 2 vs. Pfizer dose 2 | Rate ratios, unadjusted (for inter-dose interval) and adjusted for dose 1 product and interval (for dose 2, Moderna vs. Pfizer) | All ages and sexes:<br>**Inter-dose interval ≤30 vs. ≥56 d (ref)**, crude RR (95% CI):<br>Moderna: 5.2 (2.6-10.0)<br>Pfizer: 5.5 (3.1-9.6)<br>18-24 y, males: **Dose 2: Moderna vs. Pfizer (ref)**, adjusted RR (95% CI): 6.6 (3.3-13.2)<br><br>**Rate per million doses (95% CI), BC level 1-2 Myocarditis cases on or after 1 Jun 2021**<br>*Pfizer* — Dose 1 — Dose 2<br>12-17 y, male — 34.2 (15.6-64.9) — 88.1 (53.0-137.5)<br>12-17 y, female — 8.1 (1.0-29.1) — 9.7 (1.2-35.1)<br>18-24 y, male — 13.1 (1.6-47.3) — 35.5 (7.3-103.7)<br>18-24 y, female — 7.9 (0.2-44.1) — 0.0 (0.0-50.5)<br>25-39 y, male — 17.9 (5.8-41.8) — 12.6 (1.5-45.4)<br>25-39 y, female — 0.0 (0.0-14.3) — 13.1 (1.6-47.5)<br>≥40 y, male — 0.0 (0.0-14.4) — 0.0 (0.0-23.3)<br>≥40 y, female — 0.0 (0.0-14.8) — 0.0 (0.0 - 23.5)<br>*Moderna* — Dose 1 — Dose 2<br>12-17 y — NA — NA<br>18-24 y, male — 0.0 (0.0-68.7) — 299.5 (171.2-486.4)<br>18-24 y, female — 0.0 (0.0-95.1) — 69.1 (14.2-201.9)<br>25-39 y, male — 28.8 (5.9-84.3) — 72.1 (31.1-142.0)<br>25-39 y, female — 0.0 (0.0 - 45.4) — 21.5 (2.6 - 77.7)<br>≥40 y, male — 18.3 (2.2-66.2) — 0.0 (0.0-35.6)<br>≥40 y, female — 0.0 (0.0 - 40.5) — 0.0 (0.0 - 40.9)<br><br>**Rate per million (95% CI), by product (dose1-dose2)**<br>— Pfizer-Pfizer — Moderna-Pfizer<br>12-17 y — 53.8 (37.7-74.5) — NA |

| **Dataset**<br>**Dates of data** *(mmm dd yyyy)*<br>**Country of Data**<br>**Author year** *(RefID)* | **Vaccines Studied**<br>Manufacturer<br>Dose # | **Sample Size;**<br>**Demographics;**<br>**Previous Covid-19 diagnoses** | **Outcome(s)**<br>Myo-, peri- and/or myopericarditis;<br>Case Ascertainment & Risk Interval;<br>Risk/protective factors considered | **Outcome measures**<br>**Analysis** (*e.g., adjustment for confounders)* | **Results**<br>Stratified by age and sex<br>If required, zero cell correction proportional to the reciprocal of the size of the contrasting study arm (i.e., # events = 1/n of the other arm) |
|---|---|---|---|---|---|
| | | | | | 18-24 y 26.9 (14.3-45.9) 0.0 (0.0-218.8)<br>25-39 y 13.4 (7.5-22.1) 0.0 (0.0-107.0)<br>≥40 y 5.4 (3.1-8.6) 12.5 (0.3-69.7)<br><u>Moderna-Moderna</u> <u>Pfizer-Moderna</u><br>12-17 y NA NA<br>18-24 y 162.0 (108.5-232.6) 203.9 (142.0-283.6)<br>25-39 y 30.1 (16.0-51.4) 52.0 (32.2-79.5)<br>≥40 y 10.2 (4.7-19.4) 3.8 (0.8-11.0)<br><br>**Rate per million doses (95% CI), males 18-24 y, 2 doses by interval and product**<br>Events Doses Rate (95% CI)<br><u>Pfizer-Pfizer</u><br>Interval ≤30 d 2 21,160 94.5 (11.4-341.4)<br>Interval 31-55 d 8 124,235 64.4 (27.8-126.9)<br>Interval ≥56 d 1 90,424 11.1 (0.3-61.6)<br><u>Moderna-Moderna</u><br>Interval ≤30 d 4 10,623 376.5 (102.6-964.1)<br>Interval 31-55 d 20 60,352 331.4 (202.4-511.8)<br>Interval ≥56 d 3 22,641 132.5 (27.3-387.2)<br><u>Moderna-Pfizer</u><br>Interval ≤30 d 0 1,058 0.0 (0.0-3486.7)<br>Interval 31-55 d 0 5,402 0.0 (0.0-682.9)<br>Interval ≥56 d 0 2,393 0.0 (0.0-1541.5)<br><u>Pfizer-Moderna</u><br>Interval ≤30 d 6 7,720 777.2 (285.2-1691.6)<br>Interval 31-55 d 20 62,717 318.9 (194.8-492.5)<br>Interval ≥56 d 3 15,456 194.1 (40.0-567.2) |

| **Dataset**<br>**Dates of data** *(mmm dd yyyy)*<br>**Country of Data**<br>**Author year** *(RefID)* | **Vaccines Studied**<br>Manufacturer<br>Dose # | **Sample Size;**<br>**Demographics;**<br>**Previous Covid-19 diagnoses** | **Outcome(s)**<br>Myo-, peri- and/or myopericarditis;<br>Case Ascertainment & Risk Interval;<br>Risk/protective factors considered | **Outcome measures**<br>**Analysis** *(e.g., adjustment for confounders)* | **Results**<br>Stratified by age and sex<br>If required, zero cell correction proportional to the reciprocal of the size of the contrasting study arm (i.e., # events = 1/n of the other arm) |
|---|---|---|---|---|---|
| | | | | | **Rate per million doses (95% CI), dose 2 by product and interval**<br><u>Pfizer</u> — ≤30 d; 31-55 d; ≥56 d<br>12-17 y — 101.9 (55.7-170.9); 37.7 (21.6-61.3); 55.7 (20.4-121.2)<br>18-24 y — 45.3 (5.5-163.7); 34.7-15.9-66); 10.1 (1.2-36.5)<br>25-39 y — 42.5 (11.6-108.7); 8.7 (2.8-20.3); 12.3 (4.5-26.7)<br>≥40 y — 0.0 (0.0-34.4); 1.5 (0.0-8.3); 6.9 (4.0-11.1)<br><br><u>Moderna</u> — ≤30 d; 31-55 d; ≥56 d<br>12-17 y — NA; NA; NA<br>18-24 y — 353.1 (182.4-616.8); 184.0 (133.7-247.0); 103.2 (44.5-203.3)<br>25-39 y — 39.5 (8.1-115.4); 45.0 (29.1-66.4); 29.4 (10.8-64)<br>≥40 y — 0.0 (0.0-53.9); 7.4 (2.0-19.0); 7.5 (3.2-14.7) |
| **Other risk factors** | | | | | |
| EULAR COVAX*<br>Feb 5 to Jul 27 2021<br>Europe (30 countries)<br>Machado 2021 (3588) | Pfizer (n=3600)<br>Mean (SD) dose interval: 28 (12) days<br>Moderna (n=428)<br>Mean (SD) dose interval: 30 (8) days<br>74% with 2 doses; 1% with 3 doses | Reports of AEs in 4028 inflammatory (n=3218) or non-inflammatory (n=412) RMD patients.<br>70% female, mean age 61.6 (SD 15.2) years<br>History of COVID-19 NR | Myocarditis or pericarditis<br>Risk interval NR<br>Case ascertainment not reported<br>Inflammatory RMD vs. Non-inflammatory RMD | Crude ORs estimated from reported counts. | One event in a young (<30) female in I-RMD group with systemic lupus erythematosus after 2$^{nd}$ dose of Pfizer.<br>No events in NI-RMD group.<br><br>estimated OR<br>OR = (1/3599) / ((1/3600)/428)<br>OR = 428.1 |

| Dataset<br>Dates of data *(mmm dd yyyy)*<br>Country of Data<br>Author year *(RefID)* | Vaccines Studied<br>Manufacturer<br>Dose # | Sample Size;<br>Demographics;<br>Previous Covid-19 diagnoses | Outcome(s)<br>Myo-, peri- and/or myopericarditis;<br>Case Ascertainment & Risk Interval;<br>Risk/protective factors considered | Outcome measures<br>**Analysis** *(e.g., adjustment for confounders)* | Results<br>Stratified by age and sex<br>If required, zero cell correction proportional to the reciprocal of the size of the contrasting study arm (i.e., # events = 1/n of the other arm) |
|---|---|---|---|---|---|
| VAERS* Nov 30<br>Up to Nov 30 2021<br>Europe, US<br>Lane 2021 (7884) | Pfizer or Moderna<br>At least 1 dose<br>Dosing interval NR | 3066 VAERS reports of myocarditis or pericarditis<br>Demographics of total population not reported. The whole population had a bias of younger males experiencing myocarditis or pericarditis following COVID-19 mRNA vaccinations (20), whereas from the immunocompromised population 52.6% of these events occurred in males and 50.9% were under 60 years of age<br>Previous COVID-19 diagnosis NR | Myocarditis/pericarditis<br>Approximately 70% of reported events occurred within 14 days of vaccination<br>No case validation<br>Reports with comorbidities or concurrent medication indicative of transplantation, HIV infection, or cancer ("immunocompromised" population) were compared with each overall database population | Proportional reporting rates | 3066 cases, of which 57 (1.86%) were in immunocompromised individuals<br>PRR=1.36 [95% CI: 0.89-1.82] |

*Indicates passive surveillance system

COVaxON - Covid-19 Vaccinations Ontario is a central data repository for COVID-19 vaccine data and reporting in Ontario, administered by the Ontario Ministry of Health.

VSD - Vaccine Safety Datalink

EULAR COVAX- The European Alliance of Associations for Rheumatology Coronavirus Vaccine physician-reported registry. Data are entered voluntarily by rheumatologists or other members of the clinical rheumatology team; patients are eligible for inclusion if they registry have a pre-existing inflammatory/rheumatic and musculoskeletal disease or non-inflammatory rheumatic and musculoskeletal disease (NI-RMD) and have received one or more doses of any vaccine against SARS-CoV-2. Data are entered directly into an online data entry system or transferred from national registries (for Portugal). Patients with NI-RMDs are included as a control group.

VAERS – Vaccine Adverse Events Reporting System. Passive surveillance system for the United States, to which healthcare providers and patients can report adverse events from medical products, including vaccines. Providers are required to report to VAERS adverse events (including administration errors, serious adverse events, cases of multisystem inflammatory syndrome, and cases of COVID-19 that result in hospitalization or death) that occur after receipt of any COVID-19 vaccine. Limitations include possible bias in reporting, inconsistent data quality, and incomplete information; in addition, VAERS has no direct comparison group. The VAERS system was not designed to assess causality; therefore, VAERS data generally cannot be used to determine whether a causal association between an adverse event and a vaccine exists

### Risk of bias assessments for Question 1

| Dataset | Were the two groups similar and recruited from the same population? | Was vaccination status measured in a reliable or valid way? | Were all key confounding factors (age, sex, Covid-19 infection, pre-existing conditions) identified and appropriately addressed in design or analysis? | Were the outcomes measured in a valid and reliable way (medical record review)? | Was the follow up time long enough for outcomes to occur (7-30 days)? | Were the large majority of cases likely to have been identified? | Overall assessment of risk of bias |
|---|---|---|---|---|---|---|---|
| **Active surveillance studies** | | | | | | | |
| **Husby 2021** | Y | Y | U | U | Y | U | Some concerns |
| **Klein 2022** | Y | Y | U | Y | Y | Y | Some concerns |
| **Levin 2021** | NA | Y | N | Y | Y | Y | High |
| **Montgomery 2021** | NA | U | N (only for age and sex; potential confounders only reported for cases; no adjustment in analysis) | U | U | U | High |
| **Mevorach 2021** | Y | U | N (Considered age and sex, but not infection status) | Y | Y | Y | High |
| **Niesen 2021** | NA | Y | N (age [only < vs > 40; sex) | Y | Y | Y | High |
| **Patone 2021** | Y | Y | U | N | Y | U | High |
| **Tan 2021** | NA | Y | N (age and sex only) | U | U | N | High |

| | | | | | | | |
|---|---|---|---|---|---|---|---|
| **Passive surveillance studies** | | | | | | | |
| **Buchan 2021** | Y | Y | U | Y | N | Y | High |
| **Høeg 2021** | NA | N | N | U | U | N | High |
| **Strauss 2021** | NA | U | N | Y | U | U | High |
| **Su 2021a** | NA | Y | U | Y | Y | N | High |
| **Su 2021b** | NA | N | N | Y | Y | N | High |
| **Su 2022** | Y | Y | U | Y | Y | N | High |

**Risk of bias assessments for Question 2**

| **Dataset** | **Were the two groups similar and recruited from the same population?** | **Were the risk/protective factors measured similarly to assign individuals to exposed and unexposed groups?** | **Were the risk/protective factors measured in a valid and reliable way?** | **Were confounding factors identified and appropriately addressed in design or analysis (i.e. infection status, cardiac and immunodeficiency/autoimmune conditions)?** | **Were groups/ participants free of the outcome at the start of the study (or at time risk/protective factor was measured)?** | **Were the outcomes measured in a valid and reliable way?** | **Was the follow-up time long enough for outcome to occur?** | **Was follow-up complete, and if not, were reasons described and explored?** | **Overall assessment of risk of bias** |
|---|---|---|---|---|---|---|---|---|---|
| **Buchan 2021** | U | Y | Y | N | Y | Y | N | Y | High |
| **Klein 2021a** | Y | Y | Y | Y | Y | Y | Y | Y | Low |
| **Lane 2021** | Y | Y | N | N | U | N | U | N | High |

| | | | | | | | | | |
|---|---|---|---|---|---|---|---|---|---|
| **Machado 2021** | N | Y | Y | N | U | N | N | N | High |
| **Patone 2021** | N | Y | Y | Y | Y | N | Y | Y | High |
| **Su 2021** | U | Y | Y | N | Y | Y | Y | N | High |
| **Tan 2021** | Y | Y | Y | N | U | U | U | Y | High |

### Appendix 3.

#### Descriptions of passive surveillance systems used in studies within this report

EudraVigilance: Passive surveillance system for the European Economic Area. Healthcare providers and patients can report any adverse effects from medical products, including vaccines. Patients, consumers and healthcare professionals report suspected side effects to either the national medicines regulatory authority or the pharmaceutical company that holds the marketing authorisation for the medicine. These reports are then transmitted electronically to EudraVigilance.

VAERS: Passive surveillance system for the United States, to which healthcare providers and patients can report adverse events from medical products, including vaccines. Providers are required to report to VAERS adverse events (including administration errors, serious adverse events, cases of multisystem inflammatory syndrome, and cases of COVID-19 that result in hospitalization or death) that occur after receipt of any COVID-19 vaccine.

COVaxON - Covid-19 Vaccinations Ontario is a central data repository for COVID-19 vaccine data and reporting in Ontario, administered by the Ontario Ministry of Health.

EULAR COVAX- The European Alliance of Associations for Rheumatology Coronavirus Vaccine physician-reported registry. Data are entered voluntarily by rheumatologists or other members of the clinical rheumatology team; patients are eligible for inclusion if they registry have a pre-existing inflammatory/rheumatic and musculoskeletal disease or non-inflammatory rheumatic and musculoskeletal disease (NI-RMD) and have received one or more doses of any vaccine against SARS-CoV-2. Data are entered directly into an online data entry system or transferred from national registries (for Portugal). Patients with NI-RMDs are included as a control group.

#### References for included studies in questions 1 & 2 labeled based on data sources

| Study | Reference(s) |
|---|---|
| **Active Surveillance** | |
| Israel MOH May 31 | Mevorach D, Anis E, Cedar N, et al. Myocarditis after BNT162b2 mRNA Vaccine against Covid-19 in Israel. N Engl J Med. 2021. doi:https://dx.doi.org/10.1056/NEJMoa2109730. |
| Israeli Defense Forces May 7 | Levin D, Shimon G, Fadlon-Derai M, et al. Myocarditis following COVID-19 vaccination - A case series. Vaccine. 2021;39(42):6195-6200. doi:https://dx.doi.org/10.1016/j.vaccine.2021.09.004. |
| US Military Apr 30 | Montgomery J, Ryan M, Engler R, et al. Myocarditis Following Immunization With mRNA COVID-19 Vaccines in Members of the US Military. JAMA Cardiol. 2021;29:29. doi:https://dx.doi.org/10.1001/jamacardio.2021.2833. |
| VSD Dec 30 | Klein N. Vaccine Safety Datalink Rapid Cycle Analyses: Uptake and Safety of COVID-19 Vaccines in 5–11 and 12–17-Year-Olds. Presentation Jan 5, 2022 to U.S. Advisory Committee on Immunization Practices.; 2022. https://www.cdc.gov/vaccines/acip/meetings/downloads/slides-2022-01-05/04-COVID-Klein-508.pdf |
| VSD Oct 9 | Klein NP. Rapid Cycle Analysis to Monitor the Safety of COVID-19 Vaccines in Near Real-Time within the Vaccine Safety Datalink: Myocarditis and Anaphylaxis. Aug 30 Advisory Committee on Immunization Practices (ACIP) https://www.cdc.gov/vaccines/acip/meetings/downloads/slides-2021-08-30/04-COVID-Klein-508.pdf |
| DVR/DPR Oct 5 | Husby A, Hansen JV, Fosbol E, Thiesson EM, Madsen M, Thomsen RW, et al. SARS-CoV-2 vaccination and myocarditis or myopericarditis: population based cohort study. BMJ. 2021;375:e068665. |
| Singapore Military | Tan JTC, Tan C, Teoh J, Wahab MT, Tan GZ, Chin RYZ, et al. Adverse reactions and safety profile of the mRNA COVID-19 vaccines among Asian military personnel. Annals of the Academy of Medicine, Singapore. 2021;50(11):827-37. |
| NIMS/NHS Nov 15 | Patone M, Mei, W.X., Handunnetthi L., Dixon, S., Zaccardi, F., Shankar-Hari, M., et al. . Risk of myocarditis following sequential COVID-19 vaccinations by age and sex. medRxiv. 2021. https://www.medrxiv.org/content/medrxiv/early/2021/12/25/2021.12.23.21268276.full.pdf |
| Mayo Clinic Oct 17<br>US | Niesen MJM, Pawlowski C, O'Horo JC, Challener DW, Silvert E, Donadio G, et al. Three doses of COVID-19 mRNA vaccination are safe based on adverse events reported in electronic health records. medRxiv. 2021;09. https://dx.doi.org/10.1101/2021.11.05.21265961 |
| **Passive Surveillance** | |
| EudraVigilance Oct 19 | European Medicines Agency. EudraVigilance - European database of suspected adverse drug reaction reports. European Medicines Agency. Available from: https://www.adrreports.eu/en/eudravigilance.html. Published 2021. Accessed October 26, 2021, 2021. |

| Study | Reference(s) |
|---|---|
| VAERS Jun 18a | Høeg TB, Krug A, Stevenson J, Mandrola J. SARS-CoV-2 mRNA Vaccination-Associated Myocarditis in Children Ages 12-17: A Stratified National Database Analysis. 2021. doi:https://dx.doi.org/10.1101/2021.08.30.21262866. |
| VAERS Aug 6 | Lane S, Shakir S. Reports of myocarditis and pericarditis following mRNA COVID-19 vaccines: A review of spontaneously reported data from the UK, Europe, and the U. 2021. doi:https://dx.doi.org/10.1101/2021.09.09.21263342. |
| VAERS Oct 6 | Su JR. Myopericarditis following COVID-19 vaccination: Updates from the Vaccine Adverse Event Reporting System (VAERS). CDC October 21, 2021 2021. Available from: https://www.cdc.gov/vaccines/acip/meetings/downloads/slides-2021-10-20-21/07-COVID-Su-508.pdf. |
| VAERS Nov 30 | Lane S, Yeomans A, Shakir S. Systematic review of spontaneous reports of myocarditis and pericarditis in transplant recipients and immunocompromised patients following COVID-19 mRNA vaccination. 2021. https://medrxiv.org/cgi/content/short/2021.12.20.21268102 |
| VAERS Dec 9 | Su J. Adverse events among children ages 5–11 years after COVID-19 vaccination: updates from v-safe and the Vaccine Adverse Event Reporting System (VAERS). Presentation Dec 16 2022 to U.S. Advisory Committee on Immunization Practices. CDC Stacks; 2021. https://www.cdc.gov/vaccines/acip/meetings/downloads/slides-2021-12-16/05-COVID-Su-508.pdf |
| VAERS Dec 19 | Su JR. COVID-19 vaccine safety updates: Primary series in children and adolescents ages 5–11 and 12–15 years, and booster doses in adolescents ages 16–24 years. Presentation Jan 5, 2022 to U.S. Advisory Committee on Immunization Practices. 2022. https://www.cdc.gov/vaccines/acip/meetings/downloads/slides-2022-01-05/02-COVID-Su-508.pdf |
| COVaxON Sep 4 | Buchan SA, Seo CY, Johnson C, Alley S, Kwong JC, Nasreen S, et al. Epidemiology of myocarditis and pericarditis following mRNA vaccines in Ontario, Canada: by vaccine product, schedule and interval. medRxiv. 2021. https://www.medrxiv.org/content/medrxiv/early/2021/12/05/2021.12.02.21267156.full.pdf |
| EULAR COVAX | Machado PM, Lawson-Tovey S, Strangfeld A, Mateus EF, Hyrich KL, Gossec L, et al. Safety of vaccination against SARS-CoV-2 in people with rheumatic and musculoskeletal diseases: results from the EULAR Coronavirus Vaccine (COVAX) physician-reported registry. Annals of the Rheumatic Diseases. 2021;31:31. |
| Moderna Global Safety Database Sep 30 | Straus W, Urdaneta V, Esposito DB, Mansi JA, Rodriguez CS, Burton P, et al. Myocarditis after mRNA-1273 vaccination: A population-based analysis of 151 million vaccine recipients worldwide. medRxiv. 2021;12. https://dx.doi.org/10.1101/2021.11.11.21265536 |

### Appendix 4.

**Hypothesized mechanisms for myocarditis following COVID-19 vaccination and direct (myocarditis after COVID-19 vaccine) supporting/refuting empirical evidence* (Question 5)**

| Citation (citation type)<br><br>Specific aspect of hypothesis, as applicable | Main discussion points by authors, verbatim quotes and in-text citations | Direct empiric evidence supporting/refuting hypothesis (i.e., specific to COVID-19 vaccines)* |
|---|---|---|
| **Hypothesis 1: Hyper immune/inflammatory response** | | |
| Hajra et al., 2021[1] (narrative review)<br><br>Exposure to spike protein | • Children developed a more robust immune response than adults during SARS-CoV-2 infection, as demonstrated by multisystem inflammatory syndrome in children. In addition, mRNA vaccines produced more potent immunogenicity and reactogenicity in younger recipients and after the second dose. Similarly, the propensity of young adults to develop myocarditis following the second dose of vaccine supports the hypothesis of the vaccine-associated maladaptive immune response causing cardiac injury [35, 38, 45–47, 56, 58].<br>• Larson et al. [38] performed a cardiac biopsy in one patient before initiating steroids, and this did not demonstrate myocardial infiltrates.<br>• Muthukumar et al. [54] demonstrated an increase in a specific natural killer (NK) cell subset and multiple autoantibodies in a 52-year-old male with COVID-19 vaccine-associated myocarditis. In contrast, the interleukin (IL)-17 level was not raised, unlike other causes of myocarditis. The authors hypothesized that such unique immune changes might be contributing to a specific subtype of vaccine-associated myocarditis with rapid recovery.<br>• This systemic immune response, when exaggerated in predisposed individuals, might cause organ damage [59]. | Supporting:<br>Multiple case series/reports reporting on adolescents having higher incidence after second dose.<br>Muthukumar et al. In-depth evaluation of a case of presumed myocarditis after the second dose of COVID-19 mRNA vaccine. Circulation. 2021;144:487–98. Case report; increase in NK cells (lymphocytes)<br><br>Refuting:<br>Larson et al. Myocarditis after BNT162b2 and mRNA-1273 Vaccination. Circulation. 2021 ;144:506–508. Case report; no myocardial infiltrates.<br>Muthukumar et al. (see above). Case report; no IL-17 cytokine release (hence different cytokines possibly involved than with other types of myocarditis). |
| Tsilingiris et al., 2021[2] (article)<br><br>Exposure to mRNA strand | • mRNA strands are immunogenic and may themselves trigger an immune response directed against cardiomyocyte epitopes or adversely influence the myocardium in the frame of an exaggerated systemic reaction [22]. | None |
| Heymans & Cooper, 2021[3] (letter)<br><br>Exposure to mRNA strand | • The immune system might detect the mRNA in the vaccine as an antigen, resulting in the activation of proinflammatory cascades and immunological pathways in the heart. Although nucleoside modifications of mRNA reduce their innate immunogenicity, the immune response to mRNA might still drive the activation of an aberrant innate and acquired immune response, which can explain the stronger immune response seen with mRNA vaccines than with other types of COVID-19 vaccine. However, this hypothesis is not supported by the lack of immune-related adverse effects in other organs in which the mRNA vaccine is being uptaken. | None |
| Parra-Lucares et al., 2021[4] (case report and narrative review) | • This [mRNA] exogenous nucleotide material can be immunogenic and stimulate an innate immune response in organisms, generating an abnormal response with the potential to affect tissues other than the target cells of the therapy. To prevent this, nucleoside modifications are made to the mRNA used to decrease this unwanted immune response [55,59]. However, in patients with a genetic predisposition, it may not be sufficient to prevent it. The activation of | Supporting:<br>Muthukumar A et al. In-depth evaluation of a case of presumed myocarditis after the second dose of COVID-19 mRNA vaccine. Circulation. 2021;144:487–498. Case report (52 year-old) data with panel |

| | | |
|---|---|---|
| Exposure to mRNA strand | cells that express the Toll-like receptor and dendritic cells exposed to mRNA can activate pro-inflammatory cascades [59–61], which may have effects at the myocardial level.<br>• An exhaustive study of immunological mediators was conducted in one case [Muthukumar et al.]. Elevated plasma levels of interleukin-1 receptor (IL-1R) antagonist, interleukin 5 (IL-5), and interleukin 16 (IL-16) were observed, with no changes in interleukin 6 (IL-6), tumor necrosis factor (TNF), interleukin 1 beta (IL-1), interleukin 2 (IL-2), or interferon gamma (IFN). This patient also had increased plasma levels of natural killer (NK) cells, which destroy infected cells and participate in the innate immune response [65–67]. These preliminary data suggest a role for the abnormal activation of innate immunity in the development of vaccine-associated myocardial compromise. | of 48 cytokines and chemokines; elevated levels of some cytokines and NK cells. |
| Bozkurt et al, 2021[5]<br>(narrative review)<br><br>Exposure to mRNA strand or spike protein or unknown trigger | • Exposure to mRNA strand: The immune system may therefore detect the mRNA in the vaccine as an antigen, resulting in activation of proinflammatory cascades and immunologic pathways that may play a role in the development of myocarditis as part of a systemic reaction in certain individuals.<br>• Exposure to spike protein: By 1 case report, SARS-CoV-2 spike IgM and IgG neutralizing antibody levels were not significantly different in the patient with myocarditis than in individuals without myocarditis post–COVID-19 mRNA vaccination,[17](Mathukumar et al.) arguing against a hyperimmune response.<br>• Unspecified trigger: Surge in NK cells - Same patient had a 2-fold increase in the frequency of NK cells [17], which are the classical population of innate lymphoid cells, expressing a heterogeneous repertoire of germline encoded receptors that allows them to destroy cells that are infected by viruses, cancer cells, or cells that are rejected. The surge in NK cells may have either contributed to the pathology or the disease resolution process.<br>• Unspecified trigger: Dysregulated cytokine expression: (A) patient with myocarditis had elevated levels of IL-1 (interleukin 1) receptor antagonist, IL-5, IL-16, but not proinflammatory cytokines such as IL-6, tumor necrosis factor, IL-1B, IL-2, or interferon-γ levels. However, the patient had diminished levels of leukemia inhibitory factor, varying bidirectional profiles for IL-10, macrophage migration inhibitory factor, and vascular endothelial growth factor relative to an unvaccinated individual or a vaccinated individual without myocarditis.[17]<br>• *Bozkurt notes: It is not clear whether the differences seen in this patient regarding relative increases in NK cells, autoantibodies, and a dysregulated cytokine profile reflect a causal pathological immune response or reactive adaptive responses to myocardial inflammation* | <u>Supporting:</u><br>Unknown trigger, with surge in NK cells & dysregulated cytokine expression:<br>Muthukumar A et al. In-depth evaluation of a case of presumed myocarditis after the second dose of COVID-19 mRNA vaccine. Circulation. 2021;144:487–498. Case report data with panel of 48 cytokines and chemokines; elevated levels of some cytokines and NK cells<br><br><u>Refuting:</u><br>Exposure to spike protein:<br>Muthukumar A et al. In-depth evaluation of a case of presumed myocarditis after the second dose of COVID-19 mRNA vaccine. Circulation. 2021;144:487–498. Case report data; similar spike IgM and IgG neutralizing antibody levels |
| Das et al., 2021[6]<br>(case series)<br><br>Exposure to spike protein and other unknown trigger | • Exposure to spike protein: Anti-spike IgG antibody titers in a small subset of our patients were variable (data not shown) and did not correlate with the extent of cardiac injury.<br>• Exposure to unknown trigger: Furthermore, Muthukumar et al. conducted detailed immunologic investigation in a 52-year-old man who developed myocarditis 3 days after receiving the second dose of Moderna mRNA COVID-19 vaccine and reported that his antibody responses to 18 different SARS-CoV-2 antigens did not differ from (and were lower for some antigens) vaccinated controls who did not develop complications.[16] | <u>Refuting:</u><br>Exposure to spike protein or other unknown trigger, with antibody response:<br>Their case series data (n=25, 12-18 years)(Das)<br>Muthukumar A et al. In-depth evaluation of a case of presumed myocarditis after the second dose of COVID-19 mRNA vaccine. Circulation. 2021;144:487–498. Case report data; antibody responses to 18 different SARS-CoV-2 antigens same as controls. |
| Boursier et al.,, 2021[7]<br>(case reports)<br><br>Exposure to unknown trigger | • The DOTATOC-PET images showed an increase in myocardial uptake relative to blood activity, predominantly in the lateral and inferior walls. Myocardial/blood SUVmax ratio was >2.2 in both cases and, thus, higher than what we commonly observe in non-myocarditis patients. This likely reflects a myocardial infiltrate of inflammatory cells overexpressing somatostatin receptors (lymphocytes, macrophages, activated monocytes) [1–4], presumably within specific antigenic sites. | <u>Supporting</u>:<br>Two cases (18 and 21-year old males) with PET findings supporting myocardial infiltrate. |

| | | |
|---|---|---|
| Switzer & Loeb, 2021[8] (narrative review)<br><br>Exposure to unknown trigger | • A potential avenue for vaccine-associated myocarditis may be a nonspecific innate inflammatory immune response. | None |
| Verma et al., 2021[9] (letter to the editor describing 2 cases)<br><br>Exposure to unknown trigger | • Case 1: 45 year old women; endomyocardial biopsy specimen showed an inflammatory infiltrate predominantly composed of T-cells and macrophages, admixed with eosinophils, B cells, and plasma cells. Case 2: 42 year old man; autopsy revealed biventricular myocarditis...An inflammatory infiltrate admixed with macrophages, T-cells, eosinophils, and B cells was observed. | Supporting:<br>Biopsy and autopsy findings from their two cases; showing inflammatory infiltrate. |
| **Hypothesis 2: Delayed hypersensitivity (serum sickness)** | | |
| Hajra et al., 2021[1] (article) | • The development of symptoms within 1–4 days of the second dose of vaccine could be explained by a delayed hypersensitivity or serum sickness-like reaction. Additionally, patients who developed myocarditis following the first dose had a history of COVID-19 infection. In both cases, initial exposure caused sensitization to viral antigen with subsequent exposure forming antigen–antibody complexes and eventual damage to cardiac myocytes [33, 40, 55, 60]. | Supporting:<br>3 case series/reports reporting highest incidence after second dose, or history of previous COVID if experiencing myocarditis after first dose:<br>D'Angelo T et al. Myocarditis after SARS-CoV-2 vaccination: a vaccine-induced reaction? Can J Cardiol. 2021<br>Montgomery J et al. Myocarditis Following Immunization With mRNA COVID-19 Vaccines in Members of the US Military. JAMA Cardiol. 2021<br>Shay DK et al. Myocarditis Occurring After Immunization With mRNA-Based COVID-19 Vaccines. JAMA Cardiol [Internet]. 2021 [cited 2021 Sep 16] |
| Tsilingiris et al., 2021[2] (article) | • In the foreground stand immune or autoimmune mediated processes as possible mechanisms, and the highest frequency of occurrence after the second vaccine dose (after allowing for a presumed sensitization process to take place after the first dose) seems to strengthen this notion. | None |
| D'Angelo et al, 2021[10] (case report) | • In fact, the first vaccine dose may have presumably acquired sensitization. Moreover, the hypothesis of a delayed hypersensitivity after the second dose would be concordant either with the timing of symptoms, and with the mild peripheral eosinophilia seen in our case. | Supporting:<br>Case report data; 30 year-old male after second dose. |
| Bozkurt et al., 2021[5] (narrative review) | • Reports to date do not suggest a delayed hypersensitivity reaction, such as serum sickness–like reaction or eosinophilic myocarditis as a cause for myocarditis after mRNA COVID-19 vaccination.[15](D'Angelo et al.) Although rare, delayed localized skin hypersensitivity reactions have been described with mRNA COVID-19 vaccination with a median latency of 7 days,[59](Johnson et al.) unlike myocarditis emerging earlier within 3 to 4 days after vaccination. None of the case reports published to date had evidence of eosinophilia in peripheral blood or immune complex deposition or eosinophilic infiltrates in endomyocardial biopsy samples arguing against hypersensitivity, allergic or eosinophilic myocarditis.[8–17] | Refuting:<br>Several case reports and series; no eosinophilia:<br>Marshall M et al. Symptomatic acute myocarditis in seven adolescents following Pfizer-BioNTech COVID-19 vaccination. Pediatrics. Published online June 4, 2021.<br>Rosner CM et al. Myocarditis temporally associated with COVID-19 vaccination. Circulation. 2021;144:503–506. |

| | | |
|---|---|---|
| | | Abu Mouch S et al. Myocarditis following COVID-19 mRNA vaccination. Vaccine. 2021;39:3790–3793.<br>Larson KF et al. Myocarditis after BNT162b2 and mRNA-1273 vaccination. Circulation. 2021;144:507–509.<br>Ammirati E et al. Temporal relation between second dose BNT162b2 mRNA Covid-19 vaccine and cardiac involvement in a patient with previous SARS-COV-2 infection. Int J Cardiol Heart Vasc. 2021;34:100774.<br>Bautista GJ et al. Acute myocarditis after administration of the BNT162b2 vaccine against COVID-19. Rev Esp Cardiol (Engl Ed). Published online April 27, 2021;S1885-5857(21)00133-X.<br>Mclean K, Johnson T. Myopericarditis in a previously healthy adolescent male following COVID-19 vaccination: a case report. Acad Emerg Med. Published online June 16, 2021.<br>D'Angelo T et al. Myocarditis after SARS-CoV-2 vaccination: a vaccine induced reaction? Can J Cardiol. Published online June 9, 2021;S0828-282X(21)00286-5.<br>Albert E et al. Myocarditis following COVID-19 vaccination. Radiol Case Rep. 2021;16:2142–2145.<br>Muthukumar A et al. In-depth evaluation of a case of presumed myocarditis after the second dose of COVID-19 mRNA vaccine. Circulation. 2021;144:487–498.<br>Johnston MS et al. Delayed localized hypersensitivity reactions to the Moderna COVID-19 vaccine: a case series. JAMA Dermatol. 2021;157:716–720. Skin reactions rare and delayed more than myocarditis. |
| Chouchana et al., 2021[11] (retrospective study on Vigibase case and discussion) | • This may be related to greater adaptive immune response in younger individuals, which may lead to greater increases of CD4+ Th17+ cell populations, predisposing individuals to developing myocarditis. It would be interesting to see if the recently reported mRNA diagnostic of Th17 activation in myocarditis is also positive in these patients.[41] | None |
| **Hypothesis 3: Eosinophilic myocarditis** | | |
| Hajra et al 2021[1] (narrative review) | • Small pox vaccine and tetanus toxoid vaccine have been found to cause myocardial damage following immunization. Endomyocardial biopsy has demonstrated evidence of eosinophilic | None in this review; authors of cited reports [45, 58] did not examine eosinophilia. |

| | | |
|---|---|---|
| | myocarditis in such cases [62, 63]. Increased circulating eosinophils produced following immunization infiltrate cardiac tissue. Degranulation of eosinophils causes direct myocardial injury [64]. A similar mechanism might exist in the case of mRNA COVID-19 vaccine-associated myocarditis. However, the lack of peripheral eosinophilia in a few instances renders this mechanism unlikely [45, 58]. | |
| Takeda et al. 2021[12] (case report) | • Case report data: Interventricular septal biopsies obtained from the right ventricle revealed diffuse eosinophilic infiltration of the myocardial interstitium. Eosinophilic infiltration, as well as eosinophil degranulation between the myocardial fibers, was observed. | Supporting: Case report biopsy data, 53 year-old male; no data on whether from exposure to spike protein epitope. |
| D'Angelo et al, 2021[10] (case report and discussion) | • Case report data: White blood cells were 10.4 x $10^3$/μL (normal 4.0-10.0), with **mild** eosinophilia (0.9 x $10^3$/μL, normal 0.0-0.5 x $10^3$).<br>• A further hypothesis can be represented by eosinophilic myocarditis directly after immunisation, which has been reported as an extremely rare event, despite the possible underdiagnosis due to its delayed development.[5] | Refuting: Case report laboratory data (only mild eosinophilia), 30 year-old male; no data on whether from exposure to spike protein epitope. |
| Bozkurt et al, 2021[5] (narrative review) | • (In a case report and series (n=4), there was also no evidence of leukocytosis, eosinophilia, anemia, thrombocytopenia, or transaminase elevation.[19,12](Ammirarti et al. and Kim et al.)<br>• Reports to date do not suggest a delayed hypersensitivity reaction, such as serum sickness–like reaction or eosinophilic myocarditis as a cause for myocarditis after mRNA COVID-19 vaccination.[15](D'Angelo T et al.) Although rare, delayed localized skin hypersensitivity reactions have been described with mRNA COVID-19 vaccination with a median latency of 7 days,[59](Johnson et al.) unlike myocarditis emerging earlier within 3 to 4 days after vaccination. None of the case reports published to date had evidence of eosinophilia in peripheral blood or immune complex deposition or eosinophilic infiltrates in endomyocardial biopsy samples arguing against hypersensitivity, allergic or eosinophilic myocarditis.[8–17] | Refuting:<br>Ammirati E et al. Temporal relation between second dose BNT162b2 mRNA Covid-19 vaccine and cardiac involvement in a patient with previous SARS-COV-2 infection. Int J Cardiol Heart Vasc. 2021;34:100774. doi: 10.1016/j.ijcha.2021.100774: Case report with no eosinophilia<br>Kim HW et al. Patients with acute myocarditis following mRNA COVID-19 vaccination. JAMA Cardiol. Published online June 29, 2021. doi: 10.1001/jamacardio.2021.2828. Case series n=3 without eosinophilia<br>D'Angelo T et al. Myocarditis after SARS-CoV-2 vaccination: a vaccine induced reaction? Can J Cardiol. Published online June 9, 2021;S0828-282X(21)00286-5.<br>Johnston MS et al. Delayed localized hypersensitivity reactions to the Moderna COVID-19 vaccine: a case series. JAMA Dermatol. 2021;157:716–720. doi: 10.1001/jamadermatol.2021.1214. Skin reactions rare and delayed more than myocarditis.<br>Several case reports and series; no eosinophilia: (see Hypothesis 2) |
| **Hypothesis 4: Hypersensitivity to vaccine vehicle components (e.g., polyethylene glycol [PEG] and tromethamine; lipid nanoparticle sheath)** | | |

| | | |
|---|---|---|
| Kounis et al. 2021a[13] (letter)<br><br>Hypersensitivity to PEG and tromethamine | • Sokolska et al. described young patient [1] had an atopic diathesis due to his previous history of atopic asthma, pollen and pet allergy and, therefore, the induced myocarditis was presumably hypersensitivity myocarditis.<br>• In 2 cases of myocarditis following COVID-19 vaccination in the USA and in 1 in Israel, the endomyocardial biopsies revealed eosinophils and other interacting and interrelated inflammatory cells such as macrophages, T-cells, and B cells compatible with hypersensitivity myocarditis [2](Witberg et al.)<br>• This type of myocarditis is particularly difficult to recognise because the clinical features characteristic of a drug hypersensitivity reaction — including non-specific skin rash, malaise, fever, and eosinophilia — are absent in most cases [not specific to COVID vaccine cases] [3]. | Supporting:<br>Sokolska JM et al. Every rose has its thorns — acute myocarditis following COVID-19 vaccination. Kardiol Pol. 2021; 79(10): 1153–1154, doi: 10.33963/KP.a2021.0075. 1 case with allergy<br>Witberg G et al. Myocarditis after COVID-19 vaccination in a large health care organization. N Engl J Med. 2021 [Epub ahead of print], doi: 10.1056/NEJMoa2110737. 1 case with biopsy of 54 in series<br>No references for 2 cases in the USA with eosinophilia. |
| Kounis et al. 2021b[14] (letter)<br><br>Hypersensitivity to PEG and tromethamine | • Hypersensitivity or drug induced myocarditis occurs after hypersensitivity reactions to drugs or substances and is neither necrotizing nor fibrotic [7,8]. One third of patients may demonstrate no peripheral eosinophilia and most patients respond well to steroids and drug cessation [9]. Drugs and substances that can cause hypersensitivity myocarditis include vaccines, antibiotics, central nervous system drugs, antitubercular agents and a variety of other undetermined drugs [10]. Hypersensitivity myocarditis can occur in 3% to 10% of cardiac explants and in patients with a ventricular assist device.<br>• Two cases after mRNA vaccination described [by Verma et al.] had endomyocardial biopsies revealing eosinophils and other interacting inflammatory cells such as macrophages, T-cells, and B cells [11].<br>• Lymphocytic myocarditis with presence of macrophages and T cells has been diagnosed after BNT162b2 COVID-19 vaccination, but staining with hematoxylin-eosin to identify eosinophils was not performed [12]. | Supporting:<br>Verma AK, Lavine KJ, Lin CY. Myocarditis after Covid-19 mRNA vaccination. N Engl J Med. 2021;30(385):1332–4. Two cases with eosinophilia on biopsy. |
| Tsilingiris et al., 2021[2] (article)<br><br>Hypersensitivity to PEG and lipid nanoparticle sheath | • The polyethylene glycol (PEG) component and several other ingredients of the lipid nanoparticle sheath have been implicated in other hypersensitivity reactions, most notably in extremely rare but potentially life-threatening immediate cases of anaphylaxis following mRNA vaccine administration [28,29].<br>• It should be noted that in this report and in stark contrast to other available observations a small overall increase in myocarditis risk was observed after the first dose of ChAdOx1 vaccine [34].(Patone et al.) | Supporting:<br><br>Patone et al. Risks of myocarditis, pericarditis, and cardiac arrhythmias associated with COVID-19 vaccination or SARS-CoV-2 infection. Nat Med 2021. https://doi.org/10.1038/s41591-021-01630-0. |
| Bozkurt et al., 2021[5] (narrative review)<br><br>Hypersensitivity: excipients not mentioned | • Reports to date do not suggest a delayed hypersensitivity reaction, such as serum sickness–like reaction or eosinophilic myocarditis as a cause for myocarditis after mRNA COVID-19 vaccination.[15] Although rare, delayed localized skin hypersensitivity reactions have been described with mRNA COVID-19 vaccination with a median latency of 7 days,[59](Johnson et al.) unlike myocarditis emerging earlier within 3 to 4 days after vaccination. None of the case reports published to date had evidence of eosinophilia in peripheral blood or immune complex deposition or eosinophilic infiltrates in endomyocardial biopsy samples arguing against hypersensitivity, allergic or eosinophilic myocarditis.[8–17]<br>• Lipid nanoparticles or adjuvants used in mRNA vaccines have not been shown to result in an immune or inflammatory response and have not been associated with myocarditis either. | Refuting:<br>Several case reports and series (see Hypothesis 2).<br>Johnston MS et al. Delayed localized hypersensitivity reactions to the Moderna COVID-19 vaccine: a case series. JAMA Dermatol. 2021;157:716–720. doi: 10.1001/jamadermatol.2021.1214. Skin reactions rare and delayed more than myocarditis |
| **Hypothesis 5: Response to mRNA vaccine lipid nanoparticles (direct deleterious effect; not delayed – see Hypothesis 4)** | | |
| Tsilingiris et al., 2021[2] | • To counter the inherent instability of free mRNA and facilitate its entry into selected host cells, a lipid nanoparticle sheath is used as a delivery vehicle; the most crucial element of the lipid | Supporting: |

| | | |
|---|---|---|
| (article) | nanoparticles is the variable ionizable lipid (SM-102 for Moderna and ALC-0315 for Pfizer/BioNTech).<br>• The recent observation of a similar adverse event in a recipient of the non-mRNA, peptide-based NVX- CoV2373 in the frame of a phase III clinical trial with 7020 participants in the active treatment arm raises the question whether the lipid nanoparticle sheath, which is a common structural component of these platforms could be implicated in the pathogenesis of vaccine-induced myocarditis.[30] The case of myocarditis within the NVX-CoV2373 clinical trial was reviewed by an independent safety monitoring which determined that it was likely of viral origin and not related to the vaccination itself.<br>• It should be noted that in this report (Patone et al.) and in stark contrast to other available observations a small overall increase in myocarditis risk was observed after the first dose of ChAdOx1 vaccine [34].<br>• One could argue that there have been up until now essentially no reports of a similar clinical picture among receivers of other non-vaccine, LPN-containing treatments. This could be a mere result of the rarity of this adverse event combined with the massive vaccination programs, which could have allowed for the clustering and recognition of such cases. | Patone et al. Risks of myocarditis, pericarditis, and cardiac arrhythmias associated with COVID-19 vaccination or SARS-CoV-2 infection. Nat Med 2021. https://doi.org/10.1038/s41591-021-01630-0. |
| Kadkhoda, 2021[15] (letter) | • A more likely mechanism [than Hypothesis 13 of pericyte expression] is where the vaccine lipid nanoparticles leak from the injection site and enter circulation where clinical injection practices are not very well observed [7]. Then the nanoparticles reach the heart and can be endocytosed by cardiac tissue including cardiac muscle, pericytes, endothelial cells, and macrophages. | None |
| **Hypothesis 6: Autoimmunity triggered by molecular mimicry*** or other mechanism** | | |
| Hajra et al 2021[1] (narrative review)<br><br>Molecular mimicry | • Molecular mimicry: The high prevalence of myocardial damage in COVID-19 [where there is exposure to entire spike protein], combined with a tiny proportion of myocarditis in mRNA COVID-19 vaccine recipients [exposure to partial antigen i.e. small epitope of spike protein], indicates the possibility of molecular mimicry between SARS-CoV-2 spike protein and an unknown myocardial protein [33, 38, 58, 61]. | Supporting:<br>3 case series/reports of myocarditis after mRNA vaccination, indicating lower rates than due to COVID-19:<br>D'Angelo T et al. Myocarditis after SARS-CoV-2 vaccination: a vaccine-induced reaction? Can J Cardiol. 2021<br>Larson KF, Ammirati E, Adler ED, Cooper LT, Hong KN, Sapon- ara G, et al. Myocarditis after BNT162b2 and mRNA-1273 Vac- cination. Circulation. 2021<br>Ammirati E et al. Temporal relation between second dose BNT162b2 mRNA Covid-19 vaccine and cardiac involvement in a patient with previous SARS-COV-2 infection. IJC Heart Vasc. 2021;34:100774. |
| Tsilingiris et al., 2021[2] (article)<br><br>Molecular mimicry and other autoimmune | • Molecular mimicry: Among others, supported by the relatively frequent occurrence of myocardial damage and myocarditis in the frame of SARS-CoV-2 infection, a mechanism of molecular mimicry between the viral S-protein and various self-antigens (i.e., α-myosin) has been suggested [22]. In this case, relatively similar rates of myocarditis occurrence would be expected among receivers of adenoviral vector-based platforms. The currently available evidence presents a rather solid counterargument against this scenario; while cases of myocarditis/pericarditis in association with administration of the ChAdOx1 vaccine (Vaxzevria, Astra- Zeneca) have also been reported [34](Patone et al.), they do not seem to occur more frequently than expected in the absence of vaccination according to most available evidence | Refuting:<br>Molecular mimicry:<br>More cases should occur in non-mRNA vaccines, which introduce spike protein, than have been reported:<br>Patone et al. Risks of myocarditis, pericarditis, and cardiac arrhythmias associated with COVID-19 vaccination or SARS-CoV-2 infection. Nat Med 2021. |

| | | |
|---|---|---|
| | [23,24](Alberta; Australian Government), while there is so far one published only 1 case reported after Janssen Ad26.COV2.S [25].( Sulemankhil et al.)<br>• Other autoimmune: In the foreground stand immune or autoimmune mediated processes as possible mechanisms, and the highest frequency of occurrence after the second vaccine dose (after allowing for a presumed sensitization process to take place after the first dose) seems to strengthen this notion. mRNA vaccines have been already causally implicated in a number of immune-mediated adverse events such as autoimmune thrombocytopenia and thyroiditis [11,21]. | https://doi.org/10.1038/s41591-021-01630-0.<br>Alberta. Office of the chief medical officer of health. Myocarditis and/or Pericarditis following COVID-19 Vaccines 2021. https://www.alberta.ca/assets/documents/health-myocarditis-and-pericarditis-following-covid.pdf.<br>Australian Government. Department of Health. COVID-19 vaccination – guidance on myocarditis and pericarditis after mRNA COVID-19 vaccines. 2021.<br>Sulemankhil I, Abdelrahman M, Negi SI. Temporal association between the COVID-19 Ad26.COV2.S vaccine and acute myocarditis: a case report and literature review. Cardiovasc Revascularization Med : Mol Interv 2021. https://doi.org/10.1016/j.carrev.2021.08.012. |
| D'Angelo et al, 2021[10]<br>(case report and discussion)<br><br>Molecular mimicry | • The pathophysiology of our case was more likely related to an autoimmune phenomenon. Although the exact trigger for autoimmune myocarditis is unknown, literature evidence suggests a "molecular mimicry" when the viral antigen resembles proteins on the myocardium. When autoreactive sensitisation occurs, cytokines and lymphocytes migrate into the myocardial interstitial space, inducing an inflammatory response.[3] | None; nothing from case report to support & reference to influenza vaccine-induced fulminant myocarditis. |
| Heyman & Cooper, 2021[3]<br>(letter)<br><br>Molecular mimicry | • Antibodies directed to SARS- CoV-2 spike glycoproteins might cross-react with structurally similar human protein sequences, including myocardial α- myosin heavy chain. These autoantibodies might be innocent bystanders resulting from myocardial inflammation and injury, or might reflect a certain immune–genetic background that predisposes to developing hyperimmunity and myocarditis upon any trigger.[9] | Supporting:<br>Vojdani, A. & Kharrazian, D. Potential antigenic cross-reactivity between SARS- CoV-2 and human tissue with a possible link to an increase in autoimmune diseases. Clin. Immunol. 2020;217: 108480. In vitro study.** |
| Bozkurt et al., 2021[5]<br>(narrative review)<br><br>Molecular mimicry and other autoimmune | • Molecular mimicry: Another important potential mechanism for myocarditis is molecular mimicry between the spike protein of SARS-CoV-2 and self-antigens.[50] Antibodies against SARS-CoV-2 spike glycoproteins have been experimentally shown to cross-react with structurally similar human peptide protein sequences, including α-myosin.[50](Vojdani et al.) However, severe adverse events or autoimmune reactions have been very rare.[46,47](Polack et al. and Baden et al.)<br>• Other autoimmune: (One case) had higher levels of antibodies against some self-antigens such as aquaporin 4, endothelial cell antigen, and proteolipid protein 1.[17](Muthukumar A et al) In the patient studied, autoantibody levels peaked on day 2 along with symptoms, but they did not recede as expected, as the clinical condition improved, although the follow-up was rather short. Also, the autoantibodies may not be pathogenic and could also be seen as a result of myocardial inflammation. (Historically, circulating heart-reactive autoantibodies have been reported at a higher frequency in patients with myocarditis and have been implicated in pathogenesis. These autoantibodies are usually directed against multiple antigens, some of which may have functional effects on cardiac myocytes.[49]) Autoantibodies are found more frequently in first-degree relatives of patients with cardiomyopathy than in the healthy | Supporting:<br>Molecular mimicry:<br>Vojdani, A. & Kharrazian, D. Potential antigenic cross- reactivity between SARS- CoV-2 and human tissue with a possible link to an increase in autoimmune diseases. Clin. Immunol. 2020;217: 108480. In vitro study (see row immediately above for details).<br>Other autoimmunity:<br>Muthukumar A et al. In-depth evaluation of a case of presumed myocarditis after the second dose of COVID-19 mRNA vaccine. Circulation. 2021;144:487–498. |

| | | |
|---|---|---|
| | population, raising the possibility that myocarditis may develop in a subgroup of patients with the appropriate genetic background. | |
| Chouchana et al., 2021[11]<br>(retrospective study on Vigibase case and discussion)<br><br>Molecular mimicry | • The mRNA is known to be a self-adjuvant for innate immune responses, and this may help to explain their immunogenicity, and trigger excessive immune responses in some individuals, especially when there may be presence of a cross-reacting antigen. | None |
| Switzer & Loeb, 2021[8]<br>(narrative review)<br><br>Molecular mimicry and other autoimmune | • Molecular mimicry: A potential avenue for vaccine-associated myocarditis may be a nonspecific innate inflammatory immune response, or perhaps an interaction between the encoded viral spike protein of the mRNA and an as-yet undetermined cardiac protein [21,56]. Studies have hypothesized that the antibodies generated in response to the mRNA spike protein may react with surface antibodies of the cardiomyocytes of susceptible hosts, provoking an inflammatory reaction and associated tissue damage [21,57].<br>• Other autoimmune: Heart-reactive auto-antibodies have been reported at elevated levels in patients with myocarditis [2,57,64]. These antibodies may target multiple antigens, possibly having functional effects on cardiac myocytes and contributing to the pathogenesis of vaccine-induced. | None |
| Parra-Lucares et al., 2021[4]<br>(case report and narrative review)<br><br>Molecular mimicry and other autoimmune | • Molecular mimicry: The presence of mimicry between the spike protein and cardiac autoantigens (e.g., myosin) generates anti-SARS-CoV-2 antibodies with affinity to cardiac proteins, inducing an autoimmune humoral response. In in vitro studies [68] (Vojdani et al), anti-SARS-CoV-2 antibodies have been shown to crosstalk with human proteins, such as alpha-myosin, a structural protein of cardiomyocytes involved in myocardial muscle contraction. *However, to date, it has not been shown that these antibodies can generate an autoimmune response in tissues that express these proteins, both in animal models and in patients*.<br>• Other autoimmune: The presence of antibodies against self-antigens was evaluated in the clinical case described above [64](Muthukumar A et al.). Autoantibodies such as anti-aquaporin 4, anti-endothelial antigen, or anti-proteolipid protein 1 were detected. These autoantibodies have been previously reported in patients with myocarditis [69] and first-degree relatives of patients with myocarditis, which supports the existence of a myocarditis mechanism mediated by autoantibody formation. *However, it has not been demonstrated that these autoantibodies can cause an autoimmune response in organisms, both in the heart and other tissues, so it could only be a non-causal correlation*.<br>• Other autoimmune: In most cases [of patients with clinical and laboratory findings of myocarditis associated with anti-SARS-CoV-2 vaccination], significant alterations in autoimmune parameters observed in other pathologies were not detected, including rheumatoid factor (RF), antinuclear antibodies (ANA), or elevation of inflammatory parameters (C-reactive protein or erythrocyte sedimentation rate). | <u>Supporting:</u><br>Molecular mimicry:<br>Vojdani, A. & Kharrazian, D. Potential antigenic cross- reactivity between SARS- CoV-2 and human tissue with a possible link to an increase in autoimmune diseases. Clin. Immunol. 2020;217: 108480. In vitro study showing cross-reactivity with at least one protein in muscle, i.e. α-myosin (see above); caution about unknown implications.<br><br>Other autoimmune:<br>Muthukumar A et al. In-depth evaluation of a case of presumed myocarditis after the second dose of COVID-19 mRNA vaccine. Circulation. 2021;144:487–498. Case report with detected autoantibodies; caution about unknown implications.<br><br><u>Refuting</u>:<br>Other autoimmune:<br>Indicative of direct data but no citations |
| Ehlrich et al., 2021[16]<br>(case report in 40 year-old male after first dose, with biopsy) | • Case report of biopsy-proven (left ventricular endomyocardial) lymphocytic myocarditis in 40-yr male after first dose. Histology and immuno-histology of the biopsies revealed acute lymphocytic myocarditis. As the patient developed myocarditis a few days after the first vaccination in absence of anti-SARS-CoV-2-antibodies, the pathogenesis of mRNA COVID-19 vaccine associated myocarditis does not appear to depend on anti-SARS CoV-2 spike protein antibodies. Thus, the hypothesis of cross-reactivity of antibodies induced by mRNA vaccination with myocardial antigens (molecular mimicry [7]) is not corroborated by our case. | <u>Refuting</u>:<br>Molecular mimicry (after first dose):<br>Their case report data, due to lack of anti-SARS-CoV-2-antibodies |

| | | |
|---|---|---|
| Molecular mimicry | Rather, the quick cardiac infiltration of immune cells after vaccination suggests that myocarditis may be caused by other mechanisms. | |
| **Hypothesis 7: Low residual levels of double-strand RNA (dsRNA)** | | |
| Milano et al 2021[17] (special report) | • The presence of low residual levels of double-strand RNA (dsRNA) has been reported in mRNA COVID-19 vaccine preparations…dsRNA is known to be a strong exogenous inducer of immune-inflammatory reactions involving well-identified intracellular signaling cascades and mediators.[17]<br>• The current methods used to purify IVT mRNA vaccine preparations vary in terms of technical performance and, at best, allow the removal of 90% of dsRNA when using HPLC, as reported by the developers of mRNA vaccines [17].<br>• dsRNA is detected by antigen-presenting cells, endothelial cells and the airway epithelium [18], and gives rise to dose-related innate immune activation [17]. When packaged in lipid nanoparticles, dsRNA is preferentially transferred to phagocytic monocytic-derived cells, such as macrophages and dendritic cells, which are key actors in immunity [24].<br>• However, a relatively low level of clinical evidence is currently available in this [COVID-19 mRNA vaccines] context to be taken as hypothesis-generating. | None |
| **Hypothesis 8: Dysregulated micro-RNA response** | | |
| AbdelMassih et al. 2021[18] (literature review) | • MicroRNAs are short non-coding RNAs that play a crucial role in the regulation of gene expression during cellular processes. It is now established that some of the host-generated miRNAs are known to modulate the antiviral defense during viral infection. Recently, multiple DNA and RNA viruses have been shown to produce miRNAs known as viral miRNAs (v-miRNAs). viral RNA can either alter the expression of host miRNA or use cellular machinery to form viral miRNAs. We hypothesize that mRNA vaccines can either trigger the release of host miRNAs or contain themselves some miRNAs that can trigger [myocarditis].<br>• [In conclusion] the evidence reveals that the micro-RNAs implicated in myocarditis in general are as well implicated in the pathogenesis of severe COVID-19, this can explain why patients having a first dose with a history of COVID-19 can develop myocarditis from mRNA vaccines, also the relatively higher likelihood of this complication in males and younger aged individuals can be explained by the upregulation of key myocarditis related miRNAs in those two strata, due to higher muscle mass and suggests performing a sarcopenia index in recipients of the vaccine to correlate it with the likelihood of this complication. | None |
| **Hypothesis 9: Production of anti-idiotype antibodies against immunogenic regions of antigen-specific antibodies** | | |
| Tsilingiris et al., 2021[2] (article) | • This process could in theory lead to tissue-specific adverse events through the formation of immune complexes, activation, blockade and/or down-regulation of membrane receptors (e.g. ACE2), as well as complement- or immune cell-mediated cellular damage [26]. | None |
| **Hypothesis 10: Trigger of pre-existing dysregulated immune pathways in certain individuals with predispositions (e.g., resulting in a polyclonal B-cell expansion, immune complex formation, and inflammation[5])** | | |
| Bozkurt et al., 2021[5] (narrative review) | • Although nucleoside modifications of mRNA have been shown to reduce their innate immunogenicity,45 in certain individuals with genetic predisposition,48 the immune response to mRNA may not be turned down and may drive the activation of an aberrant innate and acquired immune response. The dendritic cells or Toll-like receptor expressing cells exposed to RNA may still have the capacity to express cytokines and activation markers in certain individuals, although this may be markedly less when exposed to mRNA with nucleoside modifications than when treated with unmodified RNA. The immune system may therefore detect the mRNA in the vaccine as an antigen, resulting in activation of proinflammatory | Refuting:<br>For specific predispositions:<br>Abu Mouch S et al. Myocarditis following COVID-19 mRNA vaccination. Vaccine. 2021;39:3790–3793. Case series n=6<br>Muthukumar A et al. In-depth evaluation of a case of presumed myocarditis after the second dose of COVID-19 mRNA vaccine. Circulation. 2021;144:487–498. |

| | | |
|---|---|---|
| | cascades and immunologic pathways that may play a role in the development of myocarditis as part of a systemic reaction in certain individuals.[45,48]<br>• [In 6 male cases of COVID mRNA vaccine myocarditis in Israel], serology for autoimmune disorders with antinuclear antibodies and rheumatoid factor were negative, with no evidence of predilection to individuals with pre-existing autoimmune disorders.[10](Abu Mounch et al.)<br>• In 1 case report (Mathukumar et al.), a panel testing for variants in 121 genes potentially linked to cardiomyopathy was negative,[17] arguing against an existing predisposition to cardiomyopathy attributable to known gene variants in that case. | |
| Switzer & Loeb, 2021[8] (narrative review) | • It is possible that genetic factors regulating the inflammasome activation, or interferon-signaling cascade, may contribute to an individual's risk of developing the cytokine storm responsible for triggering auto-reactive cell activity after exposure to the mRNA vaccine [58, 61, 63]. | None |
| **Hypothesis 11: Antibody-dependent enhancement of immunity or other forms of immune enhancement with re-exposure to virus after vaccine** | | |
| Bozkurt et al., 2021[5] (narrative review) | • No evidence of either cellular immune enhancement or antibody-dependent enhancement of immunity was observed in non-human primate studies after SARS-CoV-2 virus challenge, either after vaccination [not specific to approved mRNA vaccines] or previous infection.[58] These findings led a National Institutes of Health ACTIV study (Accelerating COVID-19 Therapeutic Interventions and Vaccines) panel to conclude that the risk of immune enhancement after COVID-19 immunizations was low, but required ongoing pharmacovigilance and monitoring.[58] To date, neither COVID-19 disease nor the new COVID-19 vaccines have shown evidence of causing antibody-dependent enhancement of immunity or other forms of immune enhancement with re-exposure. People infected with SARS-CoV-2 have not been reported to develop antibody-dependent enhancement of immunity on repeat exposure, and vaccine breakthrough COVID-19 cases are rare and mild. There is no evidence of acute COVID-19 infection during presentation with myocarditis cases after COVID-19 vaccination, arguing against a breakthrough infection as a cause (Table 4 review of available cases reports and series) | Refuting:<br>Multiple case reports and series reviewed and tabulated, having no evidence of acute COVID-19 infections after vaccine when presenting with myocarditis. |
| **Hypothesis 12: Direct cell invasion via the spike protein interacting with the angiotensin-converting enzyme 2 (ACE2) widely expressed and prevalent in cardiomyocytes[11]** | | |
| Chouchana et al., 2021[11] (retrospective study on Vigibase case and discussion) | • In two recently reported cases of myocarditis following mRNA vaccination, only inflammatory infiltration was assessed in the myocardium, suggesting that the ACE2 hypothesis is probably not relevant.[46] | Refuting:<br>Verma, A.K et al., Myocarditis after Covid-19 mRNA Vaccination. N. Engl. J. Med. 385, 1332–1334 (2021). Data from 2 case reports; only inflammatory infiltration was assessed in the myocardium |
| Switzer & Loeb, 2021[8] (narrative review) | • Encoded viral surface spike protein of the mRNA vaccine, which triggers the immune response, may interact with ACE2 receptors in the host, increasing the likelihood of cardiac sensitivity or inflammatory reactions [38,39]. Possible host genetic factors in ACE2 receptors, which vary across ethnic groups, may drive increased susceptibility to elevated cardiovascular symptoms or the development of an inflammatory response triggering symptom onset [39,52,58]. | None |
| **Hypothesis 13: Cardiac pericyte expression of ACE2** with **immobilized immune complex on the surface of pericytes activation of the complement system** | | |
| Kadkhoda et al., 2021[15] (letter) | • The role of pericytes in susceptibility to COVID-19 through the expression of SARS-CoV-2 receptor, i.e., angiotensin-converting enzyme 2 (ACE2) has been demonstrated [4]. It has also been shown that after infection with SARS-CoV-2, anamnestic humoral immune responses to previously-encountered common coronaviruses (CoVs) is augmented significantly [6]. Anti- | None |

| | | |
|---|---|---|
| | spike antibodies elicited as a result of past exposure to common CoVs and/or to SARS-CoV-2 spike (be it through prior infection or vaccination), may elicit anti-idiotype antibodies, that is, antibodies directed against the paratope region of anti-spike antibodies. Since the latter is the mirror image of the anti-spike antibodies, it may mimic the spike protein itself and bind ACE2 expressed on cardiac pericytes that express ACE2. This forms an immobilized immune complex on the surface of pericytes. This localized immune complex, in turn, may lead to activation of the complement system through its classical pathway and damage to the target cell. | |
| **Hypothesis 14: Spike-activated neutrophils (expressing ACE2) augmenting inflammatory response** | | |
| Kadkhoda et al., 2021[15] (letter) | • Local production of spike protein on the surface of cardiac cells and/or its shedding along with detached cell membranes may recruit neutrophils that also express ACE2 on their surface. Spike-activated neutrophils produce neutrophil extracellular traps [8] that subsequently activate alternative pathway of complement in situ, damaging cardiac endothelial cells. | None |
| Choi et al., 2021[19] (case report) | • There were three main histological findings in the heart: 1) myocarditis predominantly involving the atrial wall, with neutrophil and histiocyte predominance; 2) non-inflammatory single-cell necrosis; and 3) diffuse CBN [contraction band necrosis] throughout the myocardium, predominantly in the left ventricle... In this case, the myocarditis was histologically different from viral or immune-mediated myocarditis in that the inflammatory infiltrates were predominantly neutrophils and histiocytes, rather than lymphocytes...The underlying mechanism of myocardial injury in this case is unclear, but it may have involved cytokine-mediated or histiocyte-linked immunologic injury to the myocardium. | <u>Supporting:</u><br>Autopsy findings from Choi case report (22 year-old male); inflammatory infiltrates were predominantly neutrophils and histiocytes, rather than lymphocytes. |
| **Hypothesis 15: Hyperviscosity-induced cardiac problem** | | |
| Mungmunpuntipantip & Wiwanitkit, 2021[20] (letter to the editor) | The underlying mechanism of post COVID-19 vaccination hyperviscosity is a change of antibody level in plasma after vaccine stimulation. In the case of underlying high blood viscosity or previous COVID-19, the excessive increasing of antibody level might occur and can result in excessive blood viscosity and hyperviscosity.[2,3] | None |
| **Hypothesis 16: Strenuous exercise induced secretion of proinflammatory IL-6** | | |
| Elkazzaz et al., 2022[21] (protocol for retrospective and prospective observational study) | • Cytokine storm is suggested as one of the major pathological characteristics of SARS-CoV-2 infection, It was found that the presence of SARS-CoV-2 spike protein in epithelial cells promotes IL-6 trans-signaling by activation of the AT1 axis to initiate coordination of a hyper-inflammatory response [17].<br>• Also, It was showed that increase of TNF-a and IL-6 was found after the 1st vaccination in individuals with pre-existing COVID-19 immunity(18) and also, IL-6 were significantly higher after the second COVID vaccination dose of S-Protein Based Vaccines for COVID-19 at day 23 than those at day 2 [18].<br>• Compared to the DNA vaccine, the mRNA vaccine induced a more robust production of IL-5, IL-6 [19].<br>• Pro-inflammatory cytokines IL-6, TNF-α, a heterodimeric cytokine belonging to the IL-12 family were increased early upon vaccine administration [20].<br>• Exercise causes skeletal muscle cells to release IL-6, and it raises the plasma concentration of IL-6 100 times higher than at rest [23]. Strenuous exercise raises levels of a variety of pro- and anti-inflammatory cytokines. The concentration of IL-6 increases up to 100-fold after strenuous exercise, such as a marathon race [3,4].<br>• In addition to the induction effect of COVID-19 vaccine on IL-6, strenuous exercise (and muscle contraction) could boost the effect of IL-6 leading to myocarditis. | None |

| Differences in incidence by sex could be due to sex steroid hormones or underdiagnosis in females | | |
|---|---|---|
| Tsilingiris et al., 2021[2] (article) | • In order to explain the skewed gender distribution of cases, the influence of sex steroid hormones (estrogen, testosterone) has been suggested [34]. | None; cited reference does not refer to or investigate sex hormones |
| Heymans & Cooper, 2021[3] (letter) | • Differences in hormone signalling might be involved in the pathophysiology of COVID-19 mRNA- vaccination- related myocarditis. Testosterone can inhibit anti- inflammatory immune cells and promote a more aggressive T helper 1 cell- type immune response. By contrast, oestrogen has inhibitory effects on pro- inflammatory T cells, resulting in a decrease in cell-mediated immune responses.[1] | None |
| Bozkurt et al., 2021[5] (narrative review) | • Sex hormones: An important possible explanation relates to sex hormone differences.3,65,66 Testosterone is thought to play a role, by a combined mechanism of inhibition of anti-inflammatory cells [3,65–67] and commitment to a Th1-type immune response.[68] Estrogen has inhibitory effects on proinflammatory T cells, resulting in a decrease in cell-mediated immune responses; and pericarditis incidence is higher in women during the postmenopausal period.[69]<br>• Underdiagnosed in women: Another contributing factor could be underdiagnosis in women. By our analysis of the VAERS database, as of June 6, 2021, there were 6235 reported cases of chest pain, 69% of which were in women, versus 30% in men.[70] Despite a higher prevalence of chest pain in women, diagnostic evaluation, including ECG, laboratory biomarkers, echocardiography, and MRI, was performed and reported more often in male than in female patients presenting with chest pain after COVID vaccination (Bozkurt, unpublished data, 2021). | <u>Supporting</u>:<br>Sex hormones: None<br><br>Underdiagnosis in women:<br>Centers for Disease Control and Prevention. The Vaccine Adverse Event Reporting System (VAERS) results. June 6, 2021. Accessed July 6, 2021. https://wonder.cdc.gov/vaers.html. Mor chest pain complaints in females.<br>Bozkurt, unpublished data, 2021. Fewer investigations in females. |
| Chouchana et al., 2021[11] (retrospective study on Vigibase case and discussion) | • Although, female patients usually generate higher overall antibody levels and more adverse events following vaccination, male patients have increased enhanced type-1 immune responses.[47] These differences may be driven by sex hormone differences and testosterone is thought to play a role in commitment to a Th1 response.[38] | None |
| Parra-Lucares et al., 2021[4] (case report and narrative review) | • Testosterone has been observed to exhibit inhibitory effects on anti-inflammatory cells, increased activity of pro-inflammatory M1 macrophages, and increased CD4+ type 1 (Th1) T lymphocyte response [70]. In turn, estrogens have an inhibitory effect on pro-inflammatory T lymphocytes, causing a decrease in the cellular immune response. This fact explains the observation that the highest incidence of myocarditis or pericarditis (not specific to mRNA COVID) in women occurs in those of postmenopausal age [72]. However, given the characteristics of the published reports (several of these coming from studies carried out in soldiers, for example) [39,73], there is a significant selection bias, so it is not yet possible to confirm whether this complication is more frequent in the male population. | <u>Refuting</u>:<br>Montgomery J et al. Myocarditis Following Immunization with MRNA COVID-19 Vaccines in Members of the US Military. JAMA Cardiol. 2021, 6, 1202–1206. Source population biased towards males (but many other population-based studies exist now). |
